# Early Stopping in Experimentation with Real-time Functional Magnetic Resonance Imaging Using a Modified Sequential Probability Ratio Test

**DOI:** 10.1101/2021.01.15.21249886

**Authors:** Sarah J. A. Carr, Weicong Chen, Jeremy Fondran, Harry Friel, Javier Sanchez-Gonzalez, Jing Zhang, Curtis Tatsuoka

**Author notes:** Corresponding Author: Curtis Tatsuoka, 10900 Euclid Avenue, Case Western Reserve University, Cleveland, OH, USA 44106.

## Abstract

**Introduction:** Functional magnetic resonance imaging (fMRI) often involves long scanning durations to ensure the associated brain activity can be detected. However, excessive experimentation can lead to many undesirable effects, such as from learning and/or fatigue effects, discomfort for the subject, excessive motion artifacts and loss of sustained attention on task. Overly long experimentation can thus have a detrimental effect on signal quality and accurate voxel activation detection. Here, we propose dynamic experimentation with real-time fMRI using a novel statistically-driven approach that invokes early stopping when sufficient statistical evidence for assessing the task-related activation is observed.

**Methods:** Voxel-level sequential probability ratio test (SPRT) statistics based on general linear models (GLMs) were implemented on fMRI scans of a mathematical 1-back task from 12 healthy teenage subjects and 11 teenage subjects born extremely preterm (EPT). This approach is based on likelihood ratios and allows for systematic early stopping based on statistical error thresholds. We adopt a two-stage estimation approach that allows for accurate estimates of GLM parameters before stopping is considered. Early stopping performance is reported for different first stage lengths, and activation results are compared with full durations. Finally, group comparisons are conducted with both early stopped and full duration scan data. Numerical parallelization was employed to facilitate completion of computations involving a new scan within every repetition time (TR).

**Results:** Use of SPRT demonstrates the feasibility and efficiency gains of automated early stopping, with comparable activation detection as with full protocols. Dynamic stopping of stimulus administration was achieved in around half of subjects, with typical time savings of up to 33% (4 minutes on a 12 minute scan). A group analysis produced similar patterns of activity for control subjects between early stopping and full duration scans. The EPT group, individually, demonstrated more variability in location and extent of the activations compared to the normal term control group. This was apparent in the EPT group results, reflected by fewer and smaller clusters.

**Conclusion:** A systematic statistical approach for early stopping with real-time fMRI experimentation has been implemented. This dynamic approach has promise for reducing subject burden and fatigue effects.

## 1.0 Introduction

Analysis of task-based fMRI scans is typically performed with fixed, predetermined experimental designs. As a result, subjects must often endure stimulus protocols that are overly long in order to ensure the neural activity can be statistically discerned in the noisy data. However, this can lead to fatigue, learning effects and excessive motion, such as from agitation, as well as being costlier to administer due to longer scan times and potentially less reliable measurement. Also, the experimenter does not know if the neural activity is detectable until long after the scanning session is over. Real-time functional MRI (RT-fMRI) provides an opportunity to ameliorate these issues. RT-fMRI has been successfully applied in the field of neurofeedback and biofeedback from neural responses, where subjects may be trained to alter their brain activity based on real-time information provided from the fMRI scans. This has been reported in ADHD (1), healthy subjects with no psychiatric or neurological disorders (2, 3), Alzheimer’s disease (4) and Parkinson’s disease (5, 6). Its uses have also been described in psychoradiology to aid diagnosis and treatment planning in psychiatric disorders (7). Real-time resting state fMRI has for instance been studied and implemented as well using TurboFIRE (8). A largely unexplored application of RT-fMRI is to dynamically and statistically determine when a stimulus has been sufficiently presented in terms of replication of blocks to terminate early. The magnitude of effort and variability in neural activity while completing a task will vary from person to person. Trial administration within a block design can be stopped early if sequentially updated statistical inference on activation can be determined with sufficient accuracy based on the observed BOLD (blood oxygen level dependent) signal response up to that point. This application will be explored in detail.

The benefits of adaptive RT-fMRI include: 1<u>) Shorter scan times for fMRI testing</u>: Shorter scan times cannot only save in technology and personnel costs, but fatigue and learning effects can be avoided, improving signal quality. Scanning becomes less burdensome on the subject as well, which is an especially important consideration for children or elderly subjects. 2) <u>Real-time quality control</u>: greater consistency in activation classification error can be obtained, through statistical error-based benchmarks for stopping rules and real-time feedback on classification performance and adjustment of stimulus durations. 3) <u>Richer information</u>: Paradigms can become more complex and sophisticated. With greater time efficiency and flexibility, more variations of a stimulus, such as reflected by a broader range of difficulty levels, can be administered in the same amount of time. 4) <u>Wide applicability</u>: Dynamic adjustment of stimuli based on BOLD response in real time can be generally applied across a range of focus areas that investigate localization of brain activity, including cognition and motor functioning.

Since the advent of RT-fMRI in the mid 1990’s (9), a handful of mainstream software packages have been developed for use by the fMRI community. These include Turbo BrainVoyager (10), AFNI’s real-time plugin (9) and FSL-based FRIEND (11). There have been a few previous studies that have employed adaptive task-based RT-fMRI. In one example, it has been used to determine ‘good’ and ‘bad’ brain states to optimize learning (12). The presentation of novel scenes was prompted by the detection of ‘good’ brain states; the ‘good’ template was determined based on a prior standard acquisition test scan. They used real-time general linear model (GLM) methods described in (13) to estimate the BOLD signal magnitude at each time point (each scan) and compared it to a value within a region of interest from the earlier test scan. Another adaptive RT-fMRI study has used a person’s brain state to judge their attention to a task (14). When their attention appeared to wander, the difficulty of the task was increased bringing their attention back. The authors applied multivariate pattern analysis to determine task-relevant and task-irrelevant activity. In another example, Lorenz et al (2016) ran FSL to pre-process the scans in real-time before applying a GLM-based analysis. Their study involved eliciting activity in particular brain regions by presenting stimuli chosen based on the response to the previous stimulus. The aim was not to investigate brain activity related to a particular task but simply activate a brain region (15). Another example of adaptive RT-fMRI implemented a Bayesian optimization algorithm to estimate when brain activity was mapped to a particular network (16). The Bayesian optimization was trained on 4 difficulty levels of a task prior to switching to choosing the optimal difficulty levels to elicit the desired activity, where there were 12 other levels to choose from.

Here, we extend the use of a statistically-based dynamic approach to RT-fMRI experimentation described in (17), addressing issues related to practical implementation. This approach involves the sequential updating of voxel-level likelihood ratio tests, known as sequential probability ratio tests (SPRTs) and assessing after each scan whether there is sufficient statistical evidence to determine whether or not an associated parameter value indicates task activation. Such results, considered in aggregate across a collection of voxels, can be used as a basis for early stopping of experimentation. Most off-line, post-hoc analyses of fMRI data use the general linear model to test statistical associations of voxel activation magnitude to task administration (18-20). This approach involves the voxel-level estimation of task-related regression parameters that indicate magnitude of association between an expected hemodynamic response signal from a task and the observed BOLD signal. We have adapted this general method for real-time fMRI by sequentially updating GLM regression parameter estimates as soon as the brain volumes are collected. At the individual voxel level, we can then assess hypothesis tests related to activation that are based on these estimates. In aggregate, the voxel level analyses inform decisions on early stopping and the tailoring of fMRI experimentation (17).

In comparison to (17), we adopt a two-stage estimation approach that allows for the alternative hypothesis test parameter values that represent activation thresholds to be formulated in terms of z-score scale at the voxel level. This adaptive specification avoids the intractable problem of pre-specifying magnitudes of GLM parameter values that would be considered as “active”. Such magnitudes need to be scaled relative to error variance, which is estimated in a first stage. We determine an appropriate duration of the first stage by monitoring estimation convergence of key GLM parameters. Also, while in (17) serial independence was assumed, here we use the “sandwich” estimator to recognize serial covariance in inference (21, 22). The impact of early stopping on group analysis is considered here as well. Importantly, we now present a novel workflow to apply and implement these methods on a Philips scanner, with a dynamic feedback system that allows for real-time dynamic adjustment of the experimentation with subjects. This was facilitated with adoption of numerical parallelization techniques. This work supports the premise that adaptive, individualized experimentation is feasible and can lead to practical and useful savings in scan times by reducing experimental redundancy.

Another novel aspect of this work is the application of adaptive RT-fMRI in a sample group of 12 healthy adolescent subjects and 11 adolescents born extremely preterm (EPT). The fMRI stimulus was a mathematical version of the well-known 1-back task. Early stopping was implemented using sequential probability ratio test (SPRT) statistics and our server was a Linux workstation located in a nearby building. Processing of RT-fMRI was completed within 3 seconds before the next scan arrived. We observed time savings of up to 33 % based on early stopping when 80% of voxels were classified, which equals up to 4 minute savings with a 12-minute scan. The impact on activation analysis from the selection of early stopping criteria is assessed, as described in detail below. Finally, we conduct a comparison of group analyses between EPT versus healthy controls, to assess the effects of early stopping in this context.

## 2.0 Background Information

### 2.1 General linear model

Briefly, the general linear model involves convoluting a double gamma hemodynamic response function (HRF) with task indicator variables that denote timing of administration to reflect expected task-related BOLD responses. Voxel-level task-related regression parameters are estimated and represent the association of the observed response to expected task-activated BOLD signal. Thus, activation is assessed through statistical inference on regression parameters. For a given voxel up to time t (i.e. for scans 1 through t), the GLM takes the form:

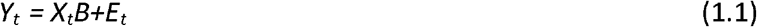

Where *Y*_*t*_ is a *t* × 1 vector of observed BOLD signal intensities for the voxel up to time *t*, and *E*_*t*_ is a *t* × 1 vector that represents the error components. *X*_*t*_ is a *t* × *p* design matrix and includes the expected BOLD signal values per task. We also include cosine functions of increasing periodicity (scan duration*2, scan duration, scan duration/1.5, scan duration/2 and scan duration/2.5) to model physiological and other low frequency noise (23). For large periodicities, cosine functions are approximately linear for the time frame of scans we consider here, and hence are essentially collinear from a GLM modeling perspective. Five regressors were thus added to the design matrix. *B* [*b*_1_ …*b*_*j*_ …*b*_*p*_]^*t*^, a p × 1 regression coefficients vector. In this formulation, a regression parameter *b*_*j*_ can represent magnitude of association with task j. *E*_*t*_ is assumed to be distributed as multivariate normal with mean zero and covariance *W*_*t*_, where *W*_*t*_ is a *t* × *t* matrix that represents the temporal autocorrelation structure. For spatial correlation, we conduct spatial smoothing, so do not explicitly model the spatial correlation structure. *Y*_*t*_ is assumed to have a multivariate normal probability distribution as follows:

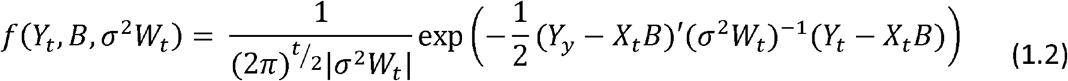

where |σ^2^*W*_*t*_| is the determinant of σ^2^*W*_*t*_. Major sources of noise in fMRI data include brain metabolism, physiology, and spontaneous fluctuations (24).

We fit regression models in parallel for all voxels under consideration in a target region of interest (ROI), which could include the whole brain. Real-time analysis requires signal and image processing steps, as well as the continual updating of statistical estimates as new scan data are received from the scanner. Hence, given the large number of voxels to be analyzed, real-time fMRI presents “big data” computational challenges.

### 2.2 Sandwich Estimator

In our previous work (17), we assumed serial independence for computational simplicity. Here we recognize potential serial correlation using the nonparametric sandwich estimator 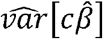 for contrast *c* (21, 22). The sandwich estimator is a robust, model-free variance estimator that does not require distributional assumptions. Importantly, it still provides asymptotically consistent variance estimates, although convergence rates can be slow (21, 22). The approach is computational feasible for real time analysis.

### 2.3 Wald’s Sequential Probability Ratio Test

At the voxel level, we can use the sequential analytic framework of (17, 25-29), to adaptively assess activation status using real-time fMRI. As we will demonstrate, Wald’s SPRT test statistic can serve as the basis of an efficient, sequential testing approach that can greatly reduce the need for experimental block administrations compared with fixed designs while attaining similar classification performance in simulation, and activation patterns with subject data. This approach relies on a SPRT statistic to conduct hypothesis testing, with the null hypothesis representing no activation with respect to a task, and the alternative hypothesis representing some threshold of activation, as represented by a GLM parameter value (17). This statistic is updated with each new observation, and its value is compared with thresholds for stopping.

The general procedure of Wald’s SPRT is described as follows. Consider a one-sided hypothesis *H*_0_: *c*′*β* = *c*′*β*_0_ of the form versus *H*_*a*_: *c*′*β* ≥*c*′*β*_1_ where *c* ′(*β*_1_ − *β*_2_) ≥ 0. Two-sided formulations are described in (25) and (17). Implementation of Wald’s SPRT involves updating Wald’s likelihood ratio statistic as new data are observed (25):

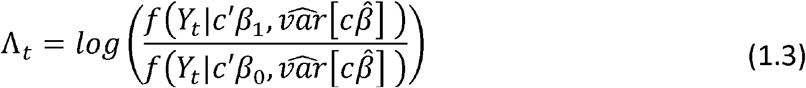

where 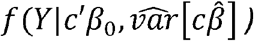 and 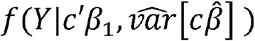 are the respective probability densities functions of *Y*_*t*_ given *c*′*β*_0_ or *c*′*β*_1_ is the true value of parameter of interest and conditioning on the estimated covariance. After *Y*_*t*_ is observed at a time point, *t*, one of three possible decisions is made according to the following rules:

1. Continue sampling if *B* < Λ_*t*_ < *A*
2. Stop sampling and accept *H*_0_ if Λ_*t*_ < *B*
3. Stop sampling and accept *H*_*a*_ if *A* < Λ_*t*_

where stopping boundaries (*A, B*) = (log((1-β_E_)/α_E_), log(β_E_/(1-α_E_))), and the target Type I and Type II error levels are respectively denoted as α_E_ and β_E_. These error levels are specified before testing. Note that both the Type I and Type II error levels are controlled for with SPRT, as opposed to standard hypothesis test formulations that only control for Type I error level. Multiple SPRTs are conducted concurrently across voxels and boundary error levels can be adjusted for instance by Bonferroni correction to account for this simultaneous testing.

A practical modification of the original SPRT formulation for stopping is to consider the truncated SPRT (30), which will additionally call for stopping if an upper bound for the number of observations is reached. In our case, this is reached when a fixed number of blocks have been administered. Additional modifications include conducting two-stage estimation to allow sufficient observation for preliminary estimates of the voxel-level error variance from a first stage where stopping is not yet considered (31). With these estimates, we can derive an alternative hypothesis value for a linear contrast of task parameters *c*′*β* that will correspond to a desired z-statistic value. As an illustration, suppose a z-statistic value of 3.10 is selected, as will be done below in our studies. Note 3.10 is the one-sided p-value = 0.001 - critical value for the standard normal distribution. Given an estimated value 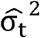 from a first stage of length t scans, we solve for the value of θ_*t*_ = *c*′*β* that satisfies 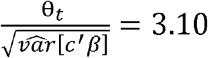, where *X*_*t*_ is the design matrix up to scan *t*. This value becomes the alternative hypothesis, and it represents the voxel-level targeted activation magnitude threshold. We update the value of θ_*t*_ and 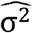 at each scan, so that the alternative hypothesis is actually dynamic, since the estimation variance for 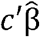 changes as well.

Ultimately, we aggregate the findings of the voxel-level SPRTs to determine whether or not experimentation within a block design should be terminated early. A “global” stopping rule that considers all voxels in a region of interest (can be whole brain or smaller ROIs) that we have adopted is to terminate task administration when a predetermined percentage of voxels have been classified by their respective SPRTs. For instance, we have used 80% as a global stopping criterion. Note that 80% classified means either as active or non-active. We choose this cut-off as it is fairly strict, and yet approximately one half of the participants still stop early. As we will see, it also facilitates correspondence with full scan data results, particularly if the activation threshold is adjusted to recognize longer scan durations. We also consider other global stopping criterion here, 70% and 90%, and assess impact on stopping times and resultant images arising from early stopping. We also choose Type I and Type II error levels that are relatively more stringent for Type I error. Note that for *c*′*β* parameter values that are “in-between” the null and alternative hypothesis values, the SPRT is indifferent to preferring one hypothesis over the other. This leads to larger numbers of scans needed before a stopping boundary is crossed. So, we have to accept a lack of decisive stopping decisions for these cases in order for overall experimentation to stop early, even as *θ*_*t*_ decreases as t increases. This can be an acceptable trade-off for shorter experimental scan times and the ability to tailor experimentation.

In sum, we propose that the important design parameters for implementation are selected through analysis of a training sample. For training, each subject undergoes the full duration of experimentation. We consider selection through the following criteria: 1) First stage duration: It is desirable for the voxel-level error variance and beta parameter estimates to stabilize – we assess this qualitatively by assessing plots from a sample of voxels. 2) A z-score activation threshold based at the end of the first stage: we choose a z-score threshold of 3.10 after the first stage since this is a standard threshold value for determining activation of a voxel. Note that thresholds at earlier scans correspond to even larger z-score thresholds at later durations, as we discuss below. 3) Type I and Type II error levels: We want to observe some level of early stopping based on these parameters while the corresponding activation maps with early stopping appear have correspondence to full duration scans (after threshold adjustment for larger number of scans). 4) Global stop rule percentage: a percentage level is selected by relying on similar guidance as when selecting the hypothesis testing error levels.

## 3.0 Methodology

### 3.1 Participants

Twelve healthy subjects were recruited, 7 males. They were aged 15-16 years old and 11 were right-handed. They had no known neurological conditions and a normal developmental history. A group of 11 adolescents born EPT were also recruited, 1 male. EPT is defined as being born at < 26-week gestation and weighing < 1000g. All were aged 15-17 years old and 8 were right-handed, 2 left-handed and 1 ambidextrous. All subjects were recruited as part of a larger study to evaluate functional and structural differences associated with mathematical abilities and working memory between those born EPT and those born at normal term. The aim of the larger study is to improve our understanding of mathematics disabilities and potentially lead to improvements in pedagogical practices for young people experiencing problems acquiring mathematics skills. Adolescents were recruited as they can handle the stress of fMRI experimentation, are mathematically advanced enough and have had time to master the subject area compared to younger children. This age range is also an advantageous time to implement interventions to improve mathematical abilities before leaving school, hence adults were not studied. EPT subjects were included to show that differences with patient populations are detectable with our methods. A subsection of the full study is reported here to demonstrate the real-time analysis.

The subjects made one two-hour visit to the MRI department at University Hospitals Cleveland Medical Center (UHCMC). Ethics approval was obtained from the UHCMC Institutional Review Board office prior to the study and complied with the Declaration of Helsinki for human subject research. Subjects and their parents gave informed consent prior to taking part.

As part of our wider study, subjects also made another, separate 3 hour visit to the study offices to undergo neuropsychological testing and a refresher of fraction calculations. In the interests of brevity, the full neuropsychological testing results are not reported here. One finding that is particularly relevant to the fMRI task considered here is that nearly two thirds (63.6 %) of the EPT cohort have lower working memory function, compared to just over one third (35.7 %) of controls subjects.

### 3.2 MRI protocols

The subjects were positioned head-first supine on the scanner bed with their head fixed in position using inflatable pads. An 8-channel head coil was used for data acquisition. Echo planar imaging scans were acquired on a Philips Ingenuity 3T PET/MR imager at UHCMC. The following fMRI scan parameters were used: TR = 3.0 s, TE = 35 ms, in-plane resolution was 1.797 mm^2^ (matrix 128 × 128), slice thickness was 4 mm, number of slices = 36 slices and flip angle = 90°. A SENSE P reduction factor of 2 was implemented and scans were acquired in an ascending interleaved fashion.

In addition to the fMRI scans, a high-resolution T1-weighted anatomical image of the brain was also acquired. This was taken using a magnetic preparation gradient-echo sequence (3D IR TFE). Imaging parameters were: TR = 7.5 ms, TE = 3.7 ms, in-plane resolution was 1 mm^2^ (matrix 256 × 256), slice thickness was 1 mm, number of slices = 200 slices and flip angle = 8°.

### 3.3 Stimulus protocols

During data acquisition subjects were presented with a mathematical version of the well-known 1-back memory task. It involved performing basic addition and subtraction calculations and required the answer to be remembered and compared to the next answer. Two difficulty levels were included. The protocol was developed by our lab as part of a battery to assess mathematical and working memory abilities in 14 – 17 year olds to evaluate the functional differences between those born EPT and those born at normal term. The stimulus was presented on an MRI compatible LCD monitor (manufactured by Cambridge Research Systems, Rochester, UK) positioned at the end of the bore and viewed via a mirror attached to the head coil. Equations were presented, for example, the subject may see “2 + 3 = ?”. The subject was required to work out the answer and then remember it while working out the next equation, for example “1 + 4 = ?”. If they thought the answers matched, then the subject pressed a button on a response box held in their right hand. If they thought the answers did not match, then they did nothing but remember the new answer to compare to the answer of the next equation. An example sequence is shown in Figure 1A.

**Figure 1:**
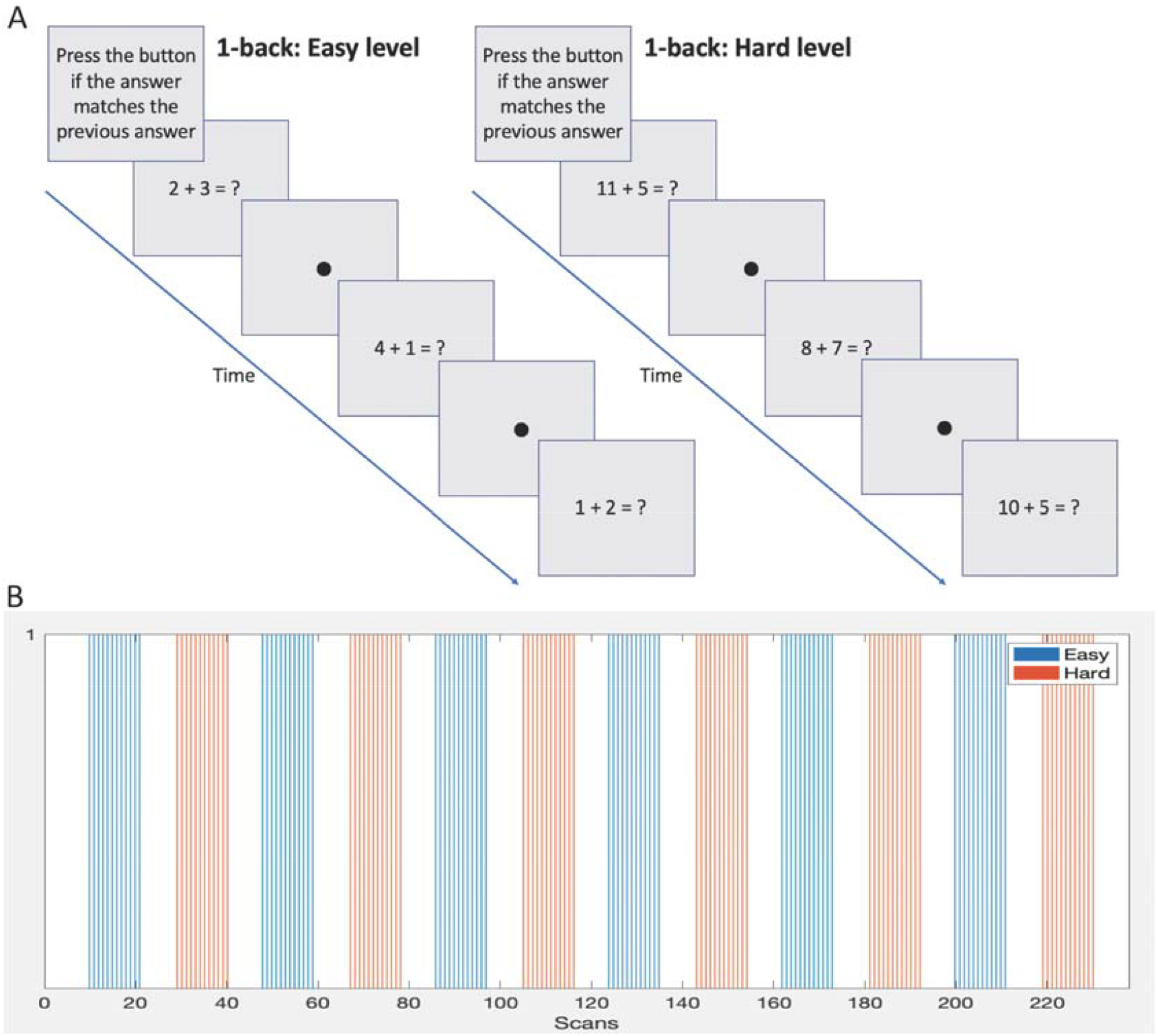
A) Sample 1-back protocols demonstrating the two difficulty levels. B) Block design and timings of each difficulty level.

The stimulus was presented in a block design, see Figure 1B and Table 1. Eight equations were presented per block. Each block lasted 36 seconds followed by 21 seconds of rest condition (fixation dot). Two difficulty levels were presented. The easier level consisted of single digit numbers to add or subtract and the answers were always a single digit. The harder level involved addition or subtraction of single or two-digit numbers and the answers were always two digits. Blocks of difficulty levels were alternated during the scan and a total of 6 blocks per level were presented. Note: although only 2 difficulty levels are used here, the setup is able to accommodate any number of difficulty levels. The full duration of the task was 238 scans or 11 minutes and 54 seconds. This was based on a moderate length of experimentation for a 1-back block design (e.g. see (32-35)), allowing approximately 6 minutes for each difficulty level.

**Table 1:** The scan number when each stimulus block is completed.

| Block | Easy level | Hard level |
| --- | --- | --- |
| 1 | 21 | 40 |
| 2 | 59 | 78 |
| 3 | 97 | 116 |
| 4 | 135 | 154 |
| 5 | 173 | 192 |
| 6 | 211 | 230 |

Two difficulty levels were included to investigate differences in neural responses associated with increasing task demand. As the brain is ‘pushed’ to solve more complex problems, differential networks may be apparent, and these may be different between normal term and EPT subjects. Additionally, increasing the difficulty level serves to maintain the subject’s attention and, generally, increases their effort. This can have the effect of increasing brain activation cluster sizes and magnitude as well as causing recruitment of additional areas, which is of interest. Incorporating difficulty levels into protocols that can be terminated early in a separate fashion demonstrates the flexibility of the proposed approach.

### 3.4 Real-time fMRI acquisition

The visual stimulus was presented using an in-house custom written program that was developed using the Python programming language (Python Software Foundation, https://www.python.org/) and libraries from Psychopy - an open source visual presentation program (36-38). The program connected to a Cedrus Lumina controller to receive stimulus responses from the subject and trigger pulses from the MRI scanner (outputted every dynamic). The timing of the presentation of the visual stimulus was synchronized to the trigger pulses to ensure that stimulus images were displayed at the expected time. A Supervisor Window displayed on the experimenter’s computer screen allowed the visual stimulus to be tracked throughout. It displayed the current block number being presented, how many remaining blocks there were and when the subject responded. The program was also able to terminate one or both of the difficulty levels if it received a signal indicating the relevant areas in the fMRI data were sufficiently classified across voxels. The software is freely available from the Bitbucket repository: https://bitbucket.org/tatsuoka-lab/fmri-presentation.

Real-time image transfer was achieved by XTC (eXTernal Control). This is a program integrated into the Philips scanner software and enabled by a research clinical science key. XTC communicates with the reconstruction and scanner processes on the scanner computer and interfaces to a network Client application using a minimalistic CORBA (Common Object Request Broker Architecture) (39) interface which uses TCP/IP as the transport layer. CORBA is platform independent, reliable, and has the ability to process large amounts of data with minimum overhead. Each CORBA message consisted of a hierarchical attribute collection identified with UUIDs (universally unique identifiers) (40). Messages carried reconstructed image data and meta-data containing details of scan protocols. Due to hospital network security protocols the reconstructed images were placed in a folder on the scanner computer and then pushed across to a Linux computer. To achieve necessary image transfer speeds to the scanner computer folder a modification to XTC was installed on the scanner to disable two-way communications as only one-way image transfer functionality was required. However, XTC does support two-way communication between the scanner and the Client.

The Linux computer was a custom-built server equipped with a solid state hard drive and two 8-core Intel Xeon E5-2687W processors running at 3.1 GHz and providing 40 MB L3 cache. It was installed with Centos 7.4 operating system. As the scans were received, custom written Python and Bash scripts implemented the analysis using core-based parallelization to preprocess the data and perform the SPRT statistical analysis. Preprocessing was performed using standard modules from AFNI (Analysis of Functional NeuroImages, https://afni.nimh.nih.gov) and FSL (FMRIB’s Software Library, https://fsl.fmrib.ox.ac.uk/fsl/fslwiki/). The analysis sequence is detailed in the following section. The setup is shown in Figure 2.

**Figure 2:**
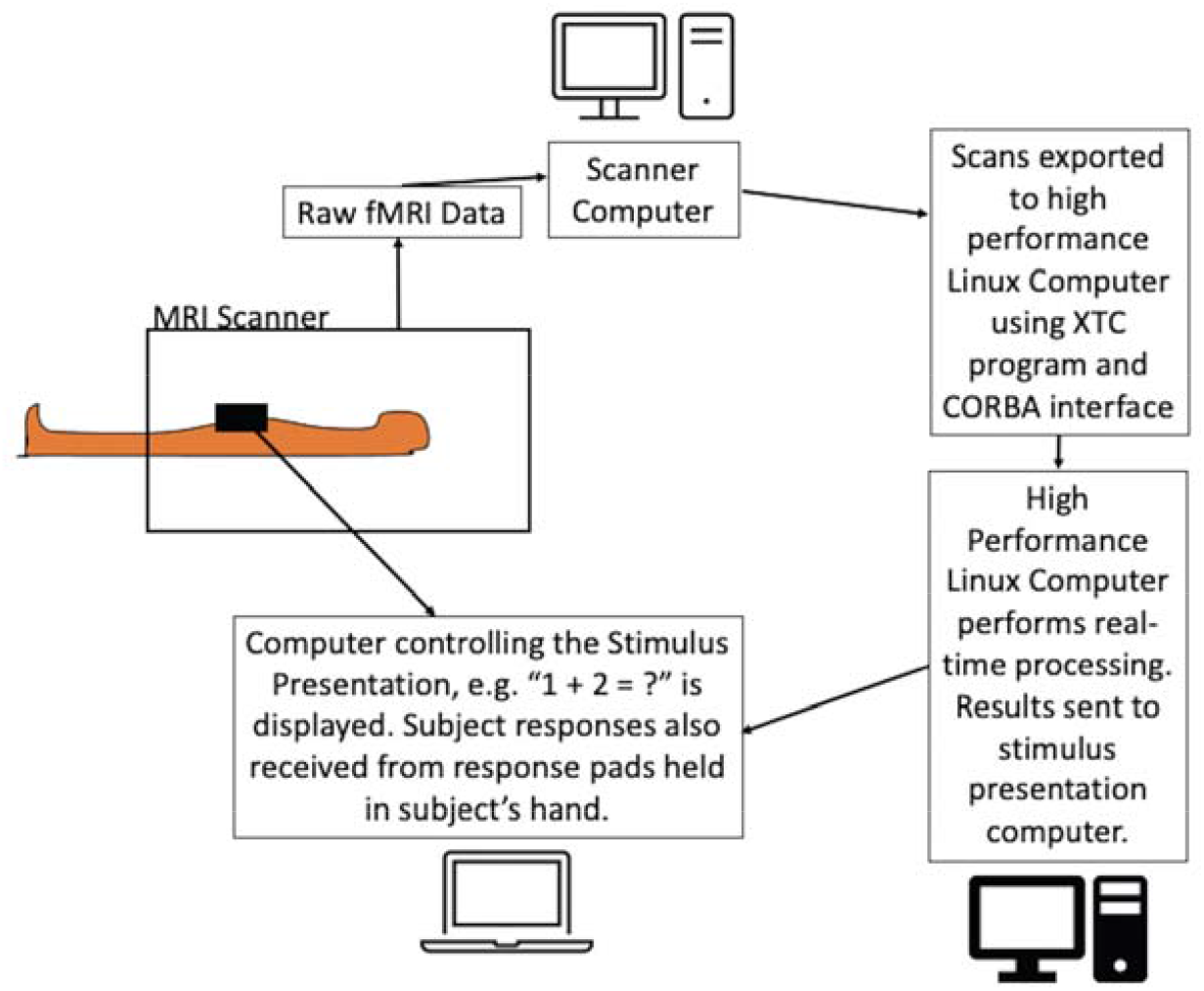
Schematic of the experimental setup of the dynamic real-time fMRI process. The equations were presented to the subject while the scans were acquired using a dedicated computer. FMRI scans were exported in real-time from the scanner computer to the Linux workstation using the Philips XTC program and CORBA interface. Scans were preprocessed on the Linux workstation and SPRT statistics were calculated. The results were relayed back to the stimulus presentation program with an instruction to either continue or terminate the stimulus.

### 3.5 MRI preprocessing

At the beginning of the scanning session a single fMRI scan (3 seconds) was acquired and used for coregistration (motion correction) purposes. In preparation, the skull was removed using FSL’s Brain Extraction Tool (BET) (41) and a mask of the full brain was created. During the real-time adaptive fMRI scan session, new scans arrived every 3 seconds and were dumped in a folder on the Linux workstation where the following actions were applied to each one. AFNI’s ‘dcm2niix_afni’ command was used to convert the .par/.rec files to nifti. Motion correction was performed using coregistration techniques. Every fMRI scan was realigned to the initial scan that was acquired before the task began, and AFNI’s ‘3dvolreg’ command was used. Spatial smoothing was also applied using an 8 mm kernel with AFNI’s ‘3dmerge’ command. The full brain mask created at the beginning of the session was applied using FSL’s ‘fslmaths’ command to remove noisy voxels outside the brain (voxels of no interest). The resulting images were then converted to ascii format for statistical analysis with SPRT.

### 3.6 fMRI SPRT analysis

The SPRT analysis was applied using highly-optimized C++ program that used Intel Cilk Plus library for multicore and vector processing of data. BLAS routines from Intel MKL were used to enable instruction-based acceleration for matrix computation. They are available from the Bitbucket repository at https://bitbucket.org/tatsuoka-lab. The design matrix was created prior to the scan session using AFNI’s ‘3dDeconvolve’ command to model the stimulus and HRF. It is possible to include the temporal derivatives of the HRF or other regressors in the design matrix where applicable in studies. Temporal derivatives were not included here due to the long durations of the block design used to present the task. Statistical analysis included the modeling of low frequency physiological noise and the associated removal of serial correlation using discrete cosine transforms. Motion parameters are also frequently used as regressors to remove correlated activations produced by movement. Here motion parameter regressors were not included with the estimation of the discrete cosine transforms due to the limitations of the computational resources.

We also tested 2 scenarios using either 2-blocks or 4-blocks of easy and hard stimuli first stage administration before allowing early stopping to occur. Where 2-blocks per difficulty level of stimulus administration were used before allowing early stopping, the first 78 scans were used for the first stage of experimentation. Where 4-blocks were used, 154 scans were used for the first stage. Recall, a full-length task protocol comprised 238 scans and lasted 11 minutes and 54 seconds.

The automatic determination of when to terminate the scanning is based on the Type I and Type II errors, α_E_ and β_E_, as described above. Typical values used in the literature were used to test stopping time performance, with α_E_ = 0.001, β_E_ = 0.1 (42, 43). We also considered α_E_ = 0.0001, β_E_ = 0.1 and α_E_ = 0.001, β_E_ = 0.01 combinations as well. A percentage of voxels that must be classified before termination was also specified during the setup, such as 80%. We also evaluate 70% and 90% threshold levels.

This 1-back arithmetic task involves not only number sense and mathematical calculations but also general cognitive skills involving working memory and sustained attention. The brain networks involved with each of these has been well characterized in the literature and lends itself to the evaluation of this real-time analysis method. There is a large amount of overlap for the active brain areas that control each of these functions and they appear as a frontoparietal network (44-46). The areas of the brain we expect to see activate in response to the experimental task are: the intraparietal sulcus, supramarginal gyrus, premotor cortex, dorsal/ventral lateral prefrontal cortex, parietal lobe, Broca’s area, occipital lobe, fusiform gyrus, precuneus, cingulate gyrus, anterior insula and frontal eye fields. Assessment of the location and extent of activations within this network will be used as additional criteria for judging appropriate stopping times, in addition to the statistical information determined through the SPRT analysis. This will include how well the cluster peaks coincide with the anatomical locations as well as their extent.

### 3.7 Group analysis

There are many possible applications in the research setting where individual level results may be the focus. A possible clinical application may be in clinical assessments for presurgical evaluation for brain surgery in patients with brain cancer or epilepsy. Still, group analyses are commonly conducted and an essential aspect of fMRI analyses. The outputted results files from the SPRT analysis can be used directly to perform a group analysis using AFNI’s 3dMEMA command (Mixed Effects Meta Analysis tool) (47). However, a group analysis was carried out using FSL which instead merges all subject data to conduct a combined mixed models analysis. We demonstrate that the data collected in real-time can still be used in a typical post-hoc analysis. Raw data was preprocessed with FSL FEAT (48). Motion correction was performed using a rigid body transform, spatial smoothing with a full-width-at-half-maximum Gaussian kernel of 6 mm was applied, high pass temporal filtering of 90 s was carried out and coregistration to (MNI) standard space was done before performing a first level individual GLM analysis. The statistical output from these were used to perform the higher level group statistics using FLAME 1 (FMRIB’s Local Analysis of Mixed Effects, (49)).

## 4.0 Results

### 4.1 Individual Subject Results of SPRT

The median control subject response time across both difficulty levels was 1.44 sec (SD 0.51 sec), and median task accuracy was 90.8 % (SD 20.2 %). When these are broken down by difficulty level, the easy level median task accuracy was 86.1 % (SD 22.6 %) with median response time of 1.28 sec (SD 0.54 sec); and the hard level median task accuracy was 90.0 % (SD 18.4 %) with median response time of 1.56 sec (SD 0.51 sec). EPT subjects had a slightly longer overall median response time of 1.91 sec (SD 0.48 sec) and overall median task accuracy was lower at 65.8 % (SD 21.2 %). For the easy level, the median accuracy was 72.2 % (SD 24.2 %) and median response time was 1.63 sec (SD 0.49 sec). For the hard level the median accuracy was 70.0 % (SD 19.8 %) with a median response time of 2.10 sec (SD 0.54 sec). Note that there are statistically significant differences in same subject differences in speed to completion by difficulty level (Wilcoxon signed rank test, two-sided p < 0.001). Comparing correctness percentages per subject across birth status groups, there are significant differences with the hard level (Mann Whitney two-sided p = 0.037), but not with the easy one (two-sided p = 0.401). These results indicate that the difficulty levels have different psychometric properties, and affect the groups differently. We also see this in activation patterns, as discussed in Section 4.2 and reflected in the group analysis results.

Real-time transfer speeds between the scanner and the Linux computer were consistently fast, with individual scan files taking less than 150 milliseconds to transfer. All subject scans were processed within the 3 second TR period. Offline testing showed that the subject with the largest number of voxels (subject 21 with 135,379 voxels) was processed in just 5 minutes and 45 seconds, or 1.45 seconds per scan. The subject with the fewest number of voxels (subject 14 with 77,359 voxels) was processed in 5 minutes and 2 seconds, or 1.27 seconds per scan. Therefore, for the subject with the largest number of voxels, the maximum time to process 1 scan in real-time would be 1.6 sec (1.45 processing time + 0.150 transfer time). Thus, it is feasible for a TR of 2 seconds or faster to be used with the software, depending on transfer speeds and the number of voxels in the brain.

Inspection of the z-score maps for each subject showed that generally, across subjects, the largest activations were centered bilaterally around the inferior and superior parietal areas, taking in the intraparietal sulcus, a region highly associated with mathematical functioning. Further activations were seen in the cuneus. These are most likely correlated with the visual processing associated with the task. Additional activations were seen in the precuneus, bilateral areas in the medial frontal gyrus, anterior cingulate, insula and inferior frontal gyrus. These areas are often associated with attention and memory systems (44, 50).

The stopping times for the 2- and 4-block first stage lengths are reported in Table 2. First, as we see for instance in Figure 3, error variance estimate is not stable after a 2-block first stage. It is important to “wait” until this happens, as it plays a central role in inference and on test statistic values. The 4-block first stage is more attractive in this way. Table 3 shows how early stopping is affected by the SPRT Type I and Type II error threshold values. Note that early stopping does not occur for Type II error levels of 0.01 and is less affected by the Type I error specification.

**Table 2:**
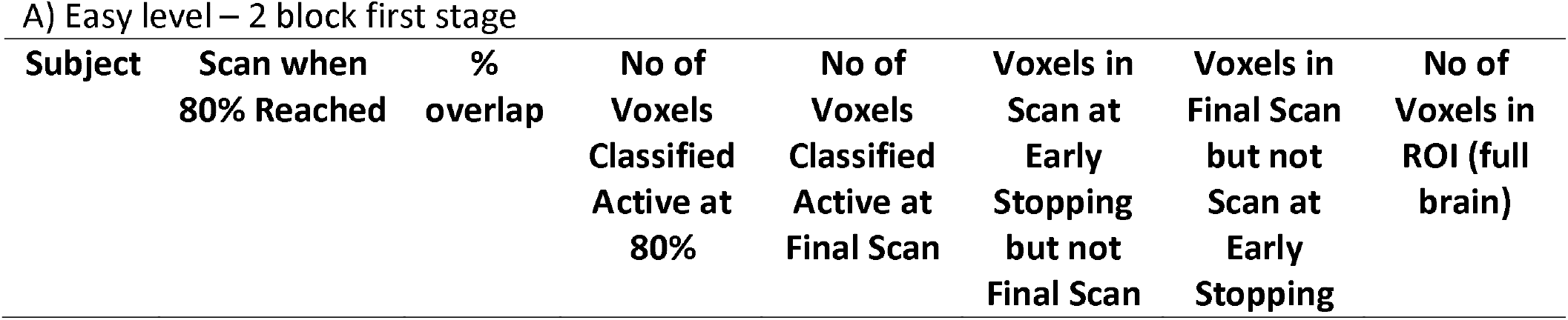
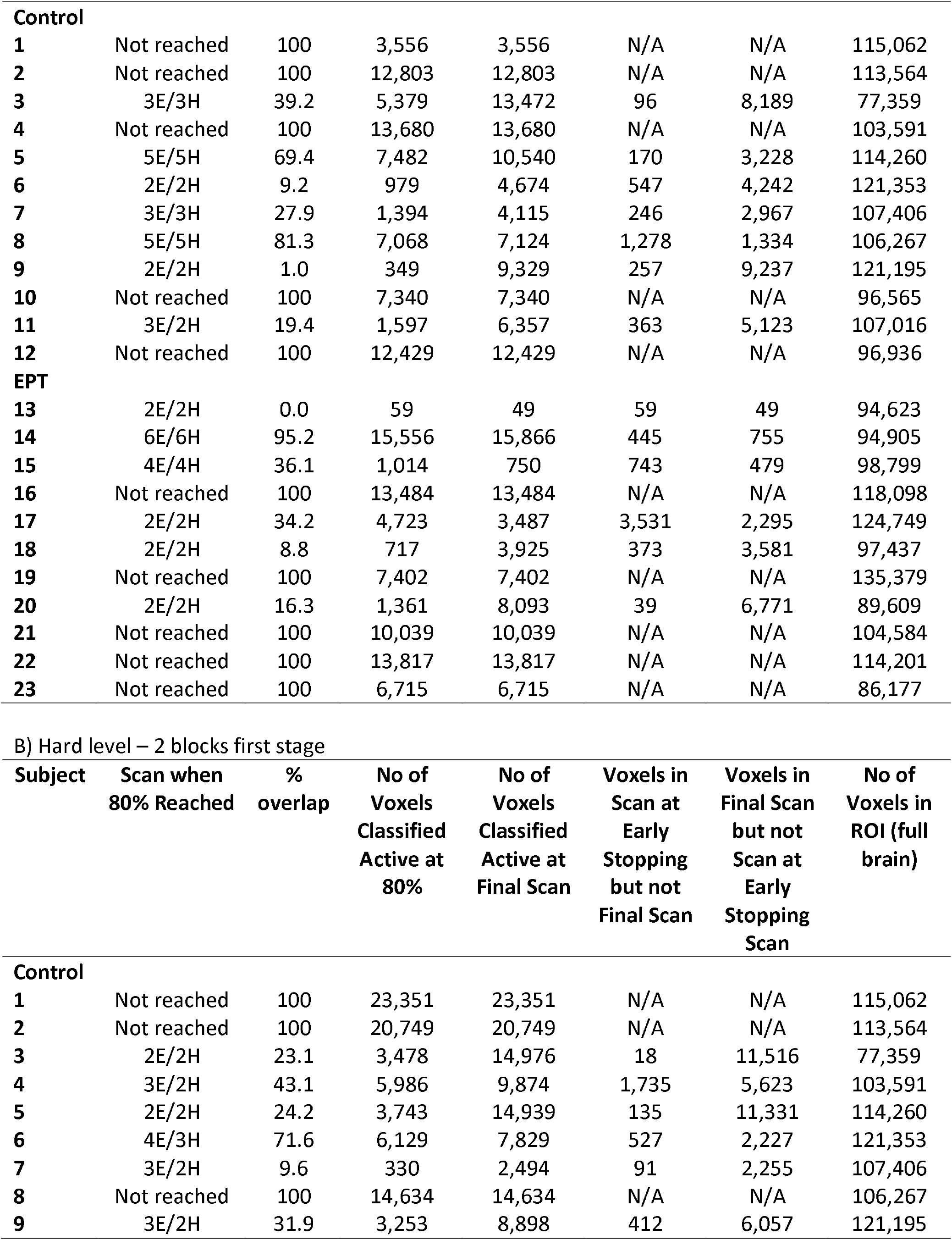
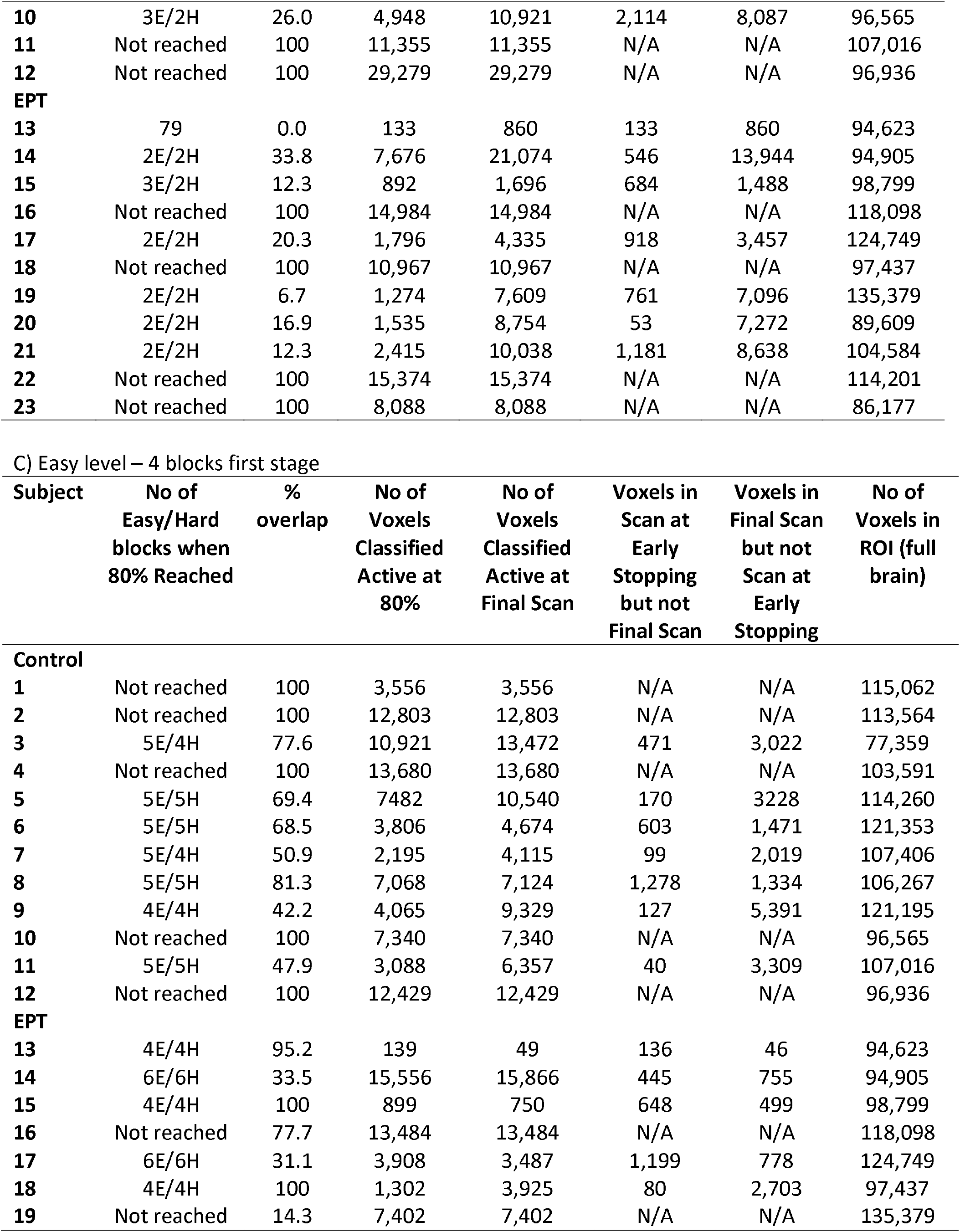
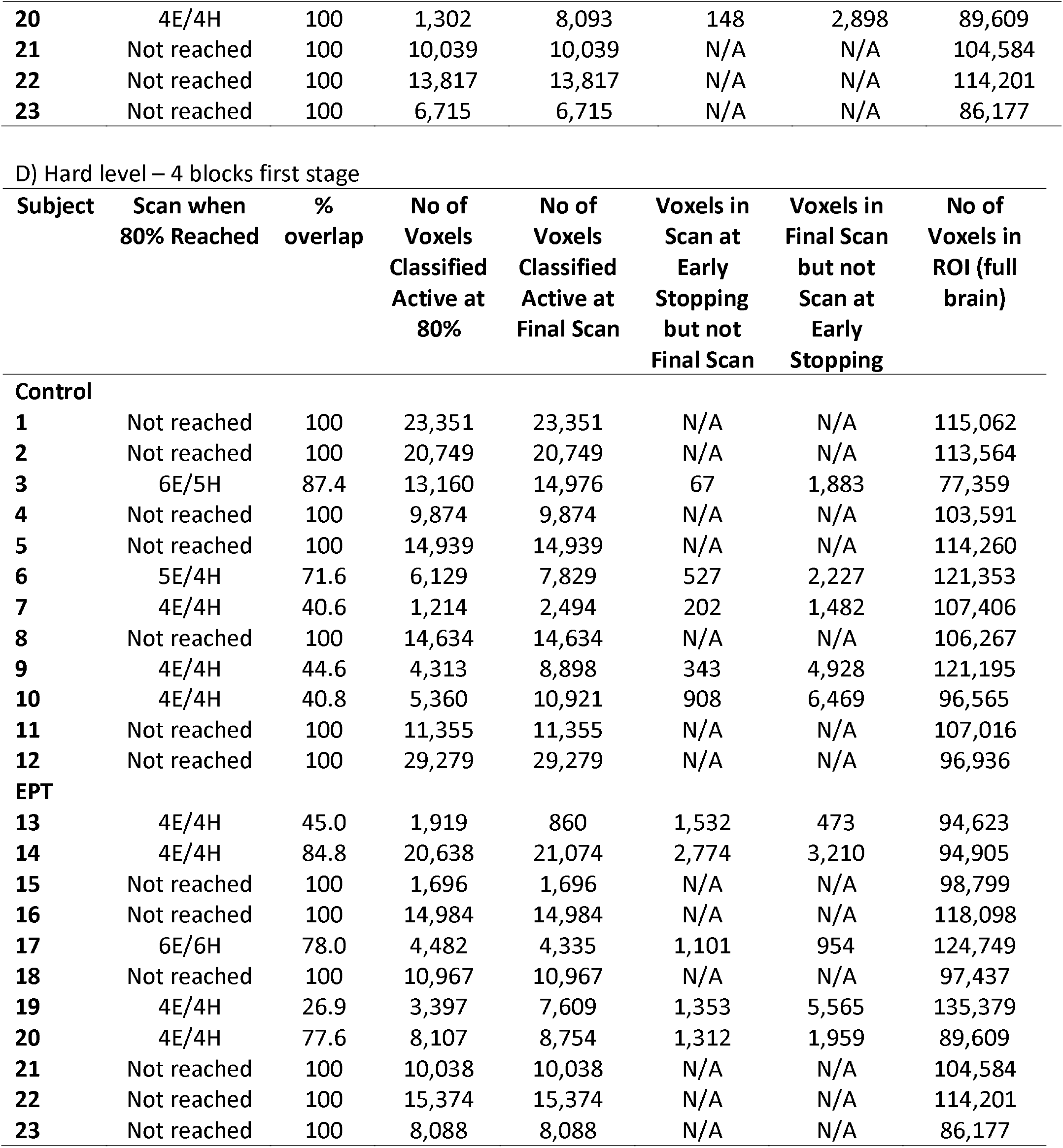
Subject results of the 1-back task using SPRT to analyse the data. Analysis reported here uses α_E_ = 0.001, β_E_ = 0.1 and thresholded at p < 0.001. A) Easy level: 2-block, B) Hard level: 2-block, C) Easy level: 4-block and D) Hard level: 4-block. After each subject’s last administered block, the number of active voxels that spatially overlap between early stopping and full duration are given. The percentage of voxels-in-common is also given relative to the total number of active voxels at the full duration scan. Maximum number of possible scans is 238, minimum is 78 scans for 2 blocks first stage of easy and hard stimulus administration or 154 scans for 4 blocks first stage of easy and hard. Median values are calculated with those who stopped early only. Information given for the point where 80% of voxels have been classified as either active or non-active. N/A = not applicable.

| Subject | Scan when<br>80% Reached | %<br>overlap | No of<br>Voxels<br>Classified<br>Active at<br>80% | No of<br>Voxels<br>Classified<br>Active at<br>Final Scan | Voxels in<br>Scan at<br>Early<br>Stopping<br>but not<br>Final Scan | Voxels in<br>Final Scan<br>but not<br>Scan at<br>Early<br>Stopping | No of<br>Voxels in<br>ROI (full<br>brain) |
| --- | --- | --- | --- | --- | --- | --- | --- |
| **Control** |  |  |  |  |  |  |  |
| **1** | Not reached | 100 | 3,556 | 3,556 | N/A | N/A | 115,062 |
| **2** | Not reached | 100 | 12,803 | 12,803 | N/A | N/A | 113,564 |
| **3** | 3E/3H | 39.2 | 5,379 | 13,472 | 96 | 8,189 | 77,359 |
| **4** | Not reached | 100 | 13,680 | 13,680 | N/A | N/A | 103,591 |
| **5** | 5E/5H | 69.4 | 7,482 | 10,540 | 170 | 3,228 | 114,260 |
| **6** | 2E/2H | 9.2 | 979 | 4,674 | 547 | 4,242 | 121,353 |
| **7** | 3E/3H | 27.9 | 1,394 | 4,115 | 246 | 2,967 | 107,406 |
| **8** | 5E/5H | 81.3 | 7,068 | 7,124 | 1,278 | 1,334 | 106,267 |
| **9** | 2E/2H | 1.0 | 349 | 9,329 | 257 | 9,237 | 121,195 |
| **10** | Not reached | 100 | 7,340 | 7,340 | N/A | N/A | 96,565 |
| **11** | 3E/2H | 19.4 | 1,597 | 6,357 | 363 | 5,123 | 107,016 |
| **12** | Not reached | 100 | 12,429 | 12,429 | N/A | N/A | 96,936 |
| **EPT** |  |  |  |  |  |  |  |
| **13** | 2E/2H | 0.0 | 59 | 49 | 59 | 49 | 94,623 |
| **14** | 6E/6H | 95.2 | 15,556 | 15,866 | 445 | 755 | 94,905 |
| **15** | 4E/4H | 36.1 | 1,014 | 750 | 743 | 479 | 98,799 |
| **16** | Not reached | 100 | 13,484 | 13,484 | N/A | N/A | 118,098 |
| **17** | 2E/2H | 34.2 | 4,723 | 3,487 | 3,531 | 2,295 | 124,749 |
| **18** | 2E/2H | 8.8 | 717 | 3,925 | 373 | 3,581 | 97,437 |
| **19** | Not reached | 100 | 7,402 | 7,402 | N/A | N/A | 135,379 |
| **20** | 2E/2H | 16.3 | 1,361 | 8,093 | 39 | 6,771 | 89,609 |
| **21** | Not reached | 100 | 10,039 | 10,039 | N/A | N/A | 104,584 |
| **22** | Not reached | 100 | 13,817 | 13,817 | N/A | N/A | 114,201 |
| **23** | Not reached | 100 | 6,715 | 6,715 | N/A | N/A | 86,177 |

B) Hard level – 2 blocks first stage
| **Subject** | **Scan when**<br>**80% Reached** | **%**<br>**overlap** | **No of**<br>**Voxels**<br>**Classified**<br>**Active at**<br>**80%** | **No of**<br>**Voxels**<br>**Classified**<br>**Active at**<br>**Final Scan** | **Voxels in**<br>**Scan at**<br>**Early**<br>**Stopping**<br>**but not**<br>**Final Scan** | **Voxels in**<br>**Final Scan**<br>**but not**<br>**Scan at**<br>**Early**<br>**Stopping**<br>**Scan** | **No of**<br>**Voxels in**<br>**ROI (full**<br>**brain)** |
| --- | --- | --- | --- | --- | --- | --- | --- |
| **Control** |  |  |  |  |  |  |  |
| **1** | Not reached | 100 | 23,351 | 23,351 | N/A | N/A | 115,062 |
| **2** | Not reached | 100 | 20,749 | 20,749 | N/A | N/A | 113,564 |
| **3** | 2E/2H | 23.1 | 3,478 | 14,976 | 18 | 11,516 | 77,359 |
| **4** | 3E/2H | 43.1 | 5,986 | 9,874 | 1,735 | 5,623 | 103,591 |
| **5** | 2E/2H | 24.2 | 3,743 | 14,939 | 135 | 11,331 | 114,260 |
| **6** | 4E/3H | 71.6 | 6,129 | 7,829 | 527 | 2,227 | 121,353 |
| **7** | 3E/2H | 9.6 | 330 | 2,494 | 91 | 2,255 | 107,406 |
| **8** | Not reached | 100 | 14,634 | 14,634 | N/A | N/A | 106,267 |
| **9** | 3E/2H | 31.9 | 3,253 | 8,898 | 412 | 6,057 | 121,195 |
| **10** | 3E/2H | 26.0 | 4,948 | 10,921 | 2,114 | 8,087 | 96,565 |
| **11** | Not reached | 100 | 11,355 | 11,355 | N/A | N/A | 107,016 |
| **12** | Not reached | 100 | 29,279 | 29,279 | N/A | N/A | 96,936 |
| **EPT** |  |  |  |  |  |  |  |
| **13** | 79 | 0.0 | 133 | 860 | 133 | 860 | 94,623 |
| **14** | 2E/2H | 33.8 | 7,676 | 21,074 | 546 | 13,944 | 94,905 |
| **15** | 3E/2H | 12.3 | 892 | 1,696 | 684 | 1,488 | 98,799 |
| **16** | Not reached | 100 | 14,984 | 14,984 | N/A | N/A | 118,098 |
| **17** | 2E/2H | 20.3 | 1,796 | 4,335 | 918 | 3,457 | 124,749 |
| **18** | Not reached | 100 | 10,967 | 10,967 | N/A | N/A | 97,437 |
| **19** | 2E/2H | 6.7 | 1,274 | 7,609 | 761 | 7,096 | 135,379 |
| **20** | 2E/2H | 16.9 | 1,535 | 8,754 | 53 | 7,272 | 89,609 |
| **21** | 2E/2H | 12.3 | 2,415 | 10,038 | 1,181 | 8,638 | 104,584 |
| **22** | Not reached | 100 | 15,374 | 15,374 | N/A | N/A | 114,201 |
| **23** | Not reached | 100 | 8,088 | 8,088 | N/A | N/A | 86,177 |

| C) Easy level – 4 blocks first stage |  |  |  |  |  |  |  |
| --- | --- | --- | --- | --- | --- | --- | --- |
| **Subject** | **No of Easy/Hard blocks when 80% Reached** | **% overlap** | **No of Voxels Classified Active at 80%** | **No of Voxels Classified Active at Final Scan** | **Voxels in Scan at Early Stopping but not Final Scan** | **Voxels in Final Scan but not Scan at Early Stopping** | **No of Voxels in ROI (full brain)** |
| **Control** |  |  |  |  |  |  |  |
| **1** | Not reached | 100 | 3,556 | 3,556 | N/A | N/A | 115,062 |
| **2** | Not reached | 100 | 12,803 | 12,803 | N/A | N/A | 113,564 |
| **3** | 5E/4H | 77.6 | 10,921 | 13,472 | 471 | 3,022 | 77,359 |
| **4** | Not reached | 100 | 13,680 | 13,680 | N/A | N/A | 103,591 |
| **5** | 5E/5H | 69.4 | 7482 | 10,540 | 170 | 3228 | 114,260 |
| **6** | 5E/5H | 68.5 | 3,806 | 4,674 | 603 | 1,471 | 121,353 |
| **7** | 5E/4H | 50.9 | 2,195 | 4,115 | 99 | 2,019 | 107,406 |
| **8** | 5E/5H | 81.3 | 7,068 | 7,124 | 1,278 | 1,334 | 106,267 |
| **9** | 4E/4H | 42.2 | 4,065 | 9,329 | 127 | 5,391 | 121,195 |
| **10** | Not reached | 100 | 7,340 | 7,340 | N/A | N/A | 96,565 |
| **11** | 5E/5H | 47.9 | 3,088 | 6,357 | 40 | 3,309 | 107,016 |
| **12** | Not reached | 100 | 12,429 | 12,429 | N/A | N/A | 96,936 |
| **EPT** |  |  |  |  |  |  |  |
| **13** | 4E/4H | 95.2 | 139 | 49 | 136 | 46 | 94,623 |
| **14** | 6E/6H | 33.5 | 15,556 | 15,866 | 445 | 755 | 94,905 |
| **15** | 4E/4H | 100 | 899 | 750 | 648 | 499 | 98,799 |
| **16** | Not reached | 77.7 | 13,484 | 13,484 | N/A | N/A | 118,098 |
| **17** | 6E/6H | 31.1 | 3,908 | 3,487 | 1,199 | 778 | 124,749 |
| **18** | 4E/4H | 100 | 1,302 | 3,925 | 80 | 2,703 | 97,437 |
| **19** | Not reached | 14.3 | 7,402 | 7,402 | N/A | N/A | 135,379 |
| **20** | 4E/4H | 100 | 1,302 | 8,093 | 148 | 2,898 | 89,609 |
| **21** | Not reached | 100 | 10,039 | 10,039 | N/A | N/A | 104,584 |
| **22** | Not reached | 100 | 13,817 | 13,817 | N/A | N/A | 114,201 |
| **23** | Not reached | 100 | 6,715 | 6,715 | N/A | N/A | 86,177 |

D) Hard level – 4 blocks first stage
| Subject | Scan when<br>80% Reached | %<br>overlap | No of<br>Voxels<br>Classified<br>Active at<br>80% | No of<br>Voxels<br>Classified<br>Active at<br>Final Scan | Voxels in<br>Scan at<br>Early<br>Stopping<br>but not<br>Final Scan | Voxels in<br>Final Scan<br>but not<br>Scan at<br>Early<br>Stopping | No of<br>Voxels in<br>ROI (full<br>brain) |
| --- | --- | --- | --- | --- | --- | --- | --- |
| **Control** |  |  |  |  |  |  |  |
| **1** | Not reached | 100 | 23,351 | 23,351 | N/A | N/A | 115,062 |
| **2** | Not reached | 100 | 20,749 | 20,749 | N/A | N/A | 113,564 |
| **3** | 6E/5H | 87.4 | 13,160 | 14,976 | 67 | 1,883 | 77,359 |
| **4** | Not reached | 100 | 9,874 | 9,874 | N/A | N/A | 103,591 |
| **5** | Not reached | 100 | 14,939 | 14,939 | N/A | N/A | 114,260 |
| **6** | 5E/4H | 71.6 | 6,129 | 7,829 | 527 | 2,227 | 121,353 |
| **7** | 4E/4H | 40.6 | 1,214 | 2,494 | 202 | 1,482 | 107,406 |
| **8** | Not reached | 100 | 14,634 | 14,634 | N/A | N/A | 106,267 |
| **9** | 4E/4H | 44.6 | 4,313 | 8,898 | 343 | 4,928 | 121,195 |
| **10** | 4E/4H | 40.8 | 5,360 | 10,921 | 908 | 6,469 | 96,565 |
| **11** | Not reached | 100 | 11,355 | 11,355 | N/A | N/A | 107,016 |
| **12** | Not reached | 100 | 29,279 | 29,279 | N/A | N/A | 96,936 |
| **EPT** |  |  |  |  |  |  |  |
| **13** | 4E/4H | 45.0 | 1,919 | 860 | 1,532 | 473 | 94,623 |
| **14** | 4E/4H | 84.8 | 20,638 | 21,074 | 2,774 | 3,210 | 94,905 |
| **15** | Not reached | 100 | 1,696 | 1,696 | N/A | N/A | 98,799 |
| **16** | Not reached | 100 | 14,984 | 14,984 | N/A | N/A | 118,098 |
| **17** | 6E/6H | 78.0 | 4,482 | 4,335 | 1,101 | 954 | 124,749 |
| **18** | Not reached | 100 | 10,967 | 10,967 | N/A | N/A | 97,437 |
| **19** | 4E/4H | 26.9 | 3,397 | 7,609 | 1,353 | 5,565 | 135,379 |
| **20** | 4E/4H | 77.6 | 8,107 | 8,754 | 1,312 | 1,959 | 89,609 |
| **21** | Not reached | 100 | 10,038 | 10,038 | N/A | N/A | 104,584 |
| **22** | Not reached | 100 | 15,374 | 15,374 | N/A | N/A | 114,201 |
| **23** | Not reached | 100 | 8,088 | 8,088 | N/A | N/A | 86,177 |

**Table 3:** Comparison of stopping times using α_E_ = 0.001 and α_E_ = 0.0001. Based on 80% of voxels being classified. Both 2-block and 4-block first stage conditions are presented. A) Easy and hard level: 2-block, B) Easy and hard level: 4-block.

**A) 2-block first stage**
| Easy | Hard |
| --- | --- |

| **Subject** | $\alpha_E = 0.001, \beta_E = 0.1,$ | $\alpha_E = 0.0001, \beta_E = 0.1,$ | $\alpha_E = 0.001, \beta_E = 0.1,$ | $\alpha_E = 0.0001, \beta_E = 0.1,$ |
| --- | --- | --- | --- | --- |
| **Control** |  |  |  |  |
| **1** | Not reached | Not reached | Not reached | Not reached |
| **2** | Not reached | Not reached | Not reached | Not reached |
| **3** | 104 | 215 | 79 | 79 |
| **4** | Not reached | Not reached | 89 | 99 |
| **5** | 178 | Not reached | 79 | 79 |
| **6** | 79 | 79 | 166 | 196 |
| **7** | 112 | 138 | 95 | 95 |
| **8** | 177 | 181 | Not reached | Not reached |
| **9** | 79 | 79 | 86 | 86 |
| **10** | Not reached | Not reached | 98 | 101 |
| **11** | 89 | 89 | Not reached | Not reached |
| **12** | Not reached | Not reached | Not reached | Not reached |
| **EPT** |  |  |  |  |
| **13** | 79 | 79 | 79 | 79 |
| **14** | 230 | Not reached | 79 | Not reached |
| **15** | 154 | 154 | 87 | 87 |
| **16** | Not reached | Not reached | Not reached | Not reached |
| **17** | 79 | 79 | 79 | 79 |
| **18** | 79 | 79 | Not reached | Not reached |
| **19** | Not reached | Not reached | 79 | 79 |
| **20** | 79 | 79 | 79 | 79 |
| **21** | Not reached | Not reached | 79 | 80 |
| **22** | Not reached | Not reached | Not reached | Not reached |
| **23** | Not reached | Not reached | Not reached | Not reached |

| **B) 4-block first stage** |  |  |  |  |
| --- | --- | --- | --- | --- |
| **Subject** | **Easy**<br>$\alpha_E = 0.001 \beta_E = 0.01$ | $\alpha_E = 0.0001, \beta_E = 0.1,$ | **Hard**<br>$\alpha_E = 0.001 \beta_E = 0.01$ | $\alpha_E = 0.0001, \beta_E = 0.1,$ |
| **Control** |  |  |  |  |
| **1** | Not reached | Not reached | Not reached | Not reached |
| **2** | Not reached | Not reached | Not reached | Not reached |
| **3** | 171 | 215 | 200 | Not reached |
| **4** | Not reached | Not reached | Not reached | Not reached |
| **5** | 178 | Not reached | Not reached | Not reached |
| **6** | 180 | 202 | 166 | 196 |
| **7** | 164 | 164 | 155 | 155 |
| **8** | 177 | 181 | Not reached | Not reached |
| **9** | 155 | 155 | 155 | 155 |
| **10** | Not reached | Not reached | 155 | Not reached |
| **11** | 186 | 197 | Not reached | Not reached |
| **12** | Not reached | Not reached | Not reached | Not reached |
| **EPT** |  |  |  |  |
| **13** | 155 | 155 | 155 | 155 |
| **14** | 230 | Not reached | 155 | Not reached |
| **15** | 155 | 155 | Not reached | Not reached |
| **16** | Not reached | Not reached | Not reached | Not reached |
| **17** | 216 | 216 | 218 | 224 |
| **18** | 155 | 155 | Not reached | Not reached |
| **19** | Not reached | Not reached | 155 | 155 |
| **20** | 155 | 155 | 155 | 160 |
| **21** | Not reached | Not reached | Not reached | Not reached |
| **22** | Not reached | Not reached | Not reached | Not reached |
| **23** | Not reached | Not reached | Not reached | Not reached |

**Figure 3:**
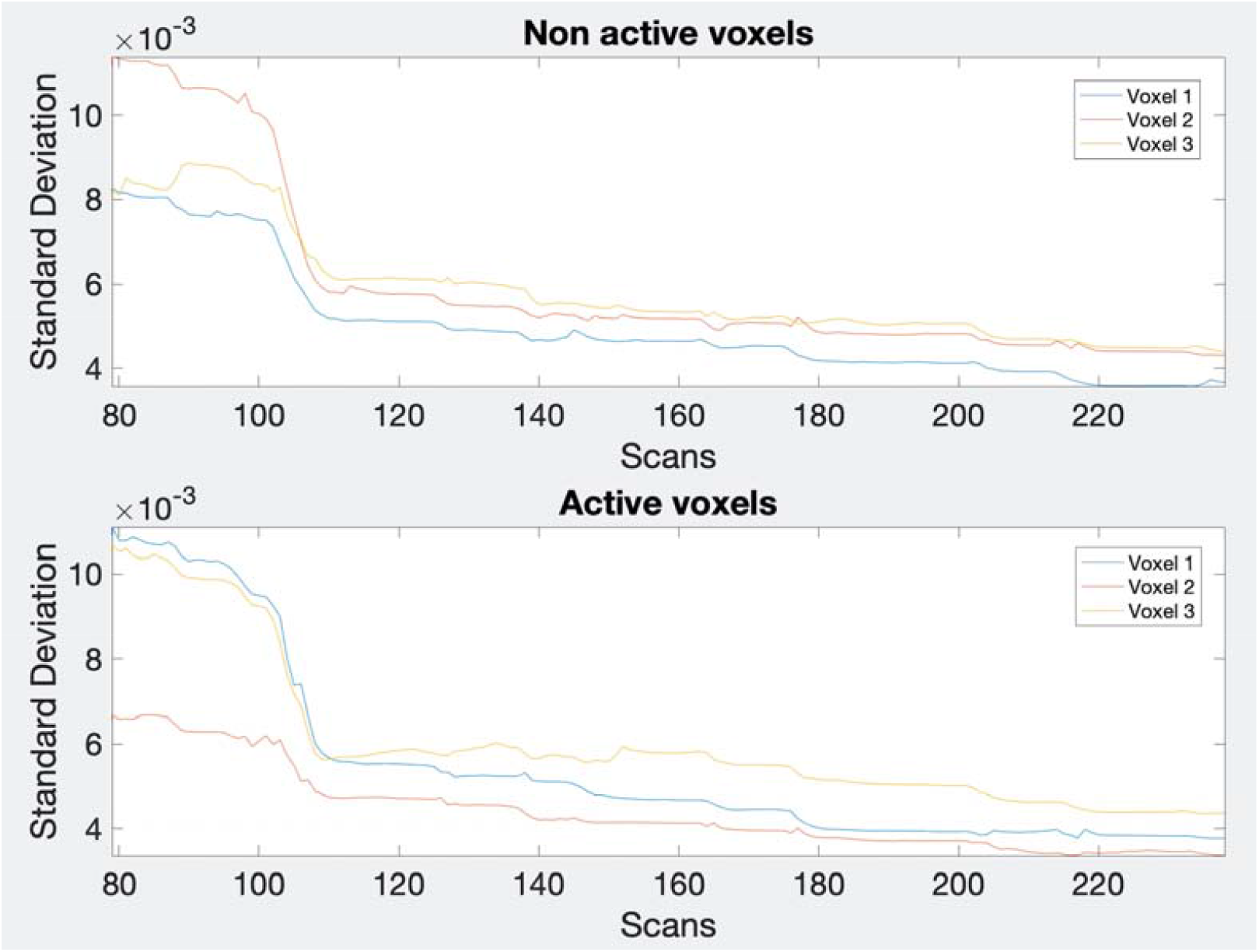
Estimated standard deviations for 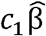. Plots for 3 sample active (bottom) and non-active (top) voxels from a control subject (subject 3) showing how the estimates decrease over time (scan number).

Stopping was reached at 80% of voxels classified as either active or non-active in around 54% of cases in both scenarios for both difficulty levels. At 80% classification for control subjects, 7/12 subjects stopped early for the easy level with both the 2-block and 4-block first stages. For the hard level, 7/12 subjects with 2-blocks and 5/12 with 4-blocks stopped early. For EPT subjects, 6/11 subjects stopped early for the easy level for both 2- and 4-block first stage conditions. For the hard level, 5/11 subjects using 4-blocks as a minimum still stopped early and 7/12 subjects using 2-blocks as a minimum stopped early. The median stopping duration for both difficulty levels for control subjects was 3 blocks of easy and 2 blocks of hard stimulus administration for 2-blocks first stage. For 4-blocks first stage, the median stopping time was 5 easy, 4 hard for the easy level and 4 easy, 4 hard for the hard level. In EPT subjects, the median stopping time for 2-block first stage was 2 easy and 2 hard blocks of stimuli. For 4-blocks first stage, the median stopping time was 4 easy and 4 hard for both difficulty levels. Depending on the number of first stage blocks, time savings of 1/3 to 2/3 (4 to 8 minutes on a 12 minute scan) can be achieved.

An early stopping rule based on a classification rate of at least 70% or 90% was also tested. Results reported in Table 4. At 70% classification most subjects stopped early. For the 2-block first stage - easy level, only 3 out of 23 subjects did not stop early and for the hard level, 1 subject did not stop early. Median stopping scan number was 79 for both the easy and hard levels. For the 4-block first stage – easy level, 5/23 subjects did not stop early and 4/23 subjects did not stop early for the hard level. Median stopping scan was 155 for both difficulty levels. At 90% classification, there were very few instances when early stopping occurred. For the 2-blocks first stage condition, 3 subjects stopped early for the easy level and 1 subject for the hard level. Only 1 subject stopped early under the 4-blocks first stage condition for the easy level. A visual comparison of the activation maps for 70% and 80% voxel classification (see Table S1 in Supplemental Information) shows that in many instances there is little difference between the two stopping points. When analysing counts of voxels classified as active or non-active between these rules, the 80% thresholds lead to more non-active classifications, but the difference in active voxels is less systematic. Given that early stopping occurs almost invariably with the 70% rule, this criterion should also be considered. Table S2 provides plots of the percentage of voxels that are respectively classified as active and non-active over the course of the full scanning duration. A general trend is that the percentage of non-active voxels gradually decreases while that of active voxels increases. Longer scan durations also may allow for some adjustment of z-score activation thresholds in post-hoc analyses, and may have some potential advantages for group analysis, as discussed below. Hence, we present results for the more conservative 80% rule, which leads to relatively longer durations even when early stopping occurs.

**Table 4:** A comparison of the early stopping times at 70%, 80% and 90% of voxels classified as either active or non-active. Conducted using α_E_ = 0.001, β_E_ = 0.1. Both 2-block and 4-block first stage conditions are presented. A) Easy and hard level: 2-block, B) Easy and hard level: 4-block.

| Subject | Easy |  |  | Hard |  |  |
| --- | --- | --- | --- | --- | --- | --- |
|  | Scan when<br>70% reached | Scan when<br>80% reached | Scan when<br>90% reached | Scan when<br>70% reached | Scan when<br>80% Reached | Scan when<br>90% reached |
| **Control** |  |  |  |  |  |  |
| **1** | 147 | Not reached | Not reached | 79 | Not reached | Not reached |
| **2** | 79 | Not reached | Not reached | 80 | Not reached | Not reached |
| **3** | 89 | 104 | Not reached | 79 | 79 | Not reached |
| **4** | 144 | Not reached | Not reached | 79 | 89 | Not reached |
| **5** | 79 | 132 | Not reached | 79 | 79 | Not reached |
| **6** | 79 | 79 | 79 | 86 | 166 | Not reached |
| **7** | 79 | 112 | Not reached | 80 | 95 | Not reached |
| **8** | 140 | 177 | Not reached | 98 | Not reached | Not reached |
| **9** | 79 | 79 | 79 | 79 | 86 | Not reached |
| **10** | 79 | Not reached | Not reached | 79 | 98 | Not reached |
| **11** | 79 | 89 | Not reached | Not reached | Not reached | Not reached |
| **12** | 79 | Not reached | Not reached | 79 | Not reached | Not reached |
| **EPT** |  |  |  |  |  |  |
| **13** | 79 | 79 | 111 | 79 | 79 | 79 |
| **14** | 84 | 230 | Not reached | 79 | 79 | Not reached |
| **15** | 107 | 154 | Not reached | 79 | 87 | Not reached |
| **16** | Not reached | Not reached | Not reached | 109 | Not reached | Not reached |
| **17** | 79 | 79 | Not reached | 79 | 79 | Not reached |
| **18** | 79 | 79 | Not reached | 197 | Not reached | Not reached |
| **19** | 79 | Not reached | Not reached | 79 | 79 | Not reached |
| **20** | 79 | 79 | Not reached | 79 | 79 | Not reached |
| **21** | Not reached | Not reached | Not reached | 79 | 79 | Not reached |
| **22** | Not reached | Not reached | Not reached | 79 | Not reached | Not reached |
| **23** | 203 | Not reached | Not reached | 79 | Not reached | Not reached |

B) 4-block first stage
| **Subject** | **Easy** |  |  | **Hard** |  |  |
| --- | --- | --- | --- | --- | --- | --- |
|  | **Scan when**<br>**70% reached** | **Scan when**<br>**80% reached** | **Scan when**<br>**90% reached** | **Scan when**<br>**70% reached** | **Scan when**<br>**80% Reached** | **Scan when**<br>**90% reached** |
| **Control** |  |  |  |  |  |  |
| **1** | 155 | Not reached | Not reached | 161 | Not reached | Not reached |
| **2** | Not reached | Not reached | Not reached | 155 | Not reached | Not reached |
| **3** | 155 | 171 | Not reached | 155 | 200 | Not reached |
| **4** | 155 | Not reached | Not reached | 155 | Not reached | Not reached |
| **5** | 155 | 177 | Not reached | 155 | Not reached | Not reached |
| **6** | 155 | 180 | Not reached | 158 | 166 | Not reached |
| **7** | 155 | 164 | Not reached | 155 | 155 | Not reached |
| **8** | 155 | 177 | Not reached | 155 | Not reached | Not reached |
| **9** | 155 | 155 | Not reached | 155 | Not reached | Not reached |
| **10** | 155 | Not reached | Not reached | 155 | 155 | Not reached |
| **11** | 155 | 186 | Not reached | Not reached | Not reached | Not reached |
| **12** | Not reached | Not reached | Not reached | Not reached | Not reached | Not reached |
| **EPT** |  |  |  |  |  |  |
| **13** | 155 | 155 | 178 | 155 | 158 | Not reached |
| **14** | 155 | 230 | Not reached | 155 | Not reached | Not reached |
| **15** | 155 | 155 | Not reached | 155 | Not reached | Not reached |
| **16** | Not reached | Not reached | Not reached | 155 | Not reached | Not reached |
| **17** | 155 | 216 | Not reached | 155 | 218 | Not reached |
| **18** | 155 | 155 | Not reached | 197 | Not reached | Not reached |
| **19** | 155 | Not reached | Not reached | 155 | 155 | Not reached |
| **20** | 155 | 155 | Not reached | 155 | 159 | Not reached |
| **21** | Not reached | Not reached | Not reached | Not reached | Not reached | Not reached |
| **22** | Not reached | Not reached | Not reached | Not reached | Not reached | Not reached |
| **23** | 203 | Not reached | Not reached | 155 | Not reached | Not reached |

The activation maps under the different conditions are shown in Figure 4 for a sample subject (subject 9). For the 2-block first stage, this subject terminated after 2 blocks of easy and hard administration for the easy level (scan 79) and after 3 blocks of easy and 2 blocks of hard administration for the hard level (scan 97). For the 4-block minimum, this subject terminated at scan 155, equal to 4 blocks of easy and hard stimulus administration, for both difficulty levels. The images show that at scan 79 there is very little activity present and the majority of the voxel classifications are non-active. By scan 155, there is much more activity which has a similar pattern to the final scan. The extent is not quite as large as the final scan, however the foci of the clusters do overlap between the two time points. As mentioned, this is likely due to the alternative hypothesis threshold *c*′*β* value corresponding to relatively lower z-score values as the number of scans increase. This pattern of ‘growing’ activations for given alternative hypothesis *c*′*β* over scan duration is thus typical of our early stopping data, particularly for the 2-block initial stage. Visual inspection of the z-score maps at the stopping scan for other subjects revealed similar patterns. In most instances, the additional active voxels at full duration were around the edges of existing clusters at the early stopping scans. Further images of other subjects are presented in Table S1 in the Supplementary Information document. Plots of the percentage of active and non-active voxels classified at each scan are given in Table S2 of the same document. The overlaps between early stopping and full duration maps are also explored further in Table 2 where we show the number of active voxels in common spatially between the two durations. Although some of these show less than 50% overlap with the final scan, it can be seen that this is due to the smaller cluster sizes with early stopping. The median spatial overlap where early stopping occurs for control subjects was 27.9% (SD 30.2%) for the easy level, 2-blocks and 68.5% (SD 15.4%) for the easy level, 4-blocks. For the hard level, there was 26.0% (SD 19.9%) and 44.6% (SD 21.3%) for the 2-block and 4-block first stages, respectively. For EPT subjects the median overlap was 34.2% (SD 34.0%) and 33.5% (SD 34.3%) for the easy level, 2- and 4-block first stages, respectively. For the hard level, the median overlap values were 14.6% (SD 9.4%) and 77.6% (SD 25.2%) for 2-block and 4-block first stages.

**Figure 4:**
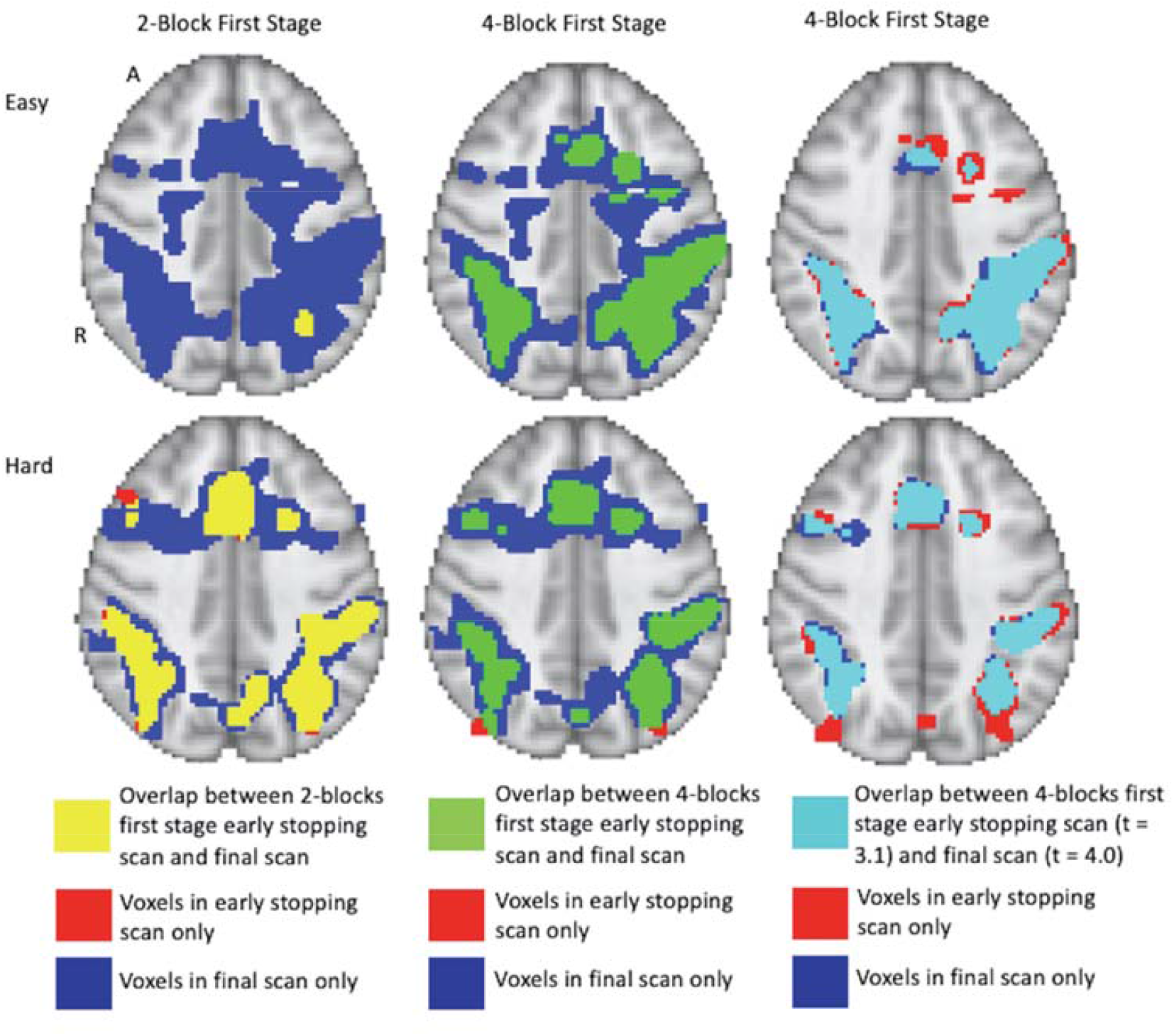
Full brain activation maps showing the overlapping voxels between the different stopping points (using 2-blocks first stage, 4-blocks first stage and final scan). Top row shows the easy level and bottom row shows the hard level for 1 subject (number 9). The active voxels that are active only at full duration are shown in blue. Those only active after 2-blocks or 4-blocks of stimulus administration are in red. Yellow shows the overlap between full duration and 2-block first stage early stopping scans. Green shows the overlap between full duration and 4-block first stage stopping scans. P ≤ 0.001 uncorrected, z > 3.1. Right hand images show the comparison of 4-blocks early stopping with z > 3.1 with full duration that has been thresholded at z > 4.0. Light blue indicates overlapping voxels. Results overlaid on MNI template, slice z = 56 shown. R = right, A = anterior.

This phenomenon is basically driven by the estimation variance of the GLM parameters steadily decreasing as more scans are accrued, while at the same time the alternative hypothesis z-score threshold is being held the same. Given that estimated beta and error variance values essentially become stable in most cases, voxel-level z-scores will increase. This leads to increasingly larger number of voxels being classified as active. We assessed a sample of the error variance estimates over scan duration, as in Figure 3. We illustrate similarities in activation patterns with early stopping and full duration if the z-score threshold increases as the scan durations increase. Assuming no serial correlation as an approximation, note that the variance of 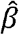 is (*σ*)^2^ (*X′X*) ^−1^ where X is the known design matrix. (*X′X*)^−1^; is thus known as well for all scans, and it decreases as scan duration increases. Stopping early at a given threshold can thus be similar to stopping later with a stricter threshold for activation, provided error variance and beta parameter estimates stabilize, which we assess for with the first stage. A z-score of 3.1 for 154 scans approximately corresponds to a z-score of 4.0 for the final scan (228 scans). At 78 scans, a z-score of 3.1 corresponds to a z-score of approximately 8.37 and 5.92 for the easy and hard task parameter respectively, so there will be even less overlap, even if the z-score threshold is 4.0 at full duration. See Table S1 in the Supplement for images resulting from different stop rules and first stage durations. In Table S2, the trends in percentage of voxels classified as active and non-active reflect this phenomenon, at least for some of the subjects.

Importantly, the issue of whether early stopping or full duration provide better activation maps is best answered neuroscientifically, through the support of literature and hypotheses. In the Supplement, we add plots for early stopping versus full duration for each subject for which early stopping was invoked. Comparing these plots, we see that in many instances that the cluster peaks are located in the expected anatomical locations. However, at full duration results, numerous voxels with lower z-scores appear around the edges of the clusters and extend well beyond the anatomical boundaries of the gyri indicating areas of activation in white matter and cerebrospinal fluid. This suggests that these lower z-score voxels are more likely to be false positives as scan duration increases, as argued above, and indicates that scanning for full duration doesn’t necessarily improve the results.

In summary, although there are similar rates of early termination between the 2-block and 4-block first stage cases, the detected activation patterns suggest that using 4-blocks of stimulus administration is more suited to determining active voxels. In Figure 3, to illustrate the rate of decrease of the estimated 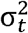 values, we present a plot of 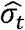 values for one subject across a set of voxels over the duration of the experiment. These values give an indication that stopping based on the *θ*-values at the end of the 2-block first stage may be too early for correspondence with full duration scans, as the estimated standard deviations are relatively larger. This implies that the alternative hypotheses can have much larger *θ*-values early on compared to full duration, while this difference in *θ*-values is less after 4-blocks. Note that in some subjects there is some volatility due to subject movement. Below, we consider full duration z-score thresholds of 4.0 versus early stopping results with a threshold of 3.1, so that activation magnitudes considered as active are more comparable. See also, Table 5 comparing the overlap in cluster locations and extent from full duration z = 4.0 with early stop scan z = 3.1.

**Table 5:**
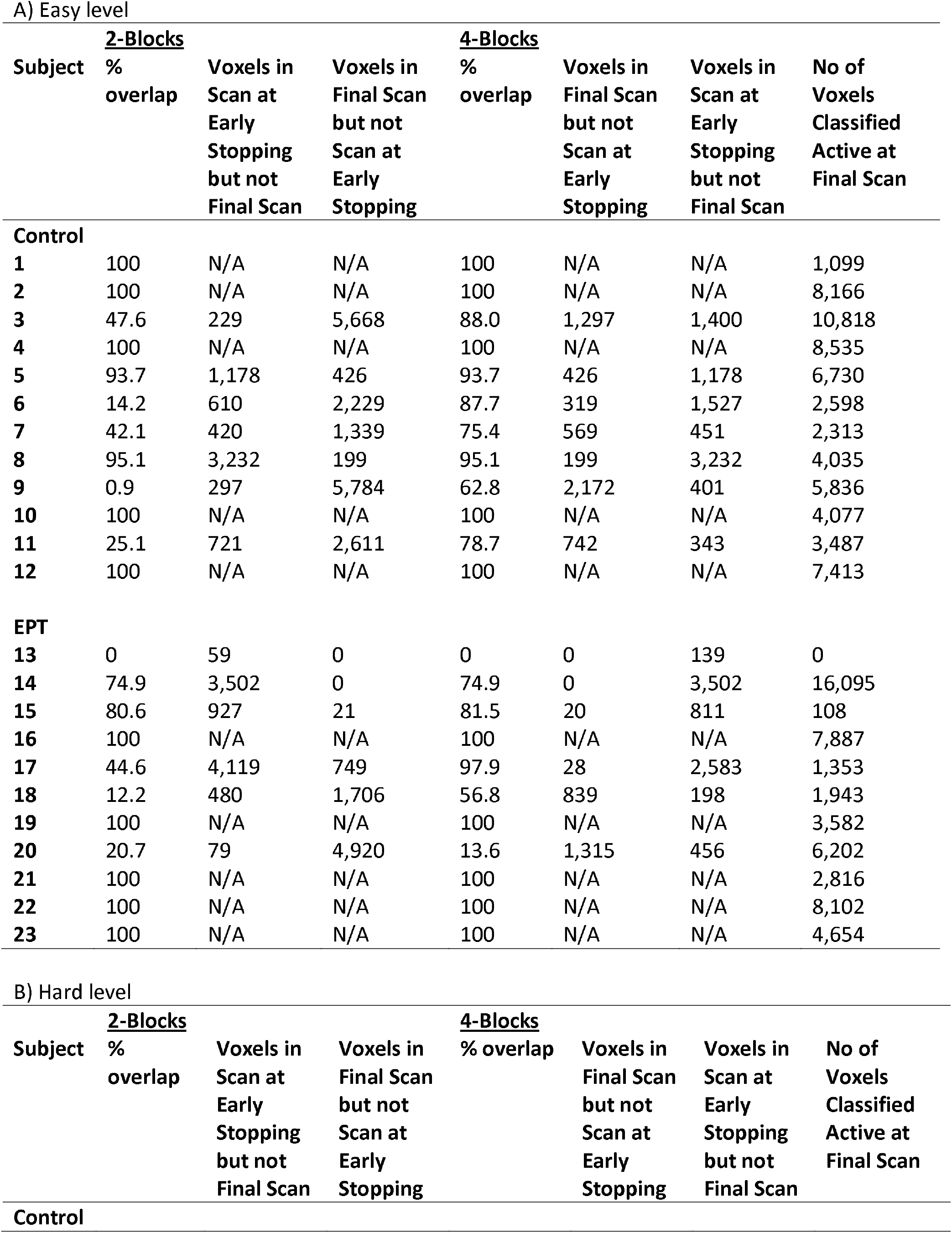
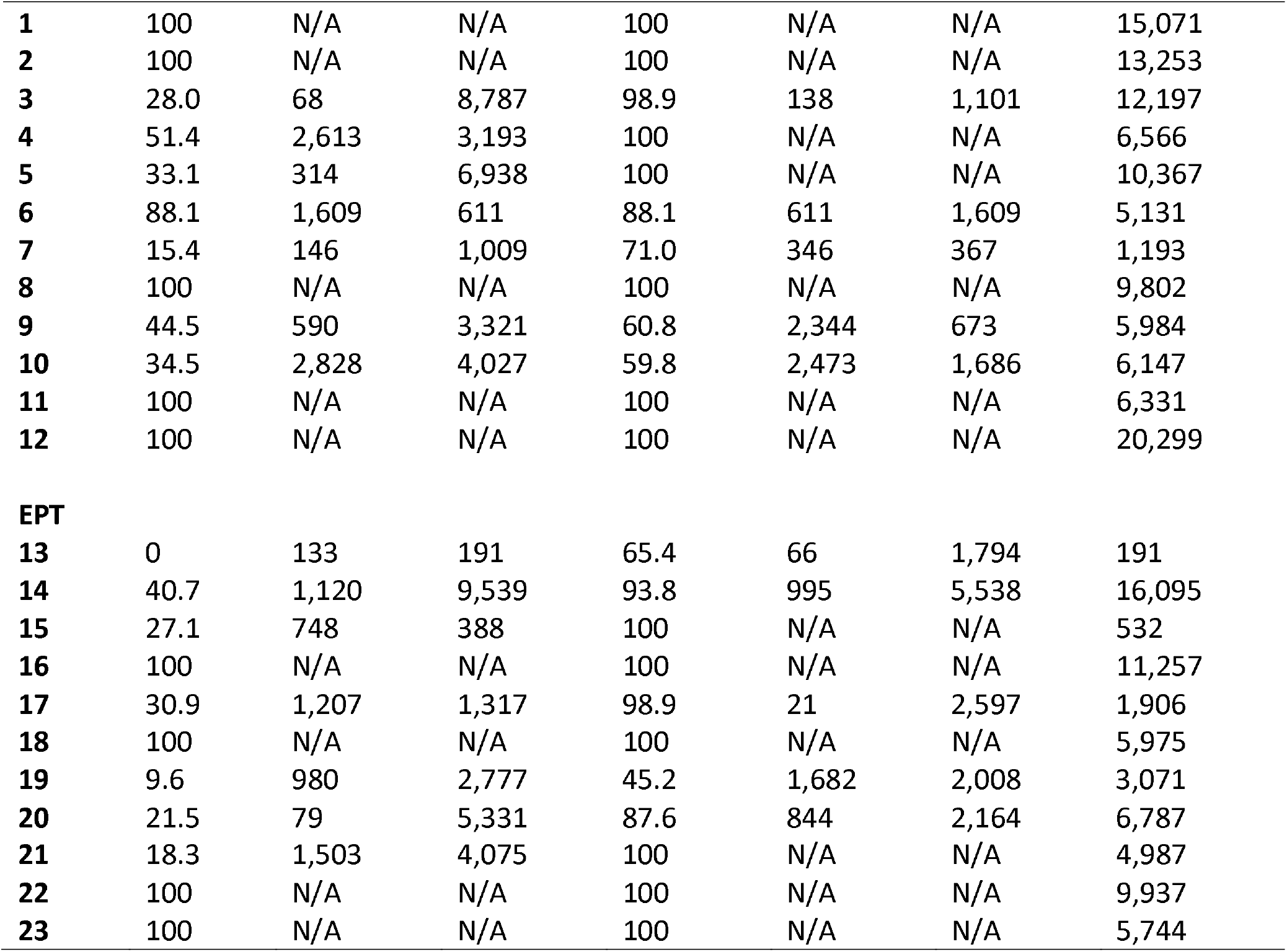
Overlap with full duration scan threshold of z = 4.0. The 2- and 4-block first stage results are thresholded at z = 3.1. The percentage of voxels-in-common is also given relative to the total number of active voxels at the full duration scan. Median values are calculated with those who stopped early only. N/A = not applicable.

In all but three EPT subjects the active voxel count increases with scan duration. Subjects 13, 15 and 17 are the exception. Subject 13 demonstrates very few active voxels at all and there is almost no consistency in location. Further investigation shows large relative framewise displacement occurs frequently throughout the scan and many of the responses have been missed or have relatively long response times, 60.5% correct overall and 2.08 s (SD 0.97 s) average response time (see plots for subject 13 in Table S3 of Supplementary Information document). Taken together these suggest that either the task level may not have been aimed at the right level and/or the subject may have been uncomfortable and distracted in the scanner thereby attending to the task less than required for robust activations to occur. Subject 15 demonstrates cluster sizes that decrease over time. Framewise displacement shows very little motion, particularly from scan 180 onwards. The response plots (in Table S3 of Supplementary Information) show the subject is paying attention and responding appropriately. Subject 17 has a similar pattern of decreasing cluster sizes. The framewise displacement plots indicate a moderate amount of motion throughout. Although the subject has missed many of the task questions (65.8% correct), the pattern of responding indicates they are awake and attending to the task. In general, EPT subjects demonstrated more motion. The median number of scans with framewise displacement above a threshold of 0.9 mm, threshold determined from (51), was 5 scans (SD 43 scans) for EPT subjects and 2.5 scans (SD 8 scans) for control subjects. One EPT subject passed the threshold a total of 124 scans out of 238 scans. In contrast, the control subject with the maximum number of threshold passes was 30/238 scans. This is further demonstrated in Figure 5 where we show subject counts for each scan when the threshold has been passed. For both EPT and control subjects, it is clear that subjects are moving more frequently in the second half of the scans and supports stopping early to reduce motion artifacts and noise in the data. Formally, we see statistically significant differences when comparing counts of motion events with framewise displacement greater than 0.9mm in the first versus second half of scanning (p= 0.003, two-sided signed rank test). EPT group also has significantly more movement in the first half of scanning (p= 0.035, two-sided Mann-Whitney test), indicating a group-level proclivity for more motion events.

**Figure 5:**
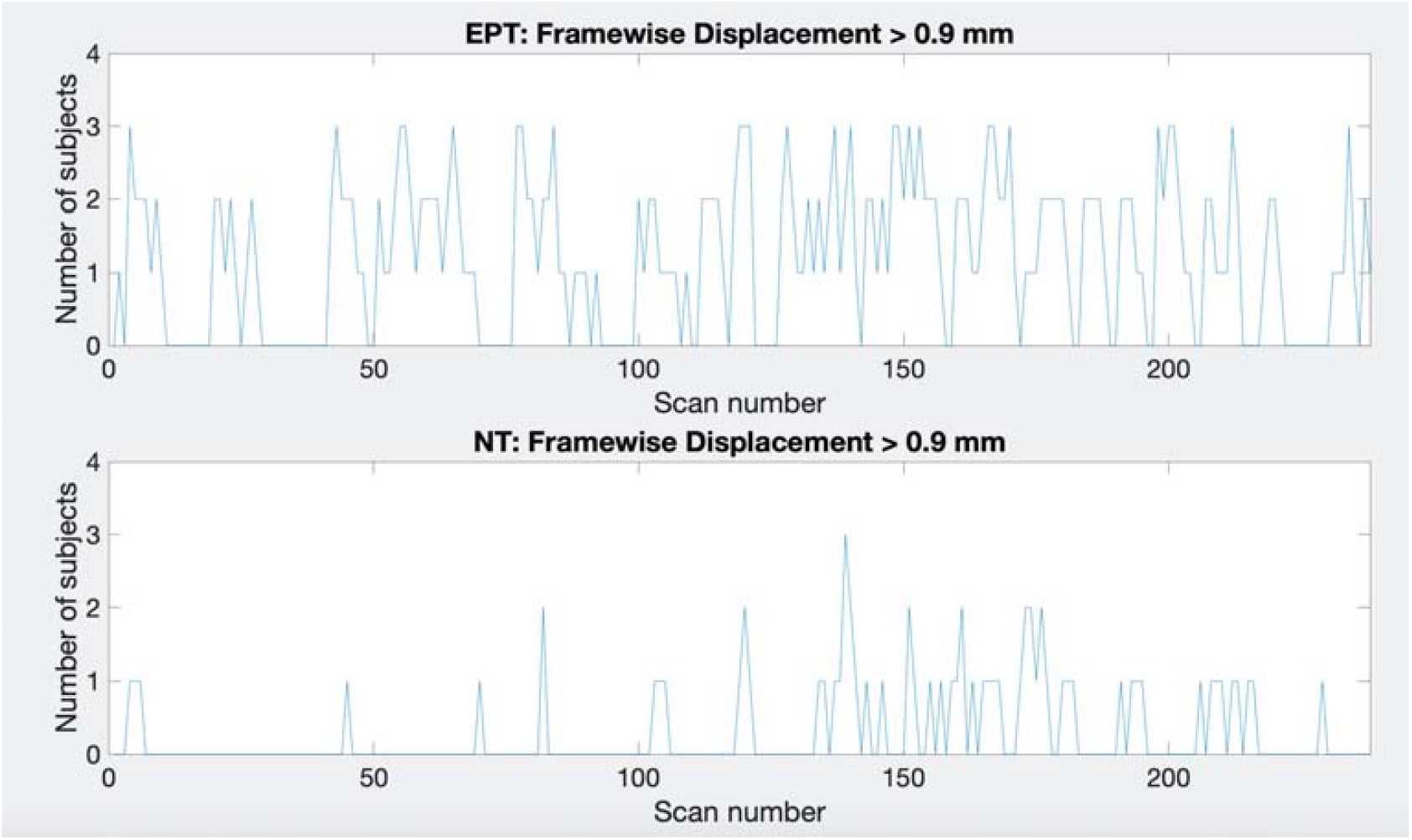
Plots showing the number of subjects that pass the framewise displacement threshold of 0.9 mm for each scan. Top: EPT subjects, bottom: control subjects.

### 4.2 Group Analysis Results

The results for the 1-back easy and hard contrasts for the 2- and 4-block first stage conditions for EPT and control subjects are shown in Figure 6. Location of activity is listed in Table S4 of the Supplementary Information. The group results of full scan durations are compared to the group results using only the scans up to the early stopping point for each subject for each difficulty level and number of blocks completed before early stopping was allowed. We examined within group differences as well as between group differences. The EPT > control and control > EPT contrasts did not show any differences with the full duration and early stopped scans, which could in part be due to sample size limitations and the within group heterogeneity of the EPT group. The focus for the results here are within group for the easy and hard levels.

**Figure 6:**
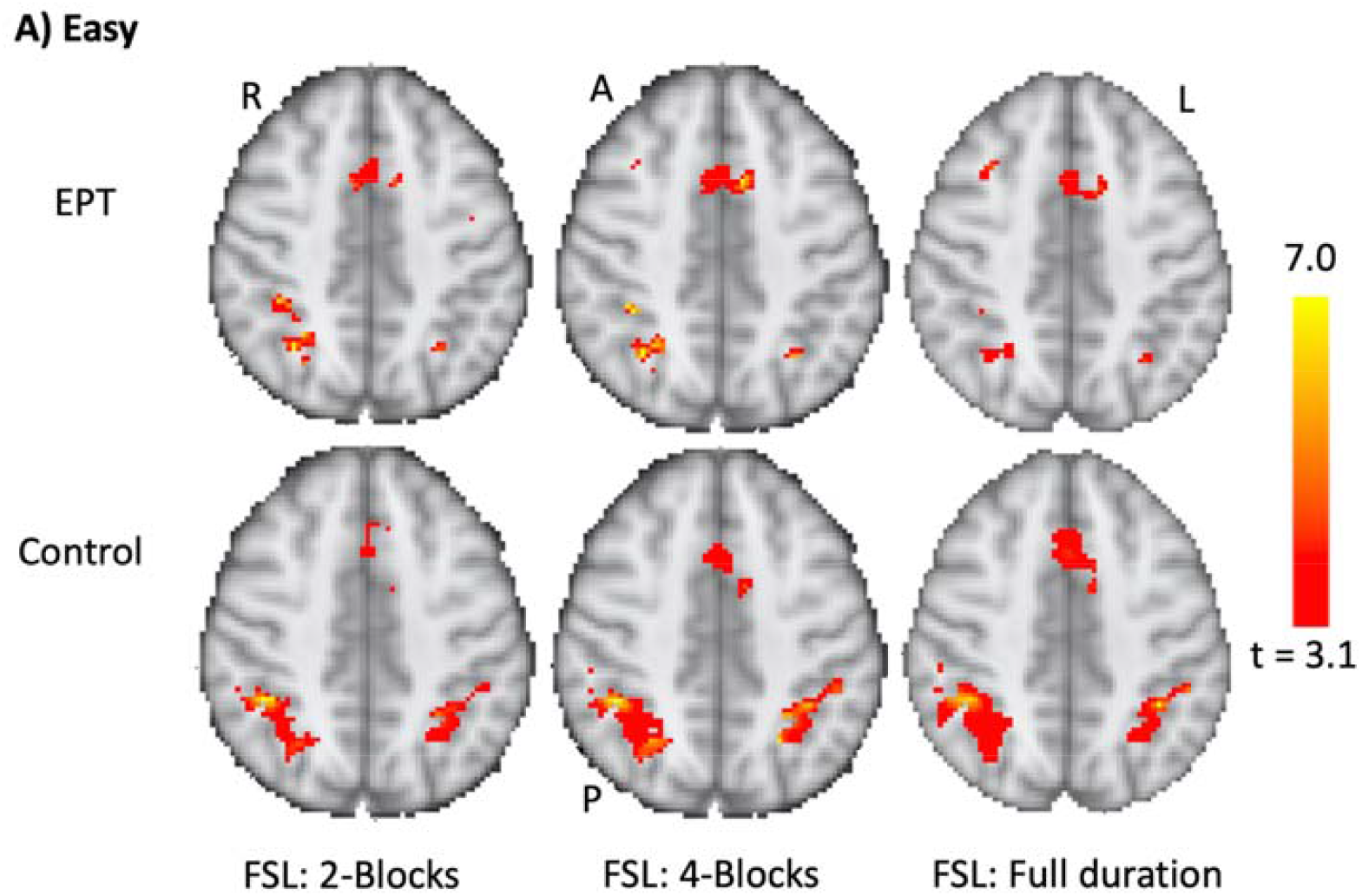
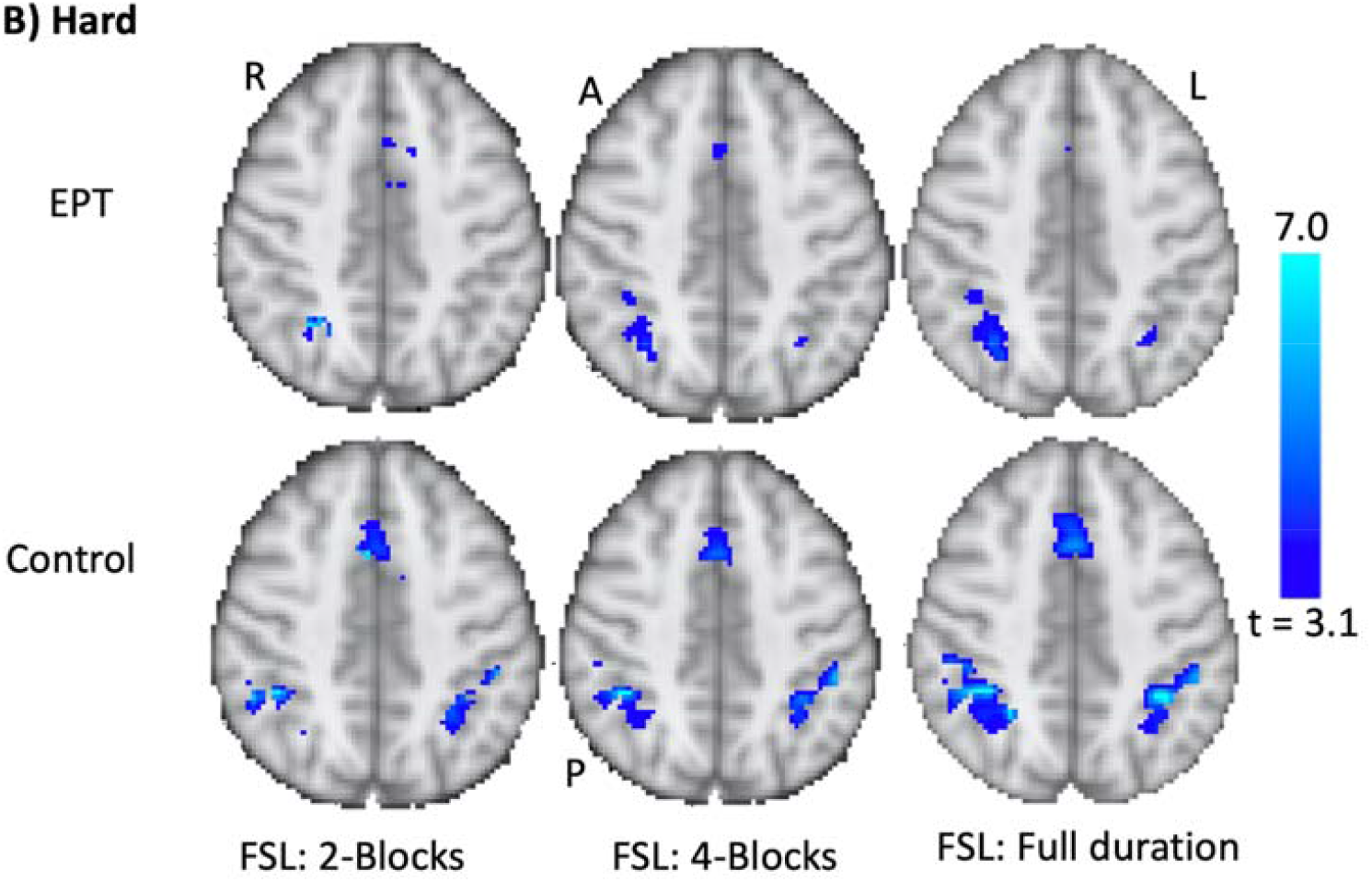
Group results for the 1-back task. Analysis performed for controls and EPT subjects using FSL. Early stopping with 2- and 4-blocks being initially administered is compared to full duration. Activations are overlaid on the MNI template brain. Red (A) = easy level results, Blue (B) hard level results. P < 0.001 uncorrected. Slices z = 58 is shown. R = right, L = left, A = anterior, P = posterior.

The control subjects show strong activations in the anterior cingulate and bilateral parietal regions, see Tables 6 and S4, and Figure 6. The easy and hard 4-block first stage scans appear similar to the final scans. There is less correspondence between the 2-block first stage scans and the final scans, reflecting the individual results reported above. The EPT group easy level scans are consistent across all stages but there is more variability in the activations across the hard level. Across all EPT scans, there is more right sided activity compared to controls. This is discussed below.

**Table 6:**
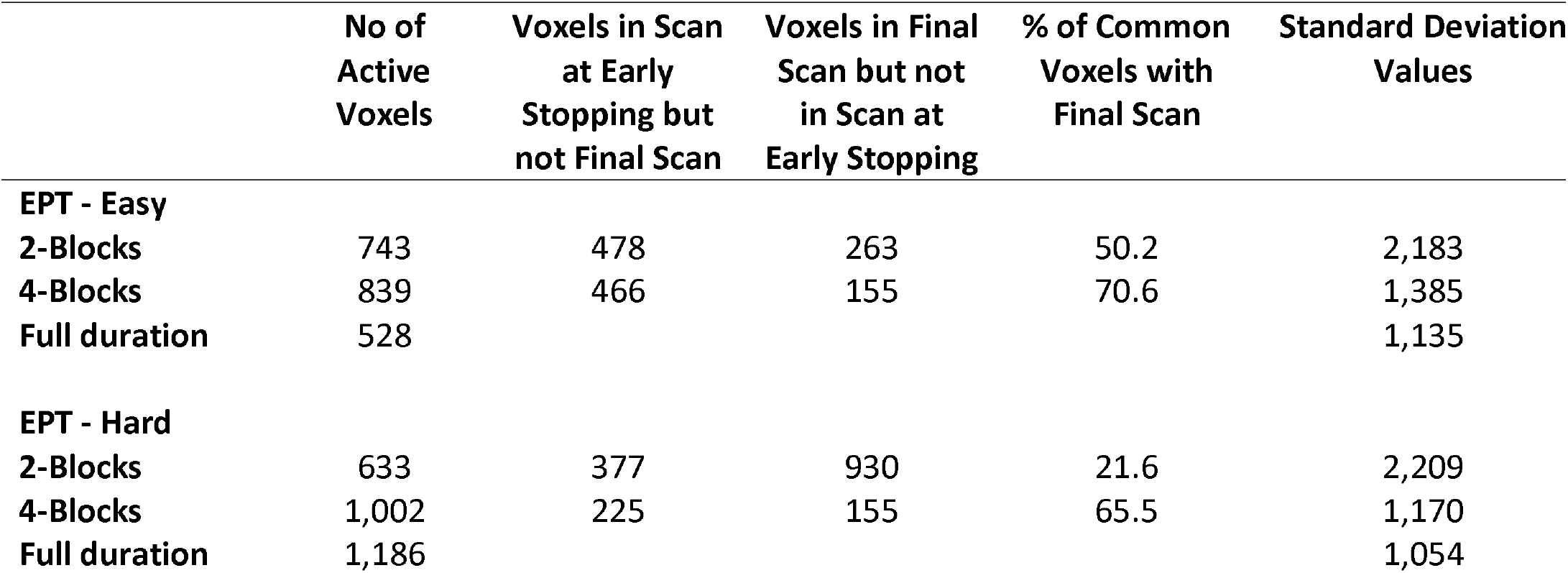
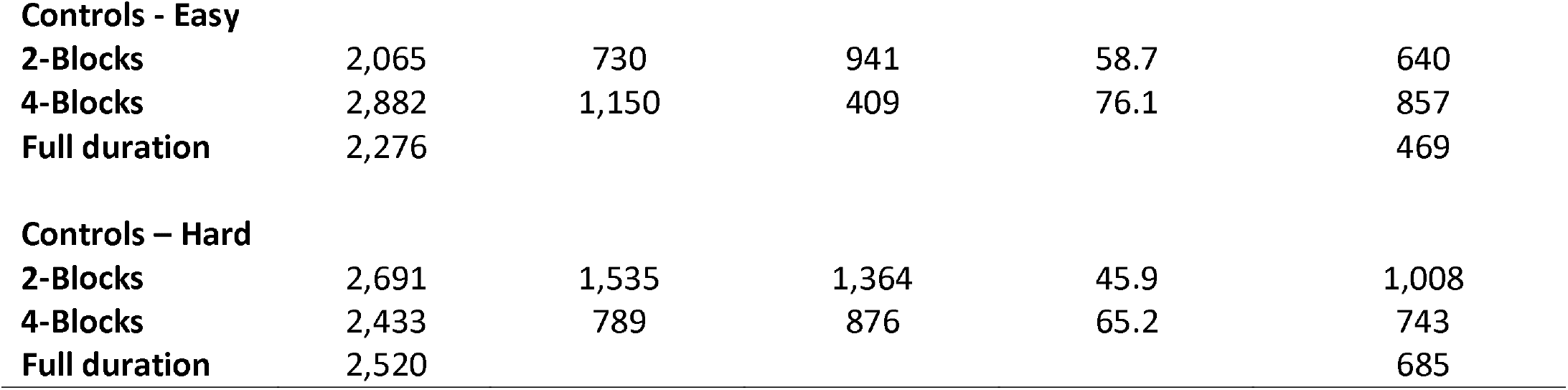
The number of active voxels that spatially overlap between early stopping and full duration group analyses are listed. Images thresholded at p < 0.001. The percentage of voxels-in-common is given relative to the total number of active voxels detected at full duration. Stopping based on 80% classification at the individual level.

|  | No of Active Voxels | Voxels in Scan at Early Stopping but not Final Scan | Voxels in Final Scan but not in Scan at Early Stopping | % of Common Voxels with Final Scan | Standard Deviation Values |
| --- | --- | --- | --- | --- | --- |
| **EPT - Easy** |  |  |  |  |  |
| **2-Blocks** | 743 | 478 | 263 | 50.2 | 2,183 |
| **4-Blocks** | 839 | 466 | 155 | 70.6 | 1,385 |
| **Full duration** | 528 |  |  |  | 1,135 |
| **EPT - Hard** |  |  |  |  |  |
| **2-Blocks** | 633 | 377 | 930 | 21.6 | 2,209 |
| **4-Blocks** | 1,002 | 225 | 155 | 65.5 | 1,170 |
| **Full duration** | 1,186 |  |  |  | 1,054 |
| **Controls - Easy** |  |  |  |  |  |
| **2-Blocks** | 2,065 | 730 | 941 | 58.7 | 640 |
| **4-Blocks** | 2,882 | 1,150 | 409 | 76.1 | 857 |
| **Full duration** | 2,276 |  |  |  | 469 |
| **Controls – Hard** |  |  |  |  |  |
| **2-Blocks** | 2,691 | 1,535 | 1,364 | 45.9 | 1,008 |
| **4-Blocks** | 2,433 | 789 | 876 | 65.2 | 743 |
| **Full duration** | 2,520 |  |  |  | 685 |

## 5.0 Discussion

Based on analysis of a training sample, we have presented a workflow for the implementation of an adaptive real-time fMRI system that allows for statistically-driven dynamic adjustment of experimentation based on voxel-level SPRT. We show that this dynamic and adaptive statistical approach is generally comparable to corresponding fixed experimental designs in terms of detected activation, particularly when adjusting for stricter z-score thresholds for full scan to account for reduced estimation variance. At the same time, time savings in experiment durations can be substantial. Moreover, with respect to individual data, as scans increased, we observed that more and more of the newly classified active voxels were located around the edges of clusters in many subjects. For some, clusters would even merge into one larger cluster across the brain that would consist of 10,000’s of voxels. This effect was addressed by the work of Saad et al. (2003) who investigated the effect of the number of time points on the extent of brain activations. They observed a similar effect that longer scanning potentially increases the detection of false positives but not the detection of true positives (52).

We explored imposing two different first stage lengths before early stopping is considered using either 2- or 4-blocks each of easy and hard stimulus administration. The 4-block first stage is justified over the 2-block because of the comparative stability of the estimation of error variances and other GLM parameters. In contrast, for the 2-block first stage, parameter estimation can be more variable. Also, correspondence in early-stop activation patterns to full scan duration requires very high z-score threshold adjustments, which may be too stringent to detect important activations. The 2-block first stage often led to most voxels being classified as non-active. See Table S2. While the 4-block first stage provides less opportunity for efficiency gains, as the window for early stopping is narrower, but it is more prudent given the need for parameter estimates to stabilize. It is possible that a 3-block initial stage could provide comparable results as the 4-block initial stage, but this was not explored here.

In the SPRT framework, other α_E_, β_E-_ pairs were considered as well, to test how different combinations impact activity detection and early stopping. For instance, given selection of α_E_ = 0.001 and β_E_ = 0.01, overall stopping did not occur. In this case, the more stringent choice of β_E_ makes it more difficult to cross either of the SPRT thresholds. We also saw that for either α_E_ = 0.001 or α_E_ = 0.0001 being paired with β_E_ = 0.1, early stopping occurred for both of the experimental conditions, with somewhat faster early stopping for the less stringent α_E_.

In terms of the global stop rule threshold, we observed that for the cases under consideration, stopping when 80% of voxels in the full brain (or smaller ROI) respectively satisfy their SPRT-based stopping criterion generally leads to early stopping of stimulus administration, while also leading to comparable activation classification as with the full protocol, after z-test score threshold adjustment for scan duration. The stricter 90% criterion was infrequently satisfied, and did not often lead to early stopping of experimentation. Recall that when GLM parameter values are “in-between” the null and alternative hypothesis values, SPRT-based stopping is less likely at the voxel level. A 100% stopping rule is thus not feasible, as are values relatively close to 100%. This phenomenon becomes less of an issue with more scans, since θ_*t*_, the alternative hypothesis threshold for *c*′*β*, decreases in value as more scans are accrued, given the z-score threshold of 3.10 is held constant. Fewer voxels are then “in-between”. The 80% rule seems conservative in that not all participants are stopped early, but there are high levels of correspondence in individual and group level activation maps with full durations, particularly when the first stage is comprised of 4 blocks, and the full scan z-score is adjusted. The 70% rule is more aggressive, and early stopping is invoked at a much higher rate. Given that the resultant images from early stopping in many cases appear similar across these two rules, the 70% rule should be considered as well.

The SPRT approach was effective at detecting brain activity at the individual level with early stopping in both the control and EPT groups. Note the individual variability among subjects in early stopping performance. Factors that can affect stopping times include the magnitudes of activation, variability in task performance, sustained attention levels, motion, and the noise levels in the BOLD signal. Those born EPT also can have structural abnormalities of the brain which can affect fMRI results and 2 subjects reported here had clear abnormalities that were obvious even in this low resolution data. Less obvious abnormalities may have been present in some of the other subjects.

The EPT group data demonstrated more right sided activity and smaller cluster sizes by comparison to control group data across all stopping points. In order to understand this result it is necessary to consider neuropsychological skills and structural and functional brain changes within the group. Working memory is a key skill required for both mathematics and this numerical 1-back task. Recall the lower accuracy and longer response times in the EPT group. fMRI studies on dyscalculia (difficulty in learning and performing mathematics) suggest that there is greater heterogeneity in activations with a more diffuse pattern being apparent (53, 54). Additionally, there is overlap in structural differences in white matter integrity, as measured from diffusion weighted imaging studies, between those born EPT and those with dyscalculia including inferior fronto-occipital fasciculus and the inferior and superior longitudinal fasciculi (55-58). These connect crucial areas associated with mathematics and working memory. A more diffuse and variable pattern of functional activity, perhaps partly due to structural differences, may confound a group analysis in this instance. More data points from individuals do seem to improve the results, perhaps allowing the variability to dampen somewhat. This is supported by the change in variance for the group between early stopping with 2- and 4-block first stages and full duration analyses, see right-hand column of Table 6. The control group variances are relatively much lower throughout, as the extremely pre-mature birth group was neurologically and cognitively more heterogeneous. If group-level analysis is a main objective, it is possible that groups could be treated differently in how early stopping is approached based on within-group heterogeneity and the need for more scan data to help overcome this. This issue needs further investigation.

With this data, a group analysis was feasible using the early stopping data in controls. A possible limitation was discovered in performing a group analysis of the EPT group, as these subjects demonstrated greater variability in location at the individual level. While it is feasible to apply our approach for patient group studies, consideration should be given to the particular patient groups of interest and the likely within group differences in brain activity when making the decision to stop early. We conjecture that larger sample sizes or stricter early stopping criteria may help overcome larger variability.

In the future, it is possible that the first stage length can be tailored at the voxel level, once it is clear error variance and other GLM parameter estimates are relatively stable, which is expected at some point due to the convergence properties of the estimators. This may facilitate earlier stopping. Alternatively, if local computational resources are limited, note that stopping can be assessed on an interval basis, and not necessarily after every scan. Although not considered here, these BOLD signal-based early stopping rules could also possibly be enriched by incorporating individual motion displacement patterns, as well as behavioural measures such as correctness rates in experimentation.

Here we demonstrated full brain analytics with parallelization using MKL Intel libraries for matrix computation with two Xeon E5-2687W 8-core processors. It is also feasible to consider only partial brain volumes where experiments demand more consideration of a particular area. Future directions for the study are to implement the SPRT and Bayesian sequential estimation methods using distributed computing approaches to increase processing speed allowing full brain real-time analyses and advance stopping rule methods in shorter scan times.

## 6.0 Conclusion

We introduce a systematic, statistically-based approach to dynamic experimentation with real-time fMRI. Saving in scan time and accurate voxel activation detection can be achieved, while redundant experimentation in block design is reduced. We investigate different aspects of how to determine early stopping rules. These analyses can be viewed as intended on a training sample to guide implementation of early stopping in future studies involving the same experiments and study populations. These methods lay a foundation for future dynamic experimentation approaches and early stopping rules with real-time fMRI, including for resting state and neural feedback. Use of high performance computing will enable the advent of more sophisticated real-time experimental designs and dynamically determined early stopping rules.

## Data Availability

The raw data supporting the conclusions of this article will be made available by the authors, without undue reservation.

## Declaration of conflicts of interest

All authors declare no conflicts of interest.

## Author contributions

SC – Study design, analysis and interpretation of data, drafting of manuscript

WC – Study design, analysis and interpretation of data, software development, drafting of manuscript

JF – Study design and software development

HF – Study design and technical support, drafting of manuscript

JSG – Software development, review of manuscript

JZ – Analysis and interpretation of data, review of manuscript

CT – Study design, analysis and interpretation of data, drafting of manuscript

## Funding

This study was supported by Phillips Healthcare and the National Science Foundation (Award number: 1561716).

## Acknowledgements

None

## Notes

### Competing Interest Statement

The authors have declared no competing interest.

### Author Declarations

UH Hospitals Cleveland Medical Center IRB

### Summary of Updates

- Methods section revised and expanded for clarity. - Results revised and updated with further analyses and explanations added. - Discussion updated to reflect results. - Supplemental file updated.

