## Supplementary information for "Early Stopping in Experimentation with Real-time Functional Magnetic Resonance Imaging Using a Modified Sequential Probability Ratio Test"

**Table S1:** Statistical maps of activation for easy and hard levels for the one back task. Maps shown are for those instances where early stopping occurred and there is < 50% overlap between the active voxels of the early stopped scans and the full duration scan, See Table 2C and D in main document. 70% and 80% of voxels classified active and non-active are shown side by side for comparison. Full duration active voxels are shown in blue for all time points for easy reference. 2-block first stage result is overlaid in red, 4-block first stage result is in green. Slice number is given in MNI space. Grey matter is highlighted in yellow.

| **Sub** | **Level** | **Slice Z** | **2-Blocks (red) 70%** | **2-Blocks (red) 80%** | **4-Blocks (green) 70%** | **4-Blocks (green) 80%** | **Full duration (blue)** |
|---|---|---|---|---|---|---|---|
| **Control** | | | | | | | |
| **3** | Easy | 52 | Scan 89<br>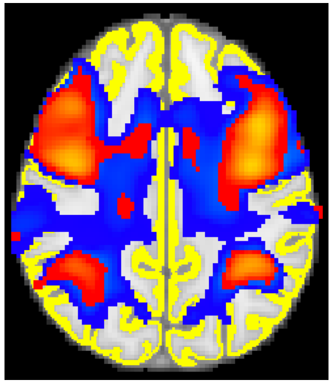 | Scan 104<br>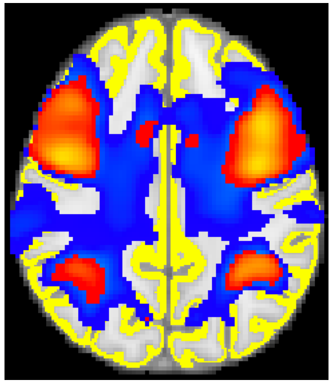 | Scan 155<br>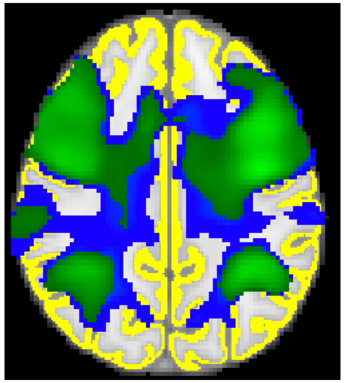 | Scan 171<br>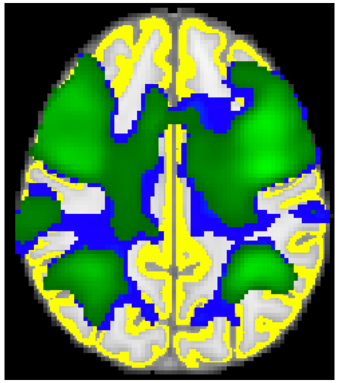 | 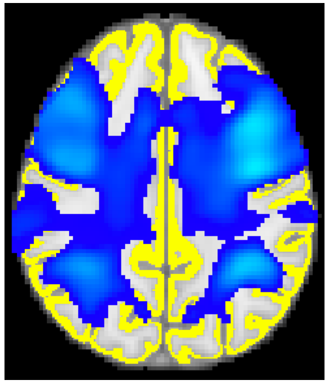 |
| | Hard | 52 | Same as 80% | Scan 79<br>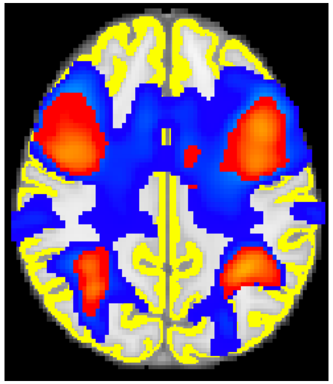 | Scan 155<br>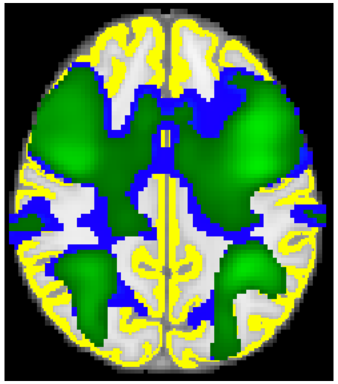 | Scan 200<br>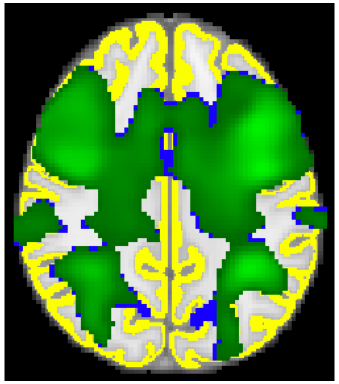 | 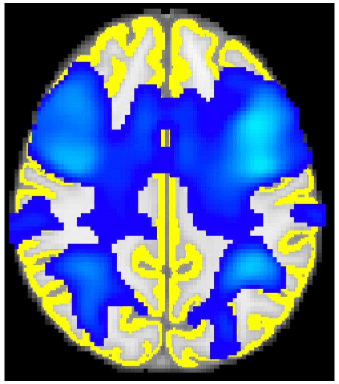 |
| **4** | Hard | 51 | Scan 79 | Scan 89 | Scan 155 | No early stoppping | |

| | | | | | | | |
|---|---|---|---|---|---|---|---|
| | | | 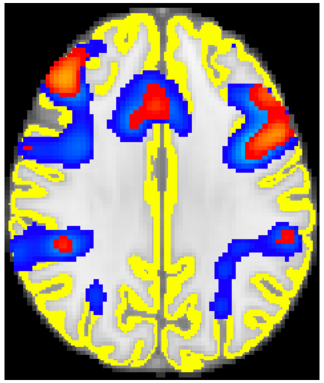 | 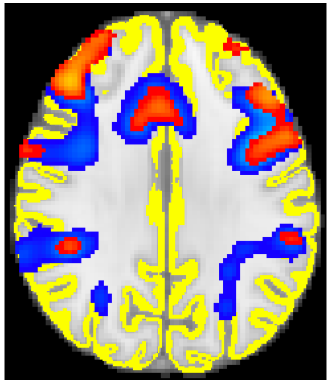 | 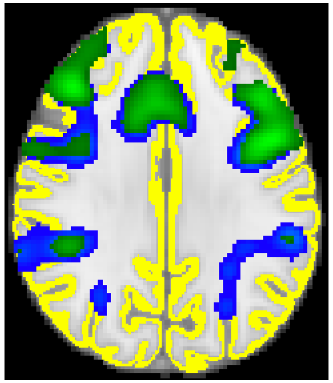 | | 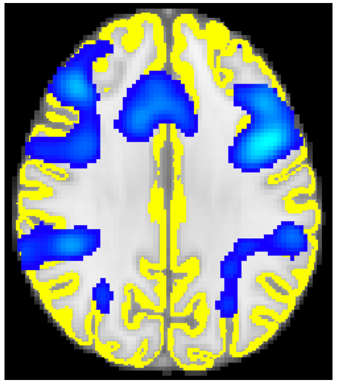 |
| **5** | Hard | 56 | Same as 80% | Scan 79 | Scan 155 | No early stopping | |
| | | | | 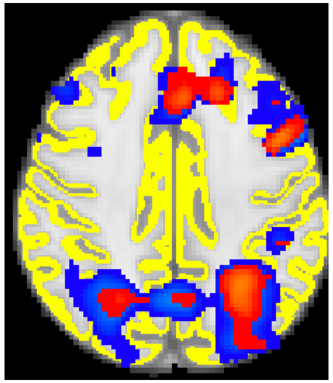 | 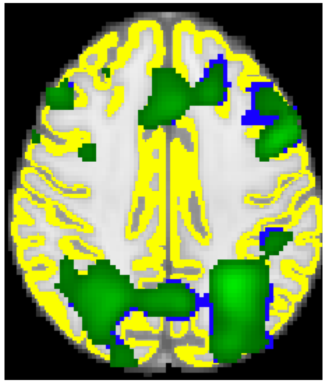 | | 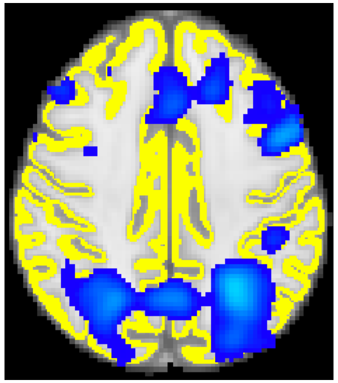 |
| **6** | Easy | 53 | Same as 80% | Scan 79 | Scan 155 | Scan 180 | |
| | | | | 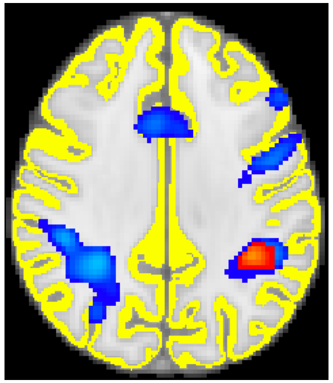 | 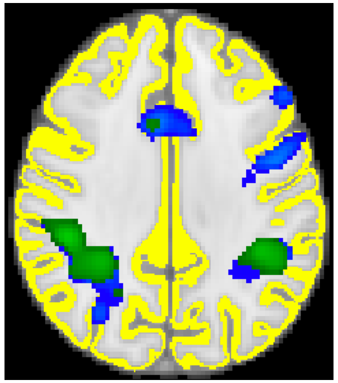 | 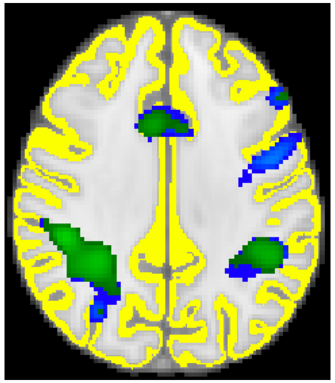 | 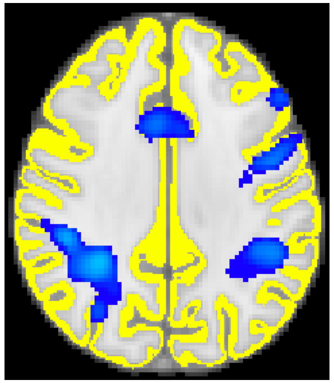 |
| **7** | Easy | 54 | Scan 79 | Scan 112 | Scan 155 | Scan 164 | |

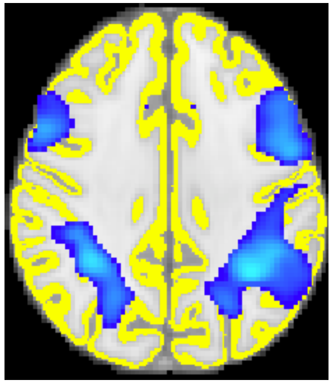
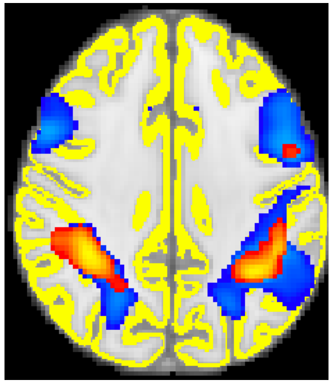
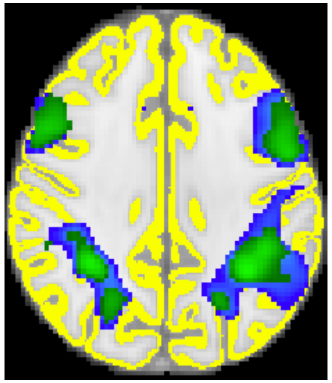
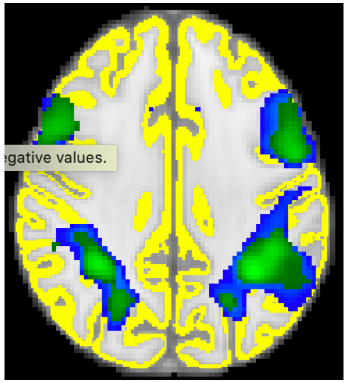

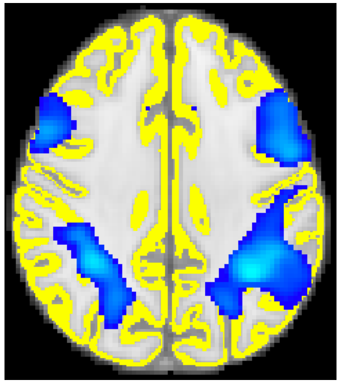

| | | | | | | |
|---|---|---|---|---|---|---|
| | Hard | 54 | Scan 80 | Scan 95 | Same as 80% | Scan 155 |

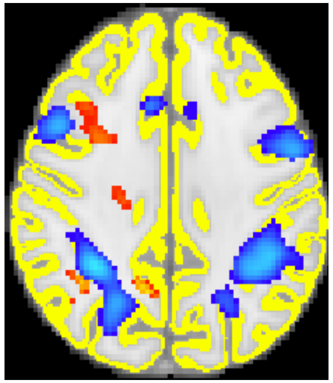
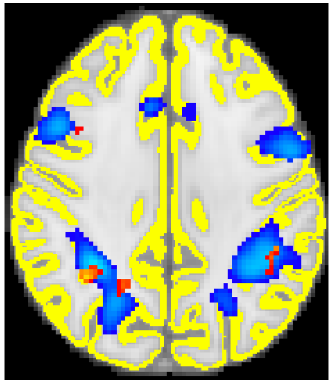
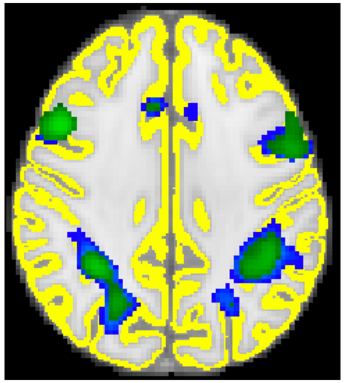
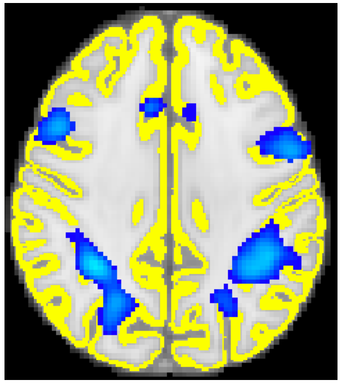

| | | | | | | |
|---|---|---|---|---|---|---|
| **9** | Easy | 55 | Same as 80% | Scan 79 | Same as 80% | Scan 155 |

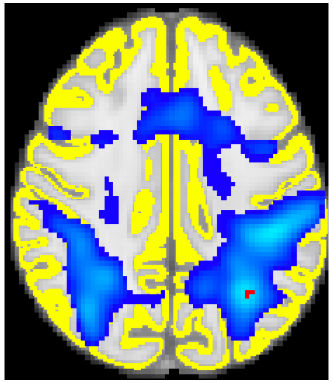
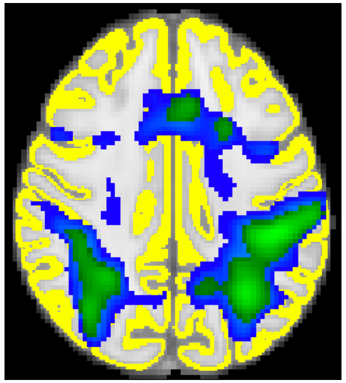
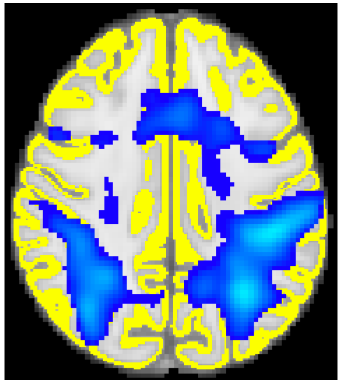

| | | | | | | |
|---|---|---|---|---|---|---|
| | Hard | 58 | Scan 79 | Scan 86 | Same as 80% | Scan 155 |

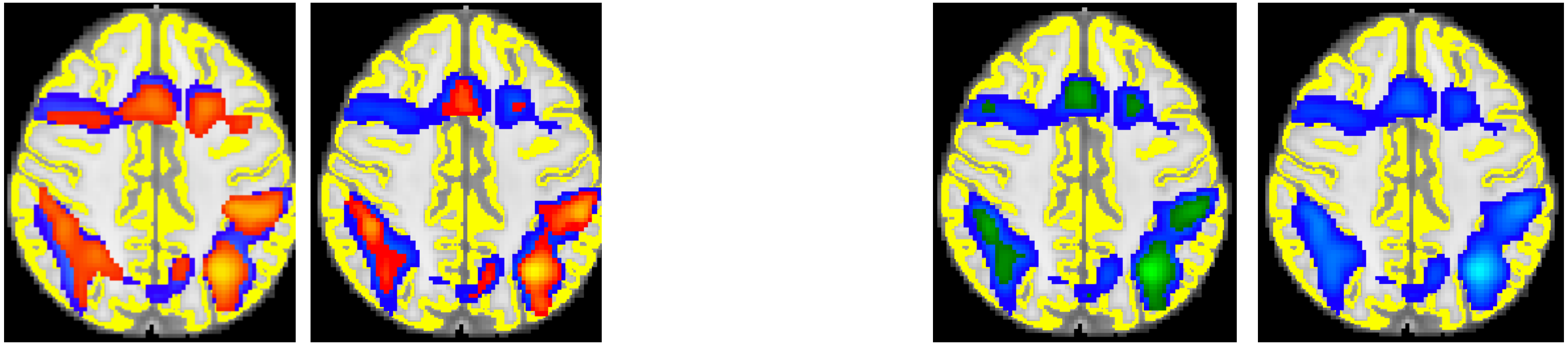

**10** Hard 52 Scan 79 Scan 98 Same as 80% Scan 155

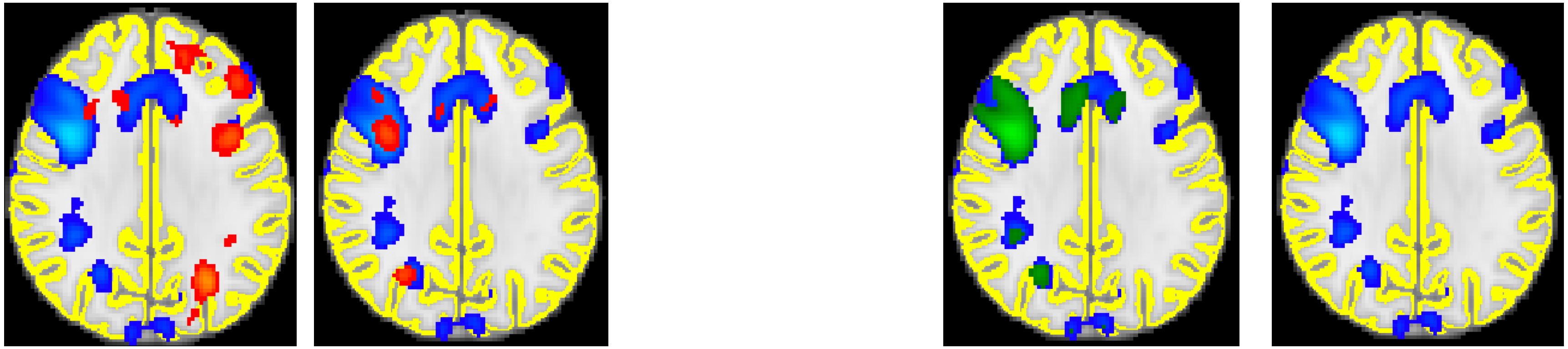

**11** Easy 58 Scan 79 Scan 89 Scan 155 Scan 186

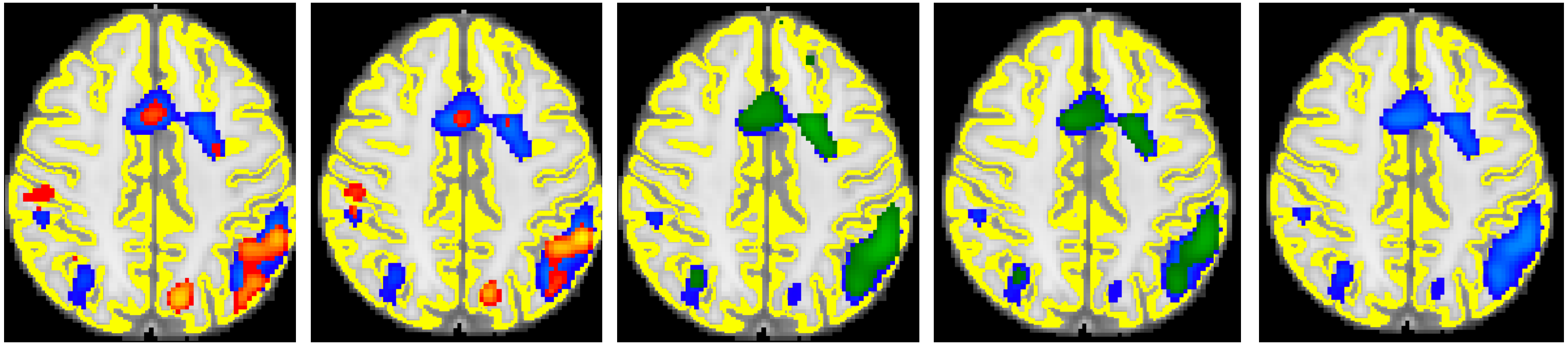

**EPT**

| | | | | | | | |
|---|---|---|---|---|---|---|---|
| **13** | Easy | | | Virtually no overlap to show visually | | | |
| | Hard | 57 | Same as 80% | Scan 79 | Scan 155 | Scan 158 | |
| | | | | 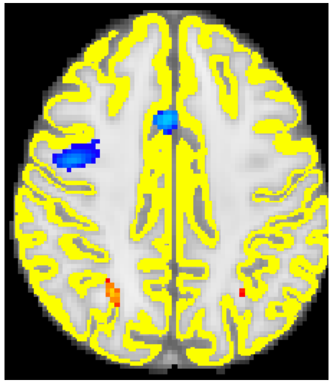 | 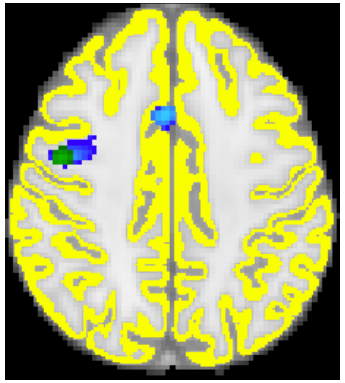 | 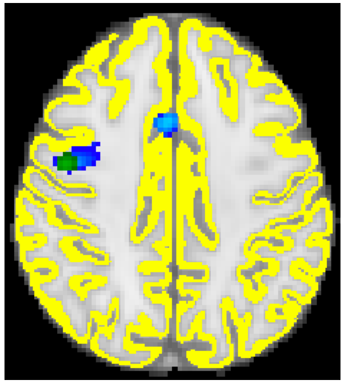 | 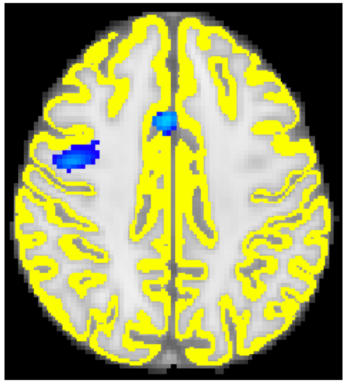 |
| **14** | Hard | 56 | Same as 80% | Scan 79 | Scan 155 | No early stopping | |
| | | | | 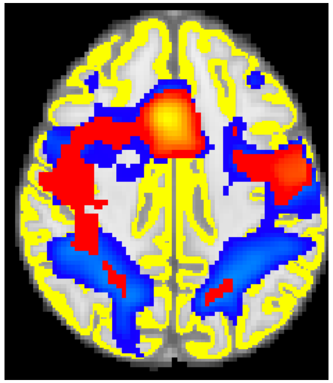 | 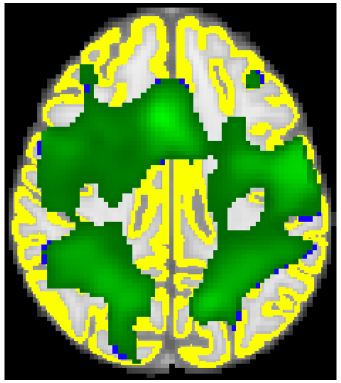 | | 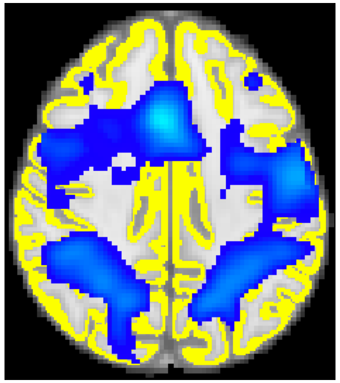 |
| **15** | Easy | 54 | Scan 107 | Scan 154 | Same as 80% | Scan 155 | |

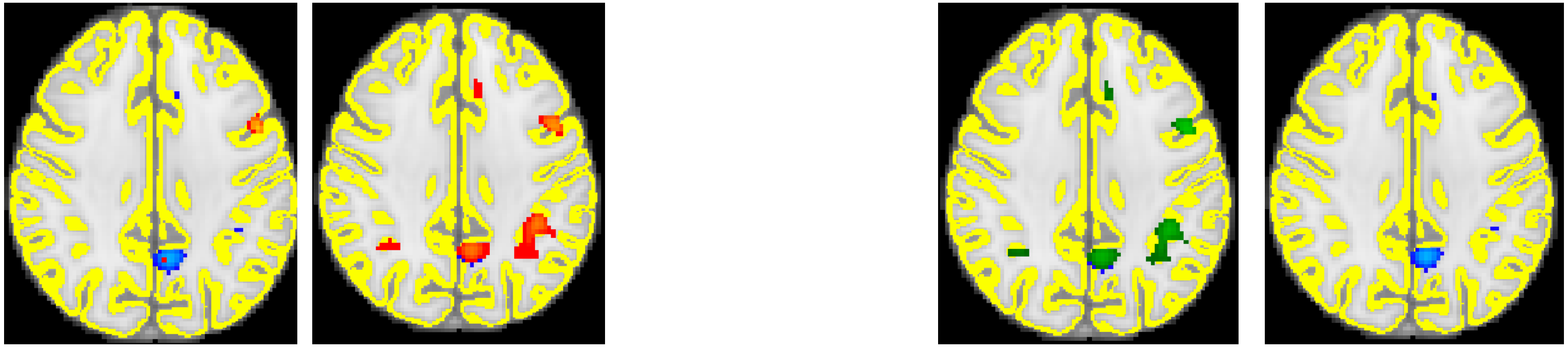

Hard 51 Scan 79 Scan 87 Scan 155 No early stopping

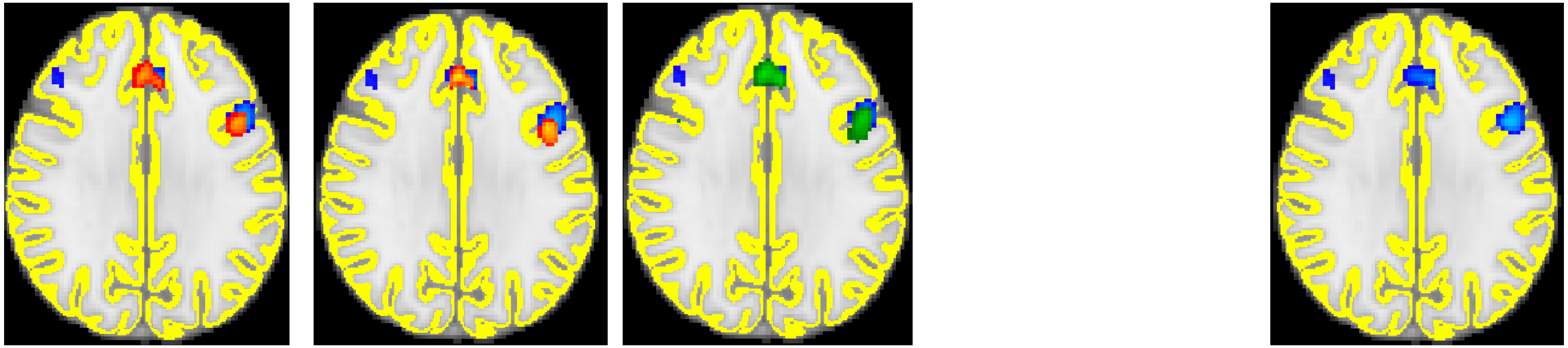

**17** Easy 57 Same as 80% Scan 79 Scan 155 Scan 216

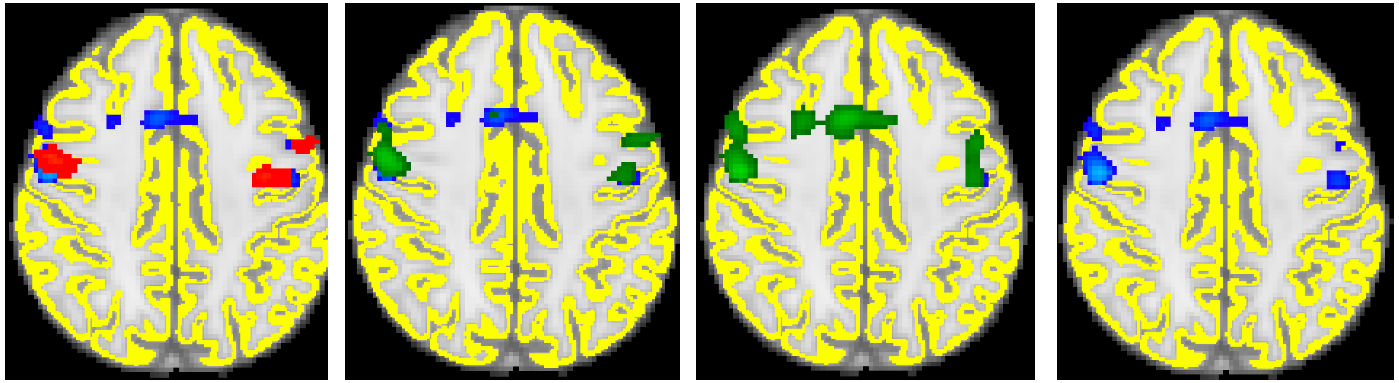

Hard 58 Same as 80% Scan 79 Scan 155 Scan 218

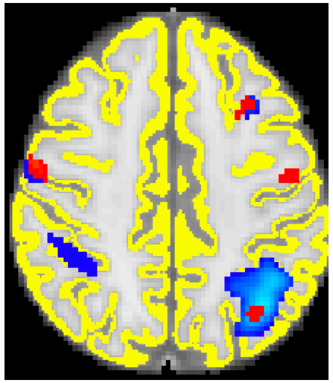 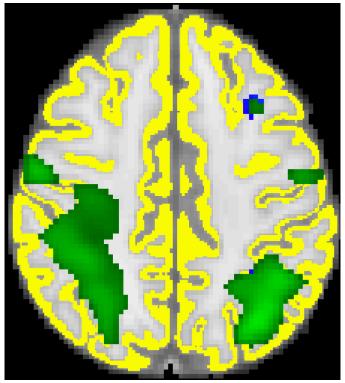 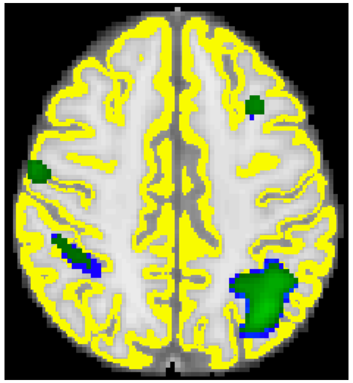 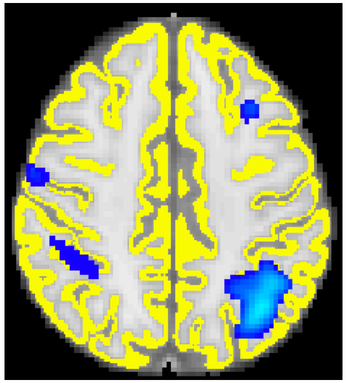

| | | | | | | |
|---|---|---|---|---|---|---|
| **18** | Easy | 49 | Same as 80% | Scan 79 | Same as 80% | Scan 155 |

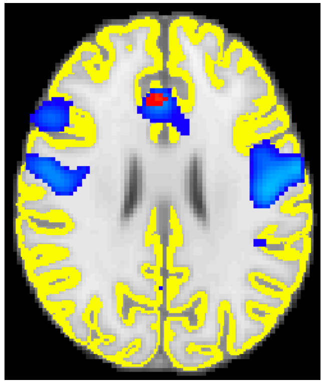 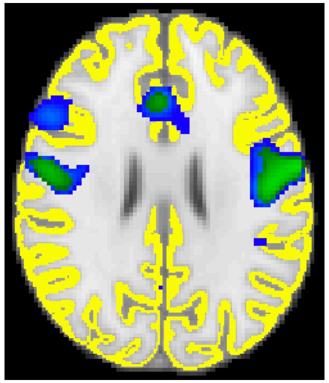 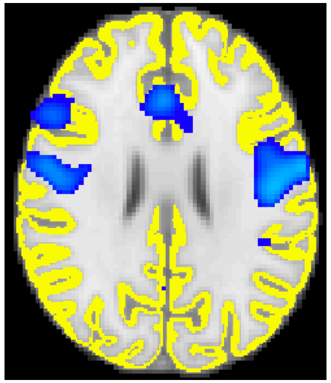

| | | | | | | |
|---|---|---|---|---|---|---|
| **19** | Hard | 61 | Same as 80% | Scan 79 | Same as 80% | Scan 155 |

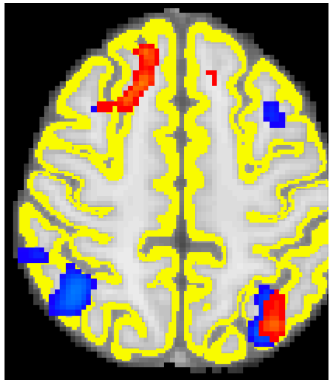 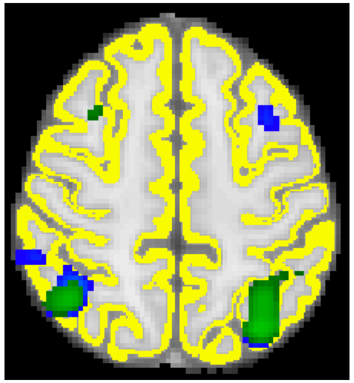 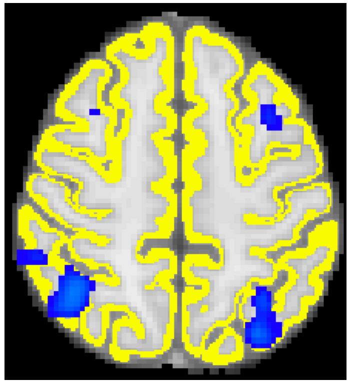

| | | | | | | |
|---|---|---|---|---|---|---|
| **20** | Easy | 54 | Same as 80% | Scan 79 | Same as 80% | Scan 155 |

| | | | | | | | |
|---|---|---|---|---|---|---|---|
| | | | | 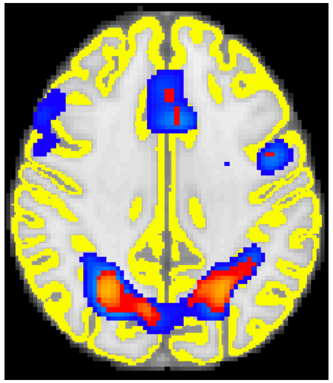 | | 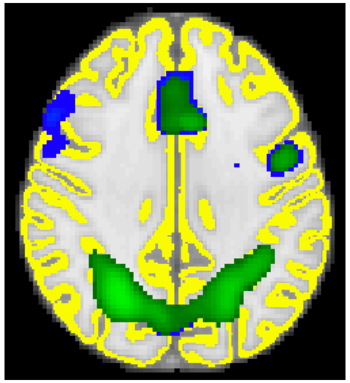 | 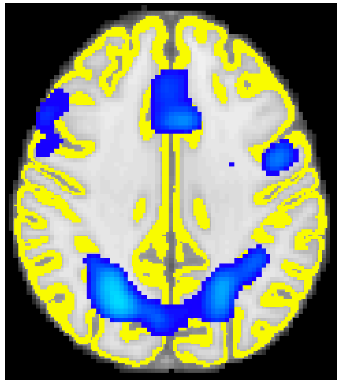 |
| | Hard | 54 | Same as 80% | Scan 79 | Scan 155 | Scan 159 | |
| | | | | 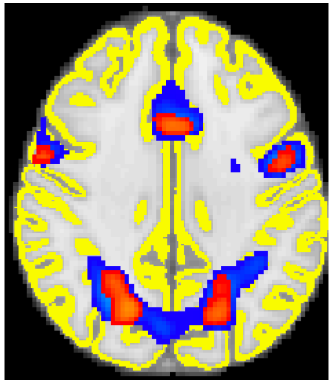 | 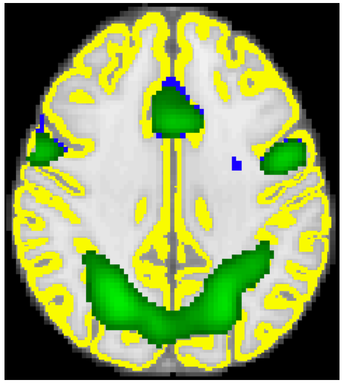 | 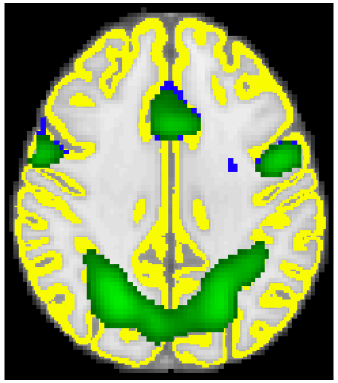 | 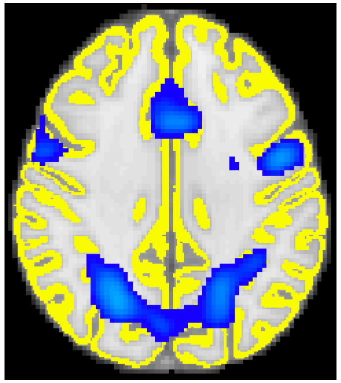 |
| **21** | Hard | 28 | Same as 80% | Scan 79 | No early stopping | No early stopping | |
| | | | | 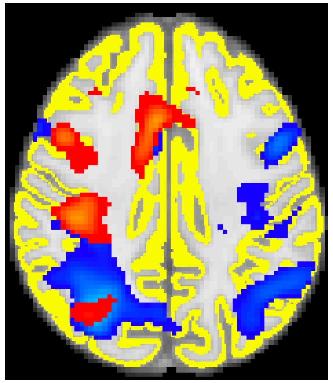 | | | 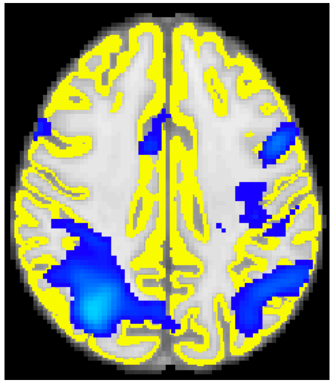 |

**Table S2:** Plots of the percentage of voxels classified as active and non-active at each scan number for each difficulty level for each subject. Information is given from scan 79 to 238.

| Subject | Total number of voxels | Classification of voxels by scan number |
|---|---|---|
| **Control 1** | 115,062 | 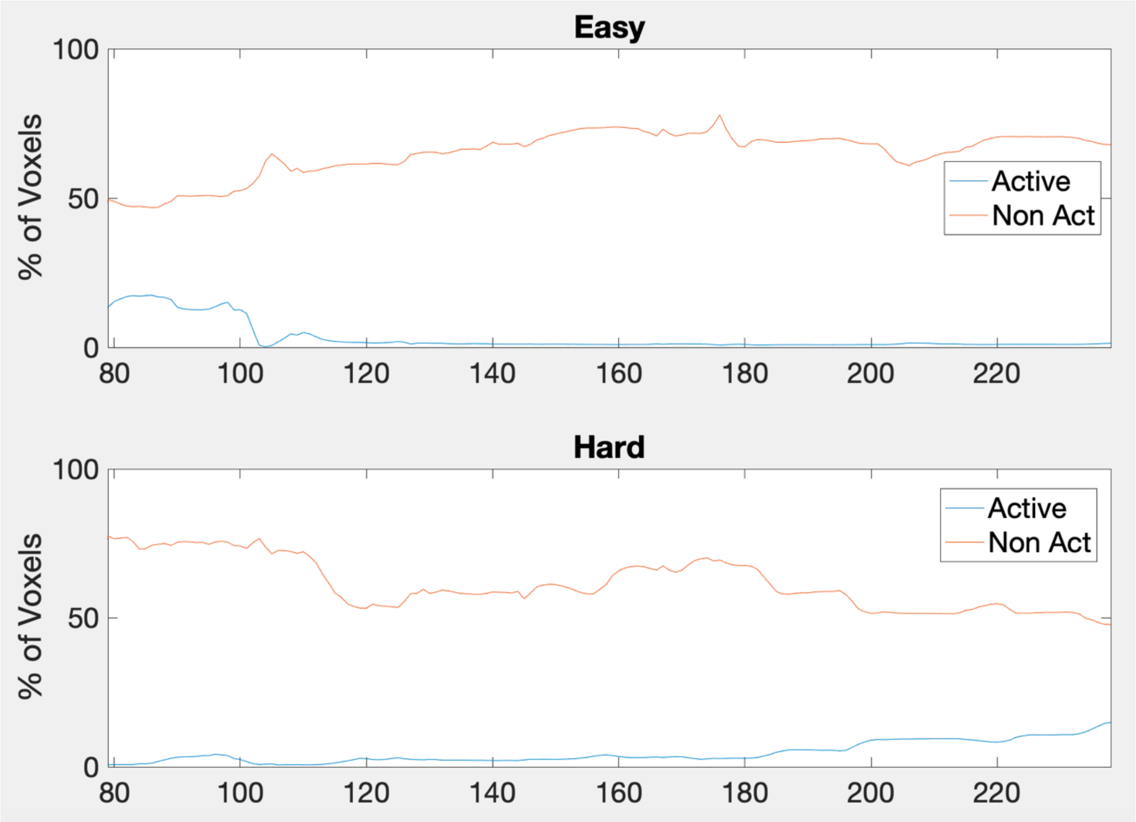 |

| | | |
|---|---|---|
| **2** | 113,564 | 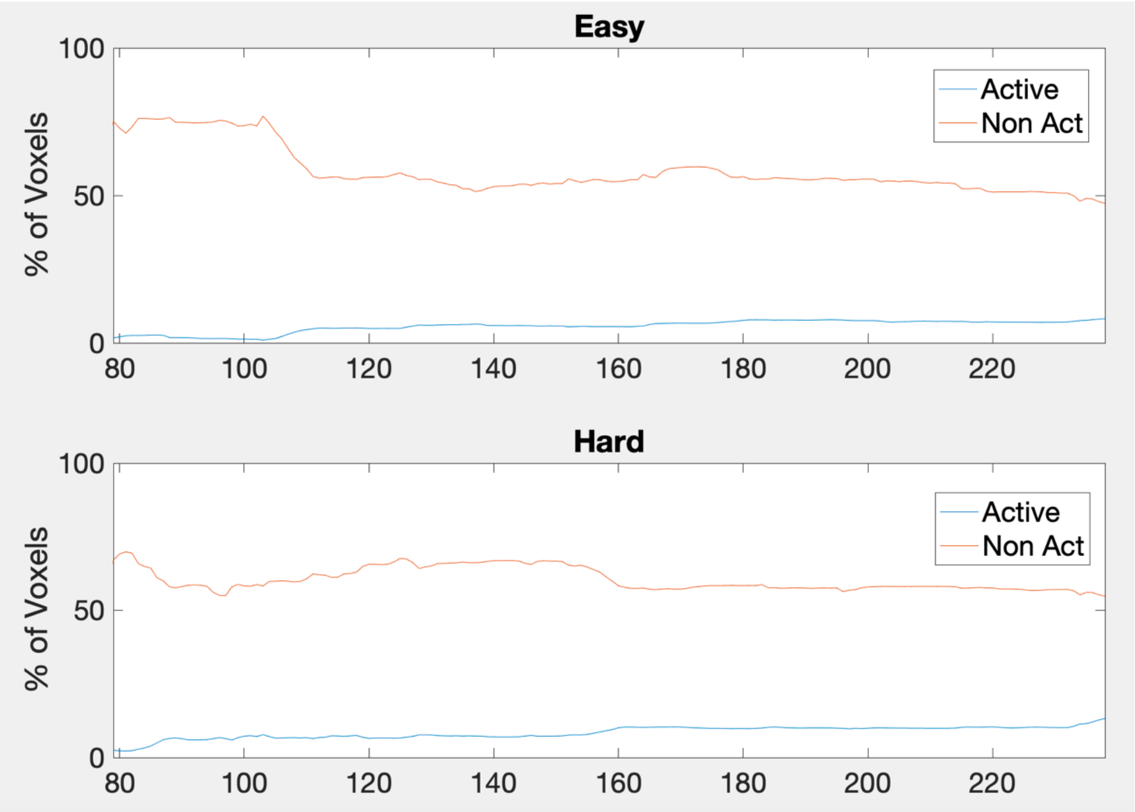 Easy<br>100<br>50<br>0<br>% of Voxels<br>80<br>100<br>120<br>140<br>160<br>180<br>200<br>220<br>Active<br>Non Act<br>Hard<br>100<br>50<br>0<br>% of Voxels<br>80<br>100<br>120<br>140<br>160<br>180<br>200<br>220<br>Active<br>Non Act |

| | | |
|---|---|---|
| **3** | 77,359 | 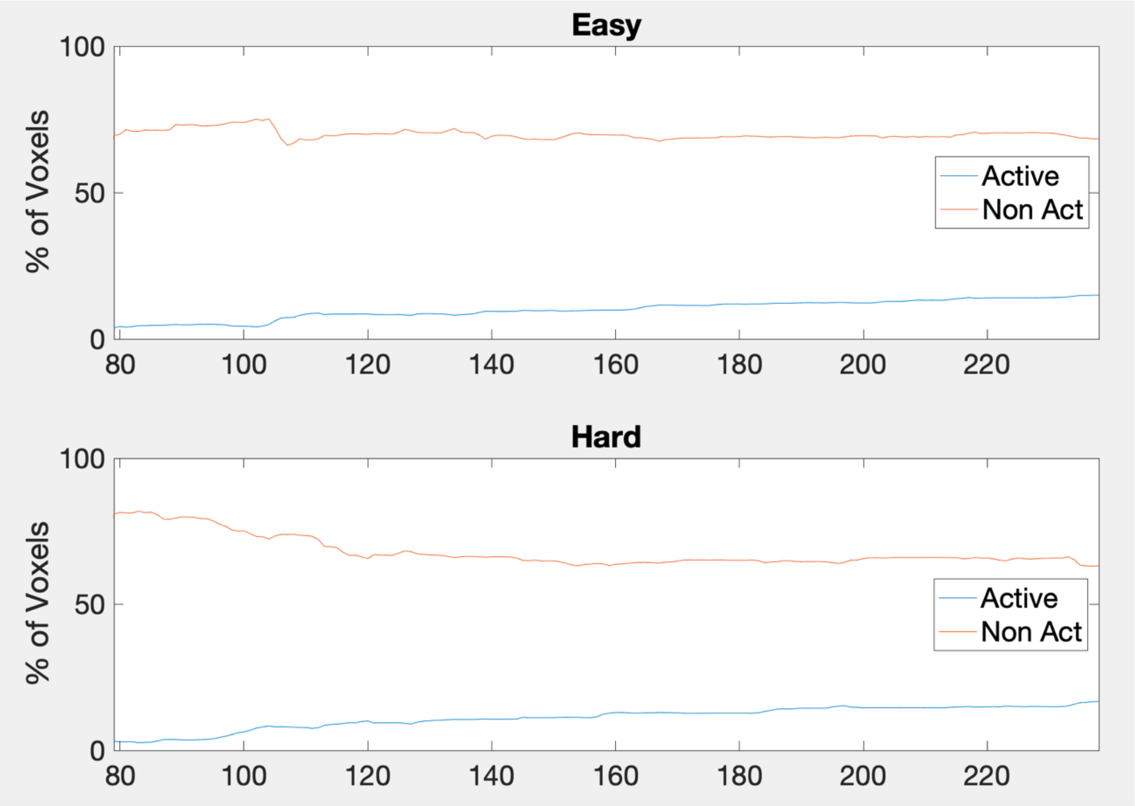 Easy<br>100<br>50<br>0<br>% of Voxels<br>80<br>100<br>120<br>140<br>160<br>180<br>200<br>220<br>Active<br>Non Act<br>Hard<br>100<br>50<br>0<br>% of Voxels<br>80<br>100<br>120<br>140<br>160<br>180<br>200<br>220<br>Active<br>Non Act |

| | | |
|---|---|---|
| **4** | 103,591 | 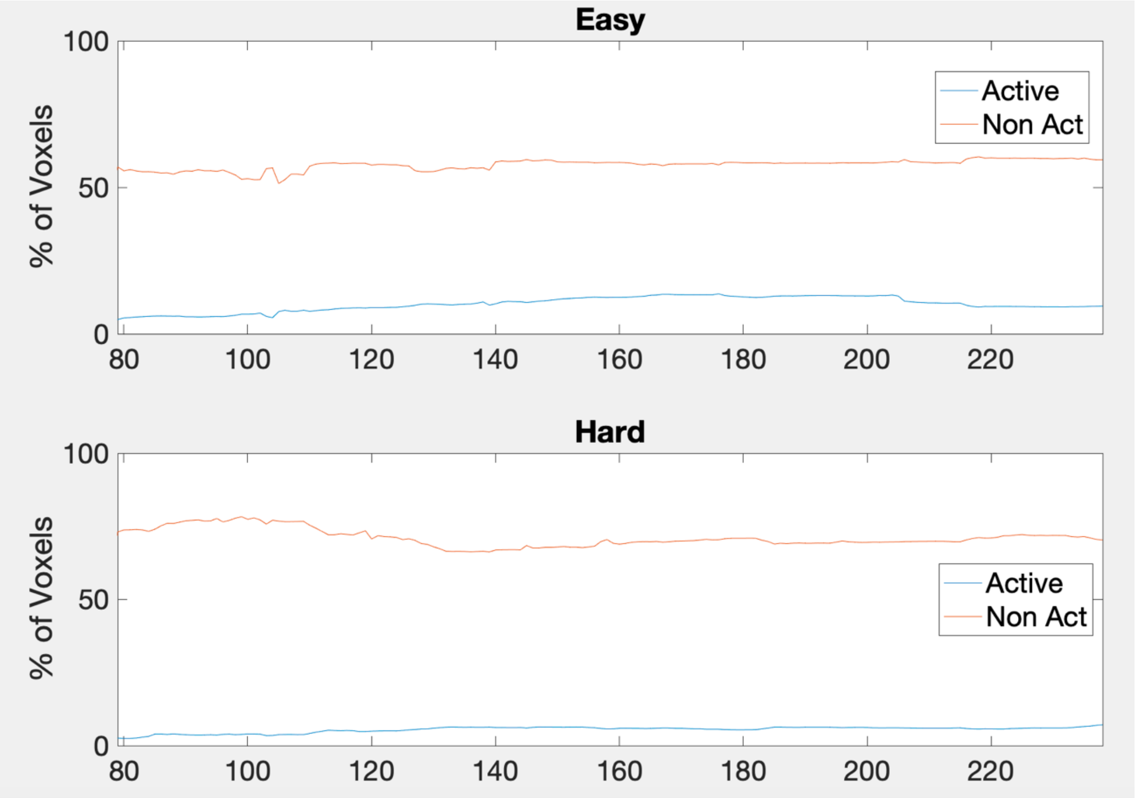 |

Easy
100
50
0
% of Voxels
80
100
120
140
160
180
200
220
Active
Non Act
Hard
100
50
0
% of Voxels
80
100
120
140
160
180
200
220
Active
Non Act

| | | |
|---|---|---|
| **5** | 114,260 | 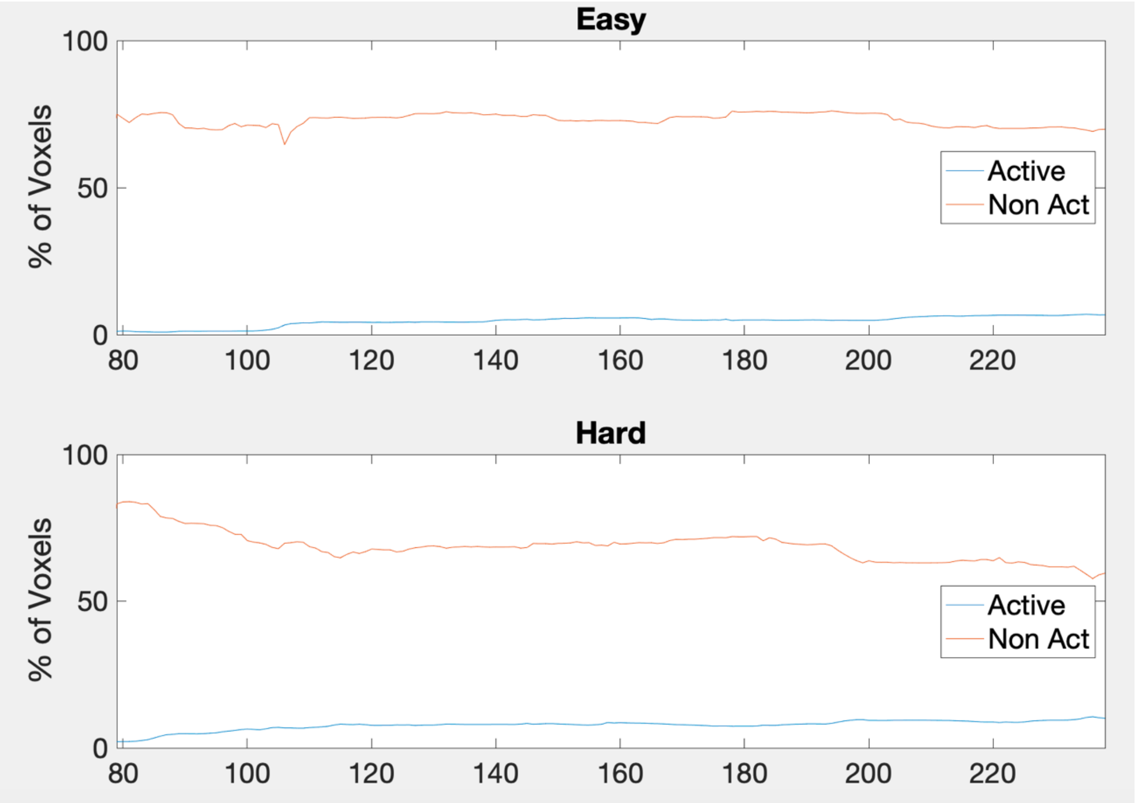 |

Easy
% of Voxels
100
50
0
80
100
120
140
160
180
200
220
Active
Non Act
Hard
% of Voxels
100
50
0
80
100
120
140
160
180
200
220
Active
Non Act

| | | |
|---|---|---|
| **6** | 121,353 | 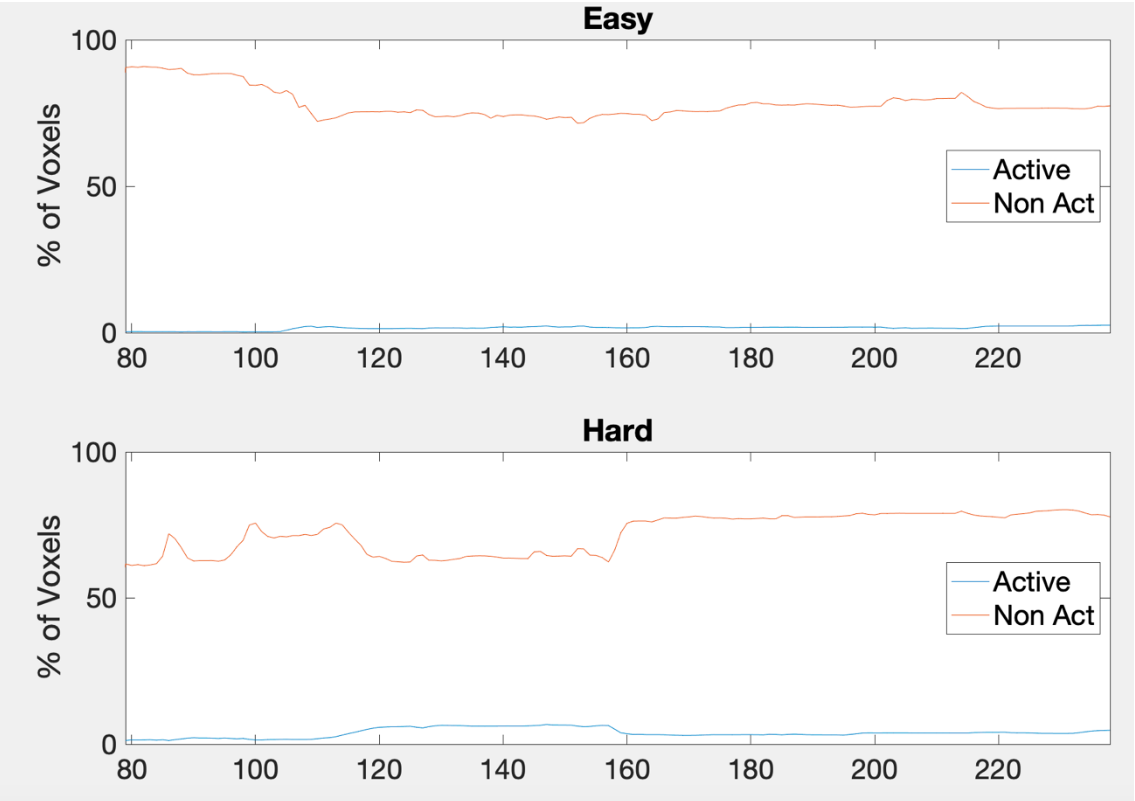Easy<br>% of Voxels<br>Active<br>Non Act<br>Hard<br>% of Voxels<br>Active<br>Non Act |

| | | |
|---|---|---|
| **7** | 107,406 | 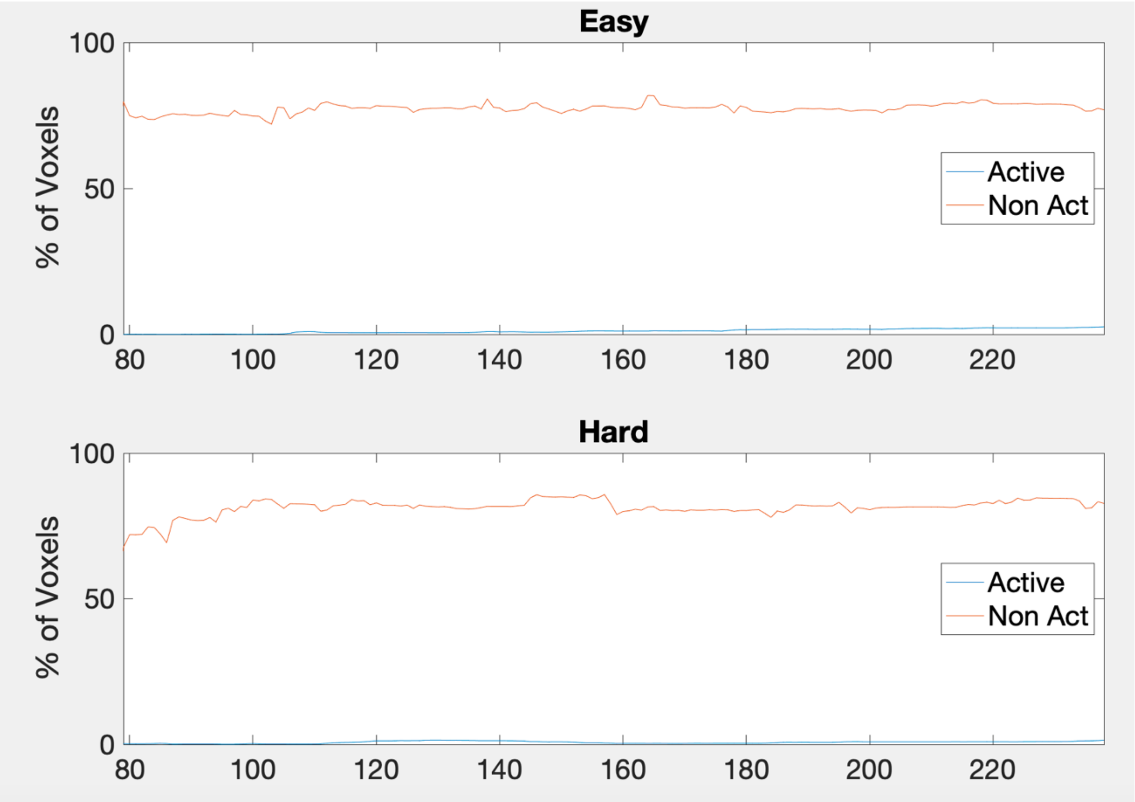 Easy<br>100<br>50<br>0<br>% of Voxels<br>80 100 120 140 160 180 200 220<br>Active<br>Non Act<br>Hard<br>100<br>50<br>0<br>% of Voxels<br>80 100 120 140 160 180 200 220<br>Active<br>Non Act |

| | | |
|---|---|---|
| **8** | 106,267 | 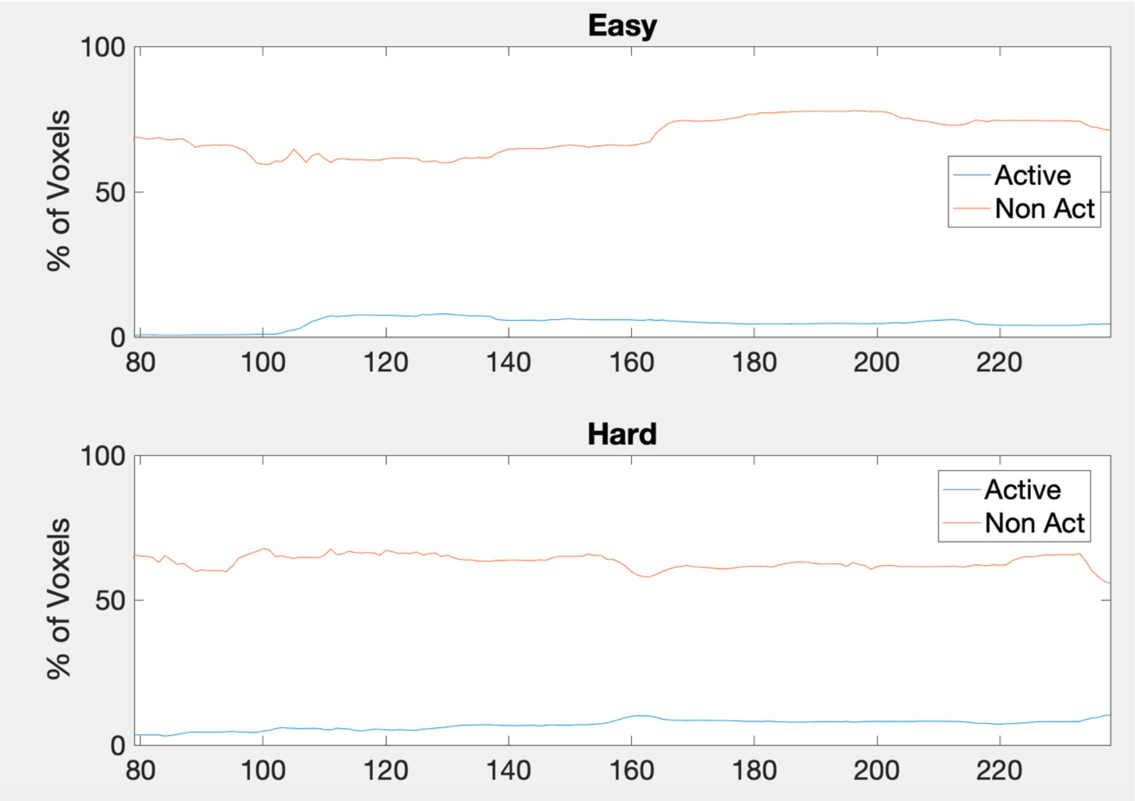Easy<br>% of Voxels<br>100<br>50<br>0<br>80<br>100<br>120<br>140<br>160<br>180<br>200<br>220<br>Active<br>Non Act<br>Hard<br>% of Voxels<br>100<br>50<br>0<br>80<br>100<br>120<br>140<br>160<br>180<br>200<br>220<br>Active<br>Non Act |

| | | |
|---|---|---|
| **9** | 121,195 | 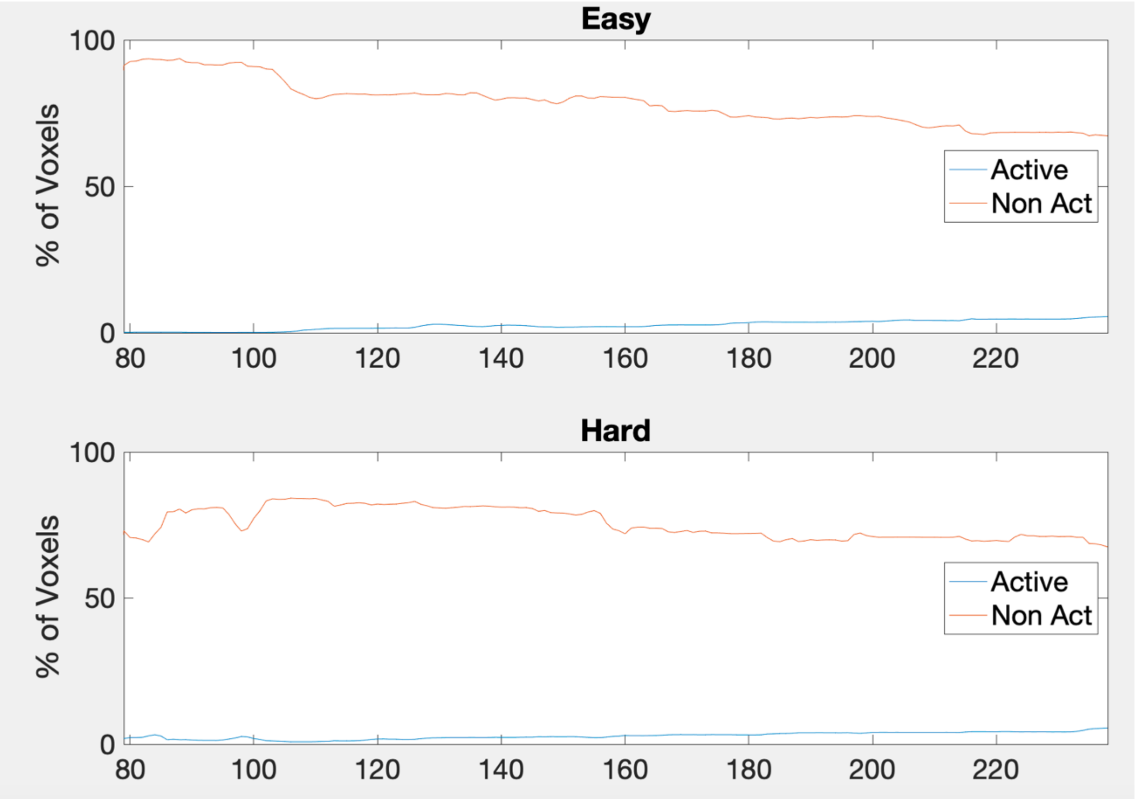 |

Easy
100
50
0
% of Voxels
80
100
120
140
160
180
200
220
Active
Non Act
Hard
100
50
0
% of Voxels
80
100
120
140
160
180
200
220
Active
Non Act

| | | |
|---|---|---|
| **10** | 96,565 | 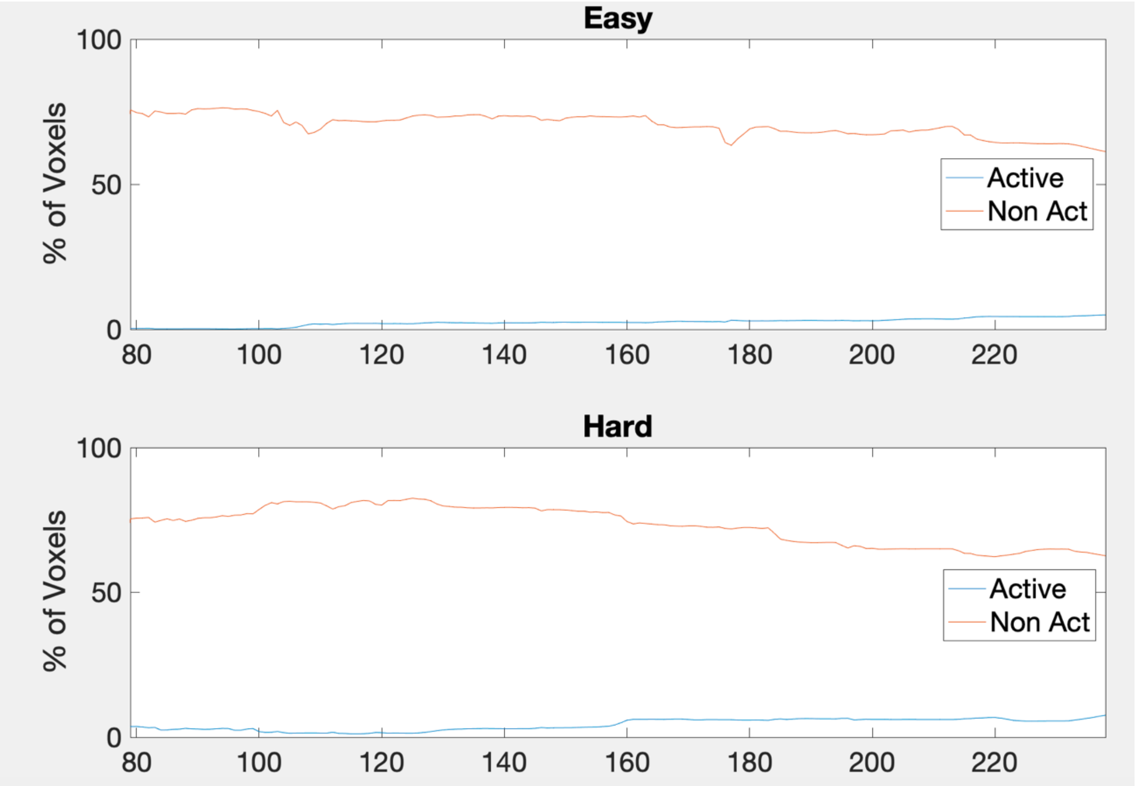 |

Easy
100
50
0
% of Voxels
80
100
120
140
160
180
200
220
Active
Non Act
Hard
100
50
0
% of Voxels
80
100
120
140
160
180
200
220
Active
Non Act

| | | |
|---|---|---|
| **11** | 107,016 | 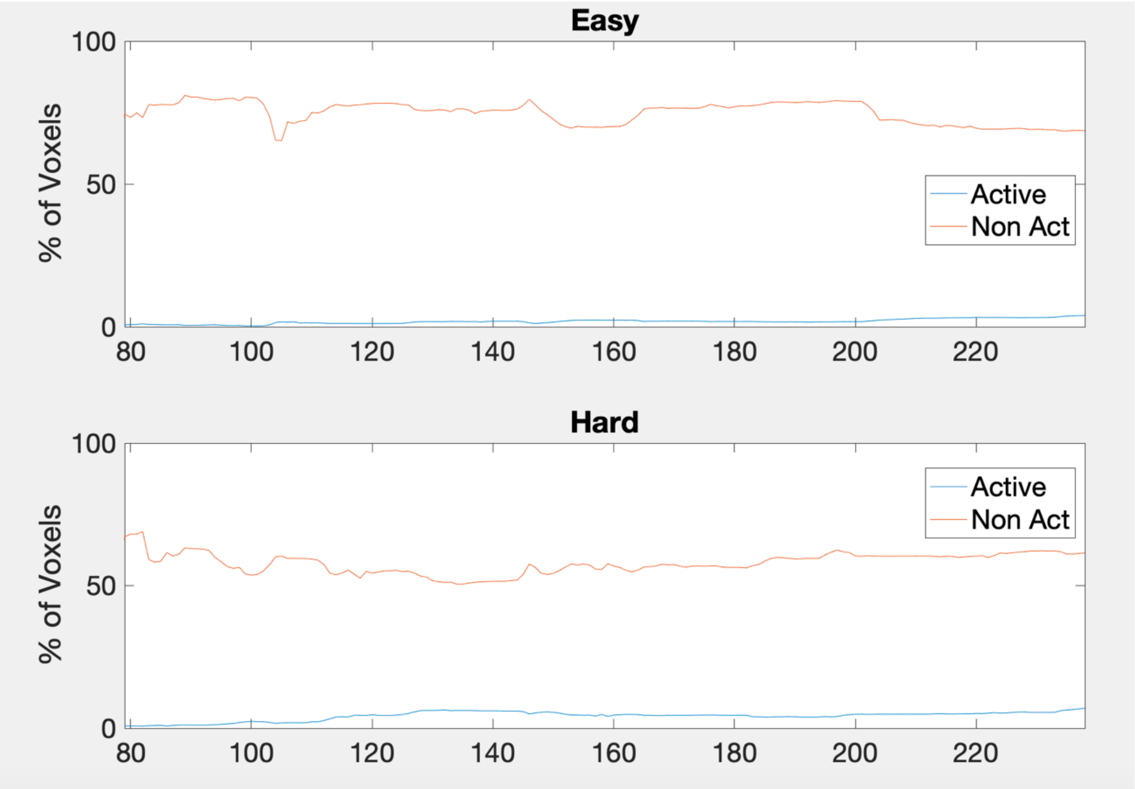Easy<br>% of Voxels<br>100<br>50<br>0<br>80<br>100<br>120<br>140<br>160<br>180<br>200<br>220<br>Active<br>Non Act<br>Hard<br>% of Voxels<br>100<br>50<br>0<br>80<br>100<br>120<br>140<br>160<br>180<br>200<br>220<br>Active<br>Non Act |

| | | |
|---|---|---|
| **12** | 96,936 | 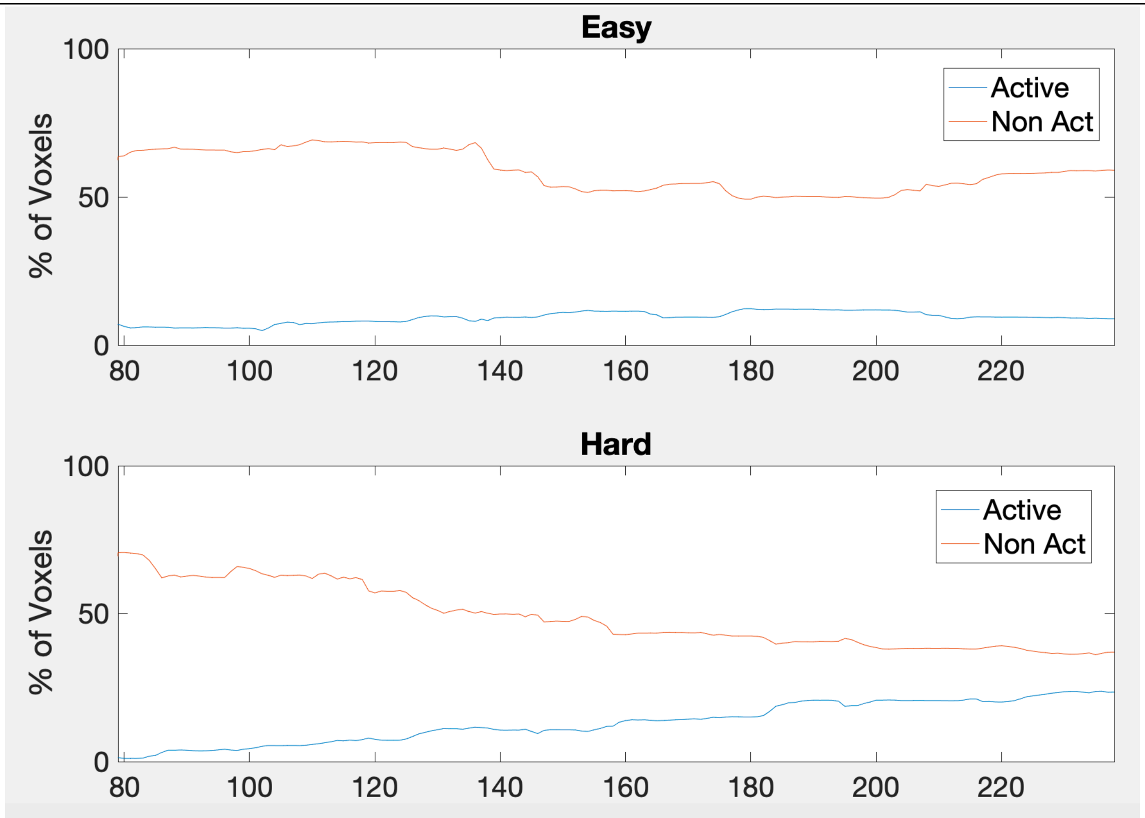 |
| **EPT** | | |

Easy
100
50
0
% of Voxels
80
100
120
140
160
180
200
220
Active
Non Act
Hard
100
50
0
% of Voxels
80
100
120
140
160
180
200
220
Active
Non Act

| | | |
|---|---|---|
| **13** | 94,623 | 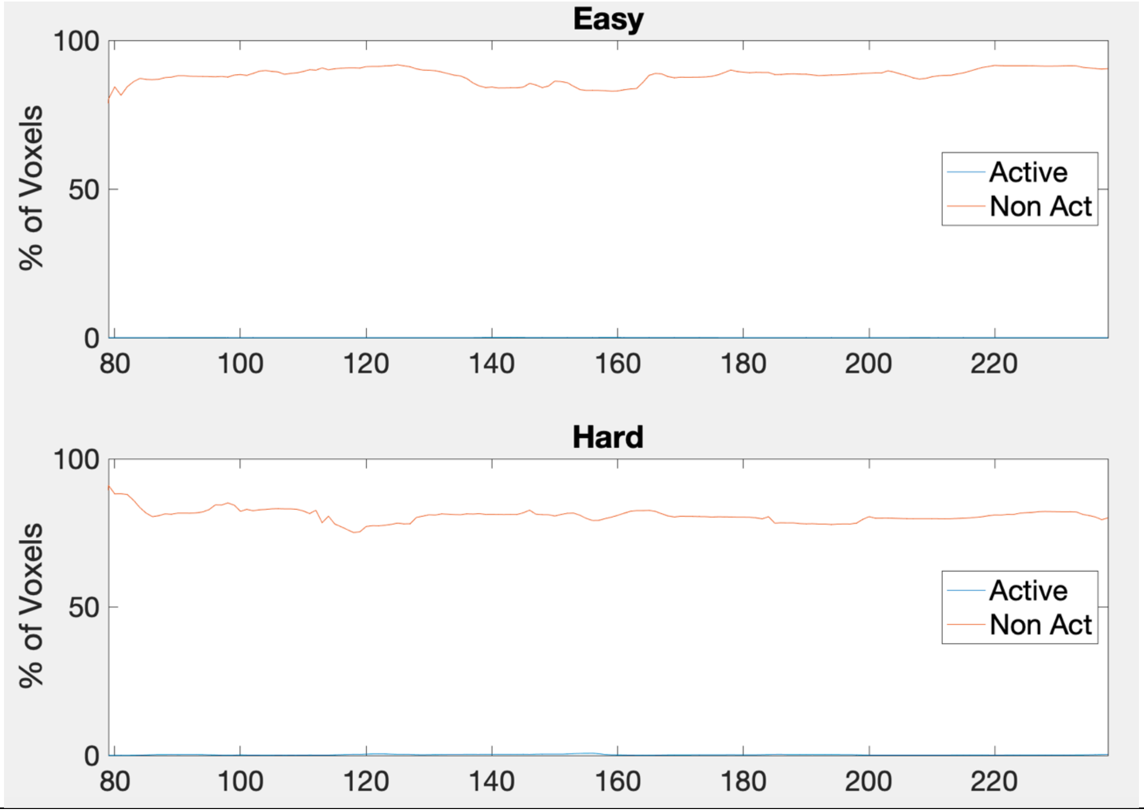Easy<br>% of Voxels<br>Active<br>Non Act<br>Hard<br>% of Voxels<br>Active<br>Non Act |

| | | |
|---|---|---|
| **14** | 94,905 | 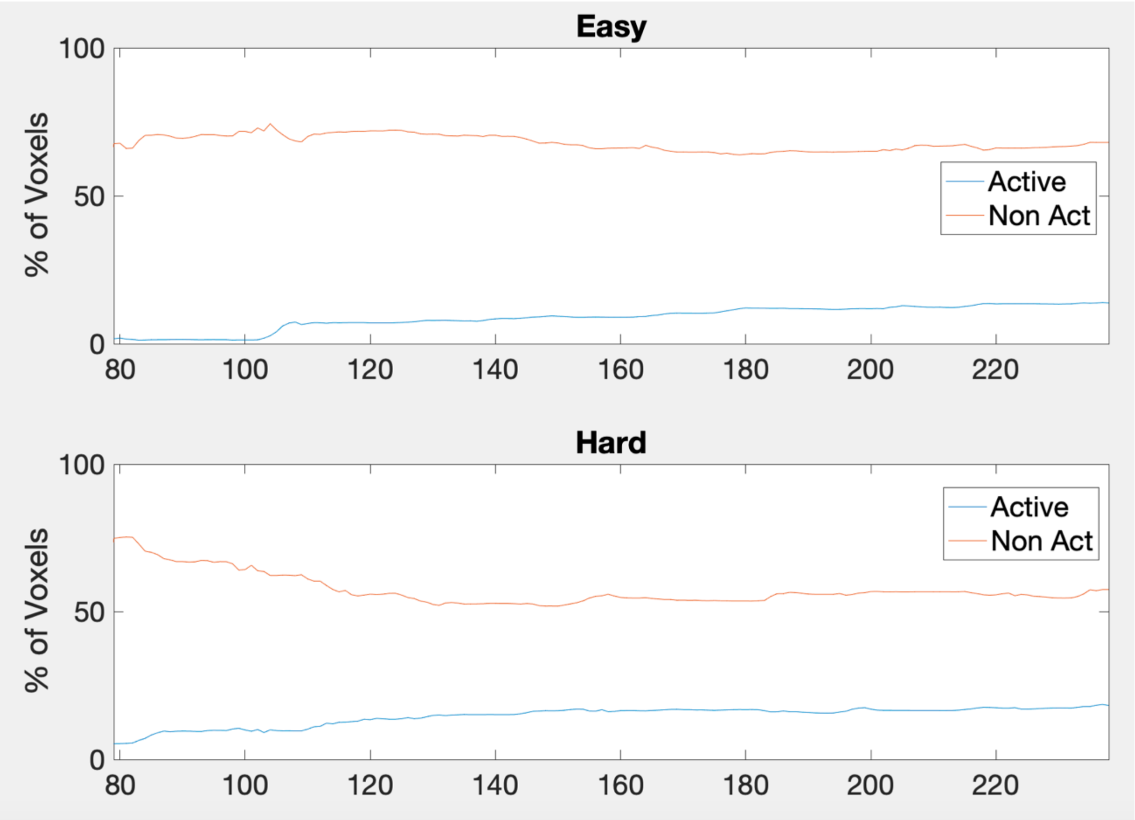 |

Easy
% of Voxels
100
50
0
80
100
120
140
160
180
200
220
Active
Non Act
Hard
% of Voxels
100
50
0
80
100
120
140
160
180
200
220
Active
Non Act

| | | |
|---|---|---|
| **15** | 98,799 | 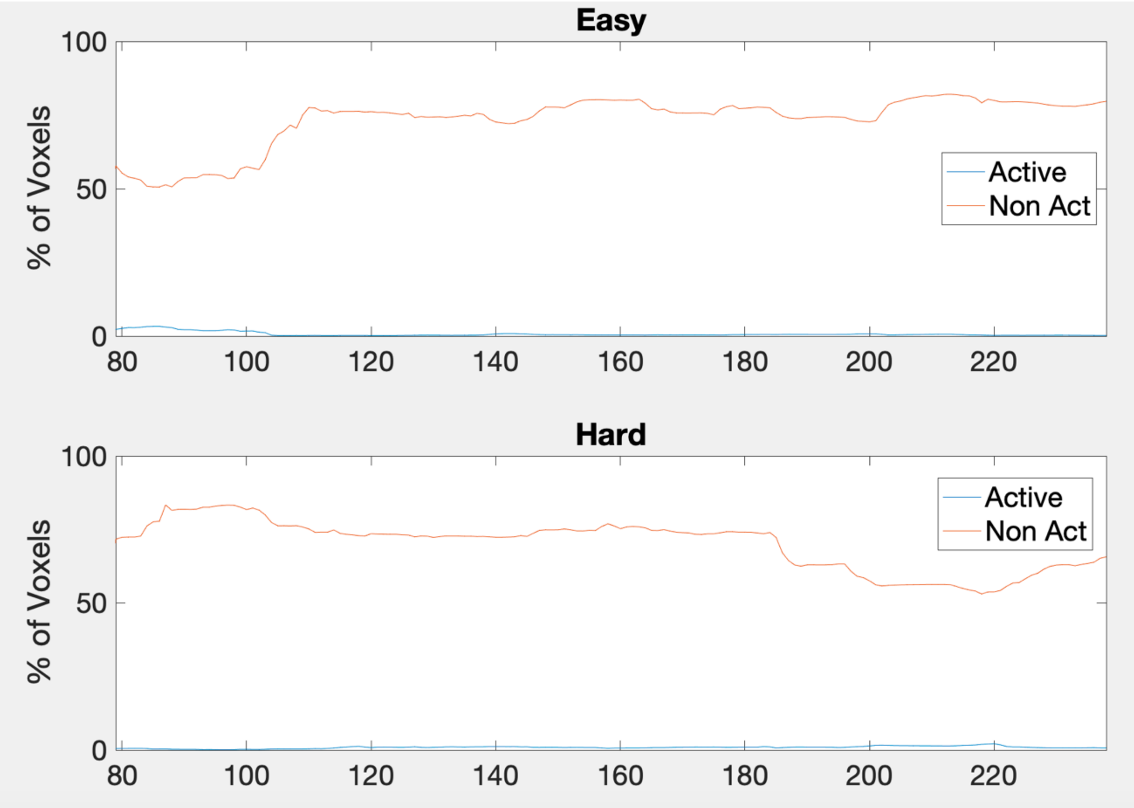 |

Easy
100
50
0
% of Voxels
80
100
120
140
160
180
200
220
Active
Non Act
Hard
100
50
0
% of Voxels
80
100
120
140
160
180
200
220
Active
Non Act

| | | |
|---|---|---|
| **16** | 118,098 | 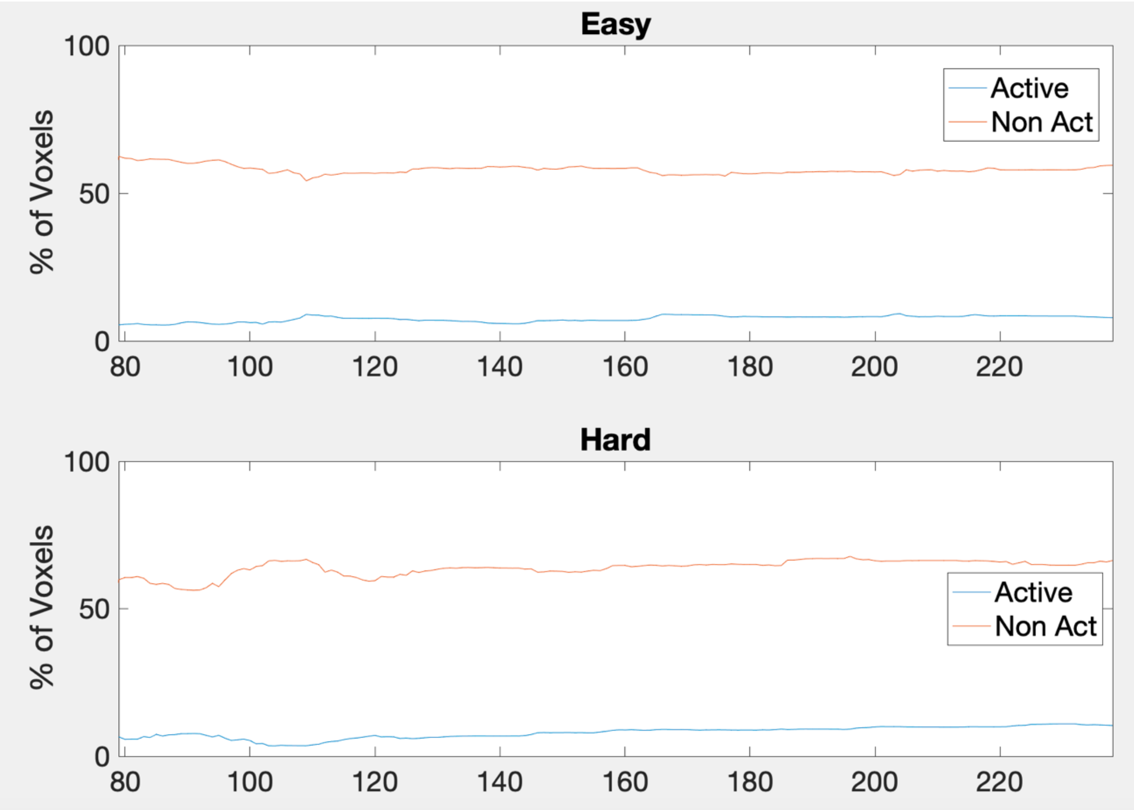 |

Easy
100
50
0
% of Voxels
80
100
120
140
160
180
200
220
Active
Non Act
Hard
100
50
0
% of Voxels
80
100
120
140
160
180
200
220
Active
Non Act

| | | |
|---|---|---|
| **17** | 124,749 | 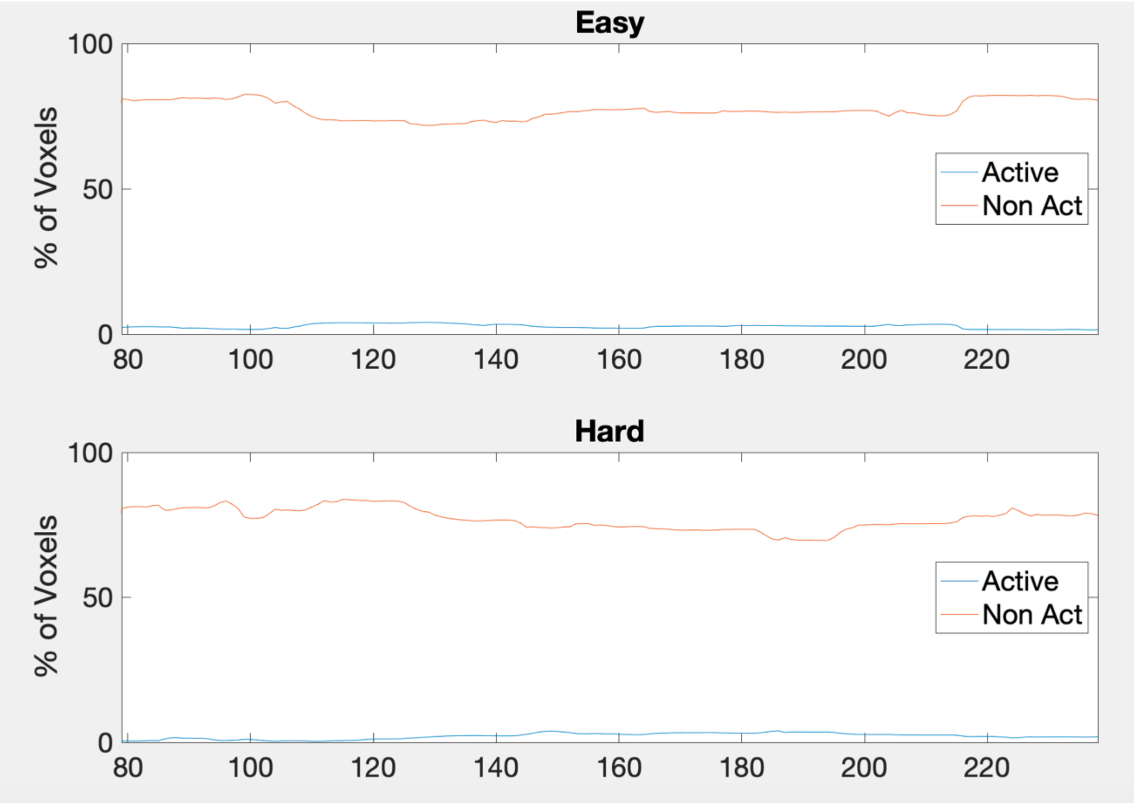Easy<br>% of Voxels<br>Active<br>Non Act<br>Hard<br>% of Voxels<br>Active<br>Non Act |

| | | |
|---|---|---|
| **18** | 97,437 | 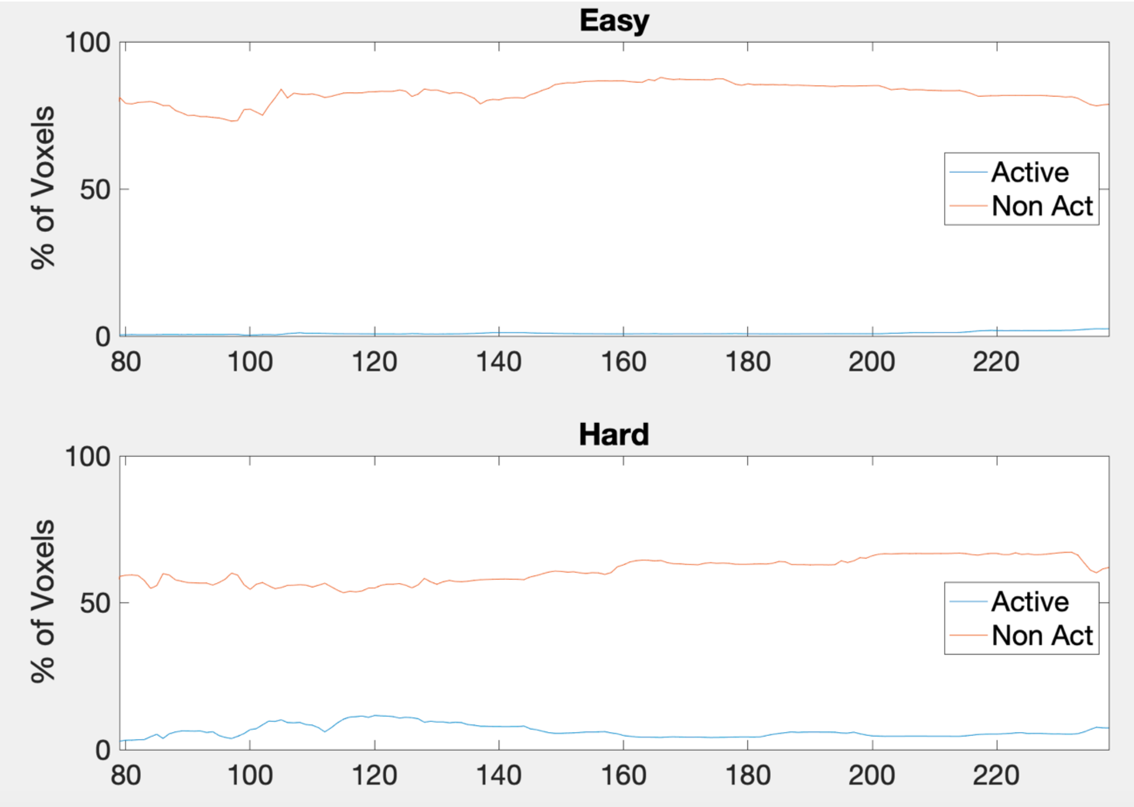Easy<br>% of Voxels<br>Active<br>Non Act<br>Hard<br>% of Voxels<br>Active<br>Non Act |

| | | |
|---|---|---|
| **19** | 135,379 | 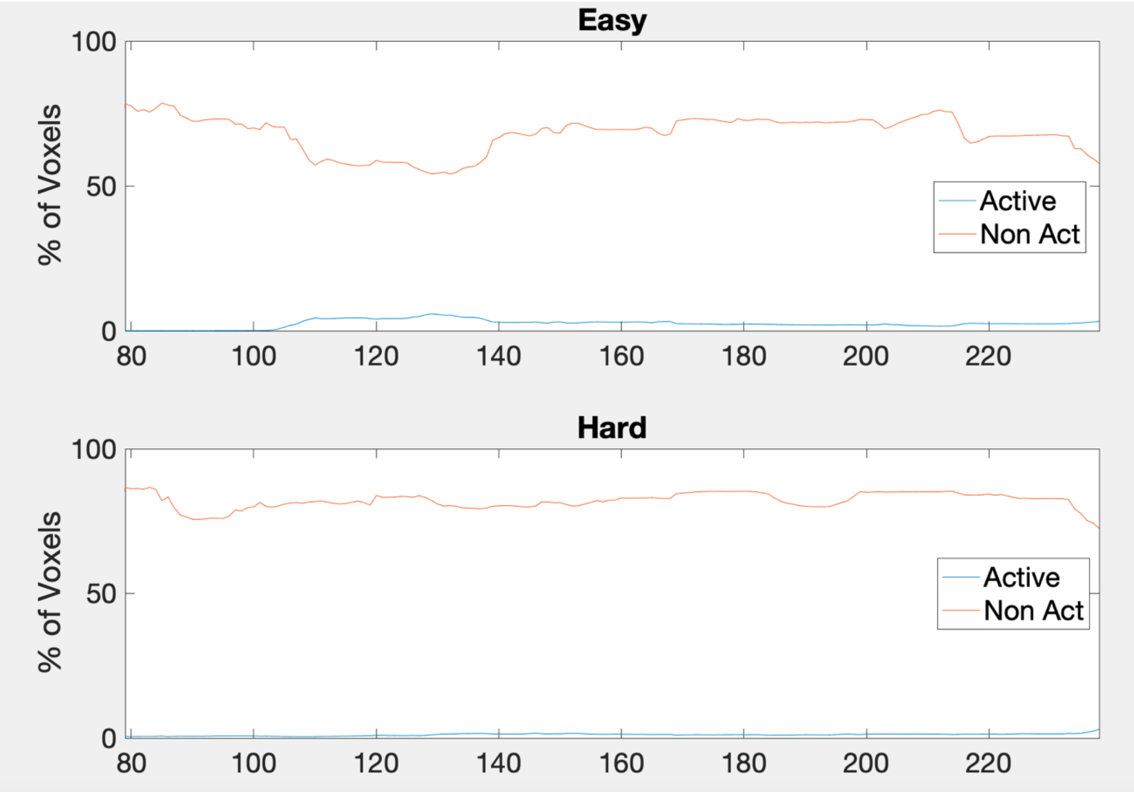 |

Easy
Hard
% of Voxels
Active
Non Act
100
50
0
80
100
120
140
160
180
200
220

| | | |
|---|---|---|
| **20** | 89,609 | 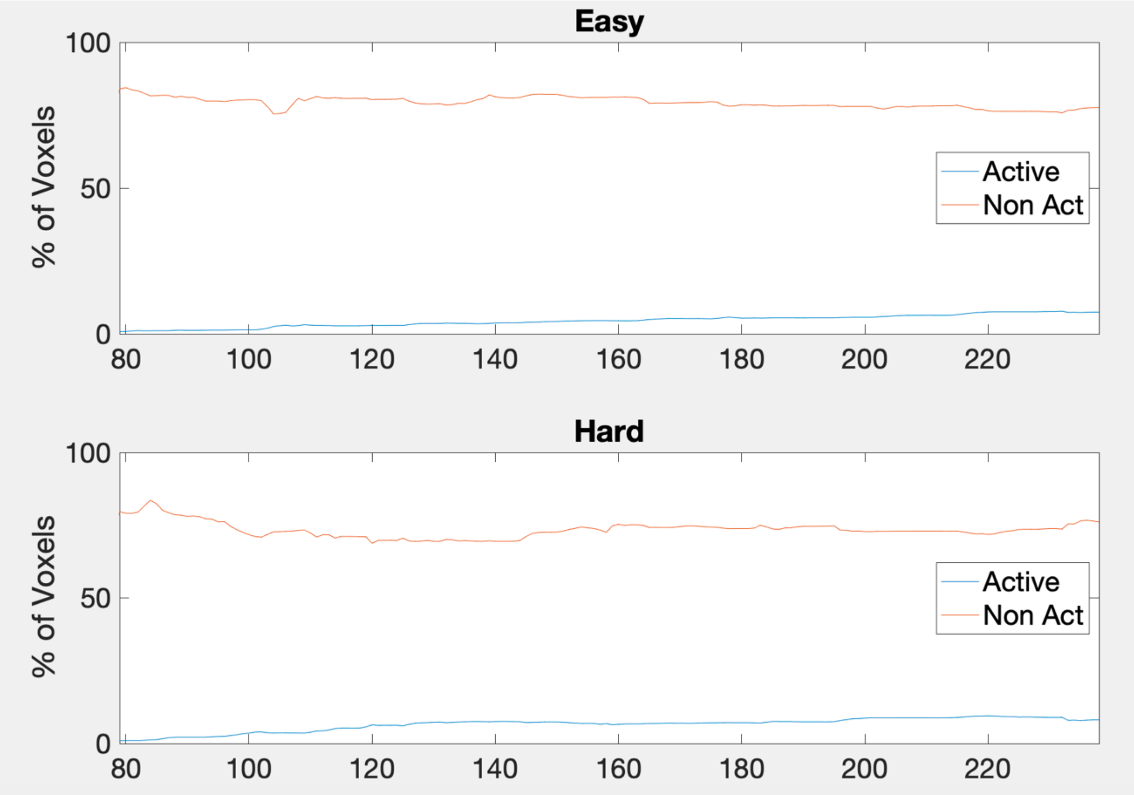Easy<br>% of Voxels<br>Active<br>Non Act<br>Hard<br>% of Voxels<br>Active<br>Non Act |

| | | |
|---|---|---|
| **21** | 104,584 | 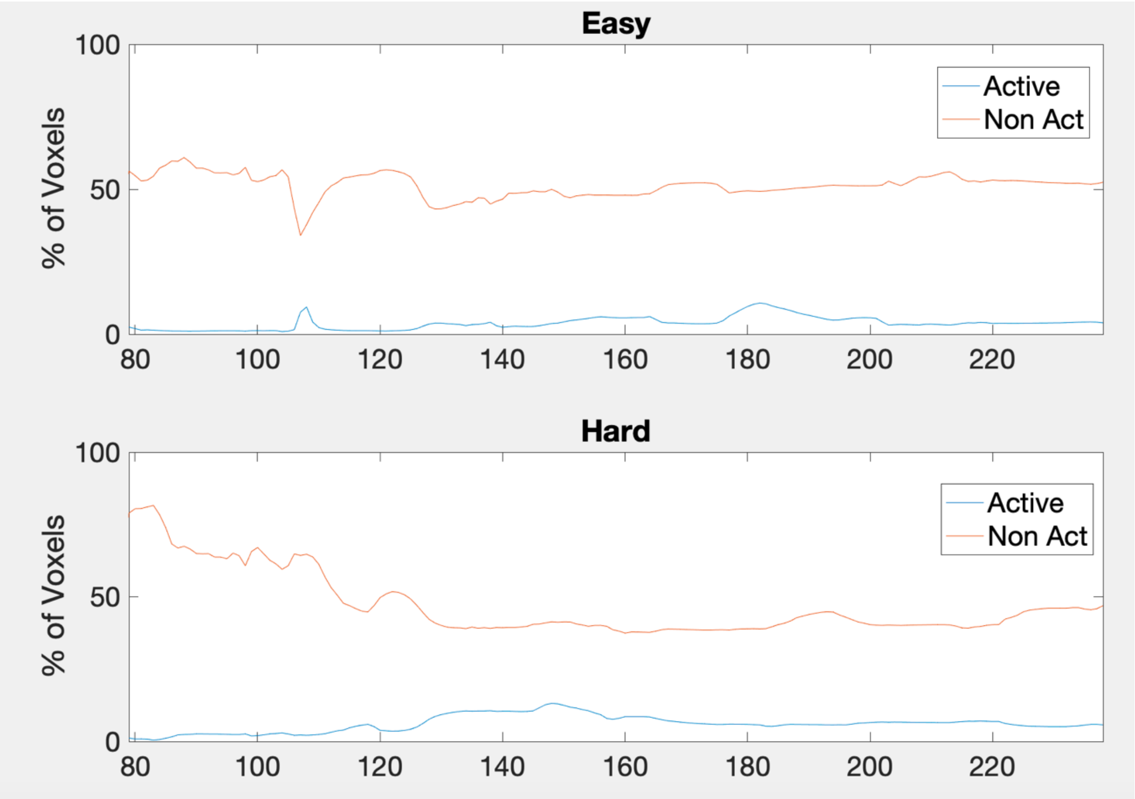 |

Easy
100
50
0
% of Voxels
80
100
120
140
160
180
200
220
Active
Non Act
Hard
100
50
0
% of Voxels
80
100
120
140
160
180
200
220
Active
Non Act

| | | |
|---|---|---|
| **22** | 114,201 | 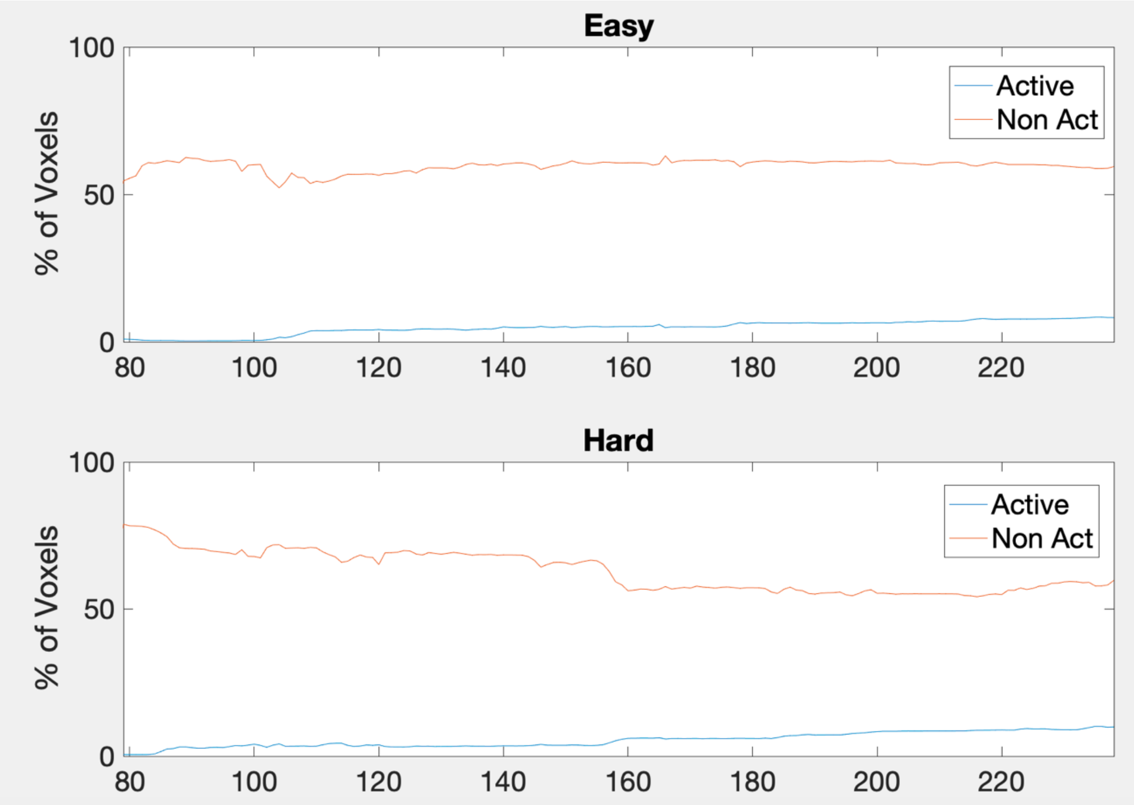Easy<br>100<br>50<br>0<br>% of Voxels<br>80<br>100<br>120<br>140<br>160<br>180<br>200<br>220<br>Active<br>Non Act<br>Hard<br>100<br>50<br>0<br>% of Voxels<br>80<br>100<br>120<br>140<br>160<br>180<br>200<br>220<br>Active<br>Non Act |

| | | |
|---|---|---|
| **23** | 86,177 | 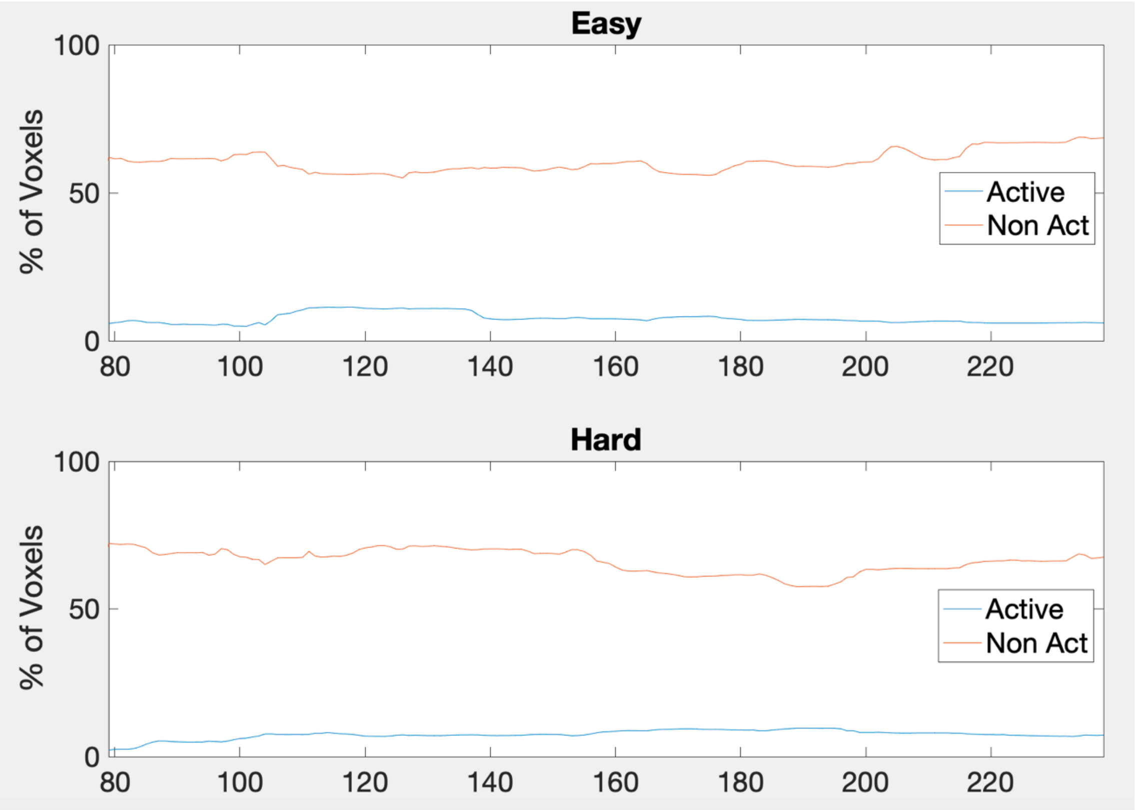 |

**Table S3:** Plots of framewise displacement and subject responses. 1st panel (top): framewise displacement estimation from motion parameters. 2nd panel: Subject responses for easy level, y-axis magnitude indicates time taken to respond. 3rd Panel: hard level subject responses, y-axis magnitude indicates time taken to respond. 4th panel (bottom): the duration of the easy and hard level blocks are shown for reference along with the timing of the expected responses during each block ('Exp' – timing of the expected response).

| **Subject Control** | **Motion and response plots** |
|---|---|

1

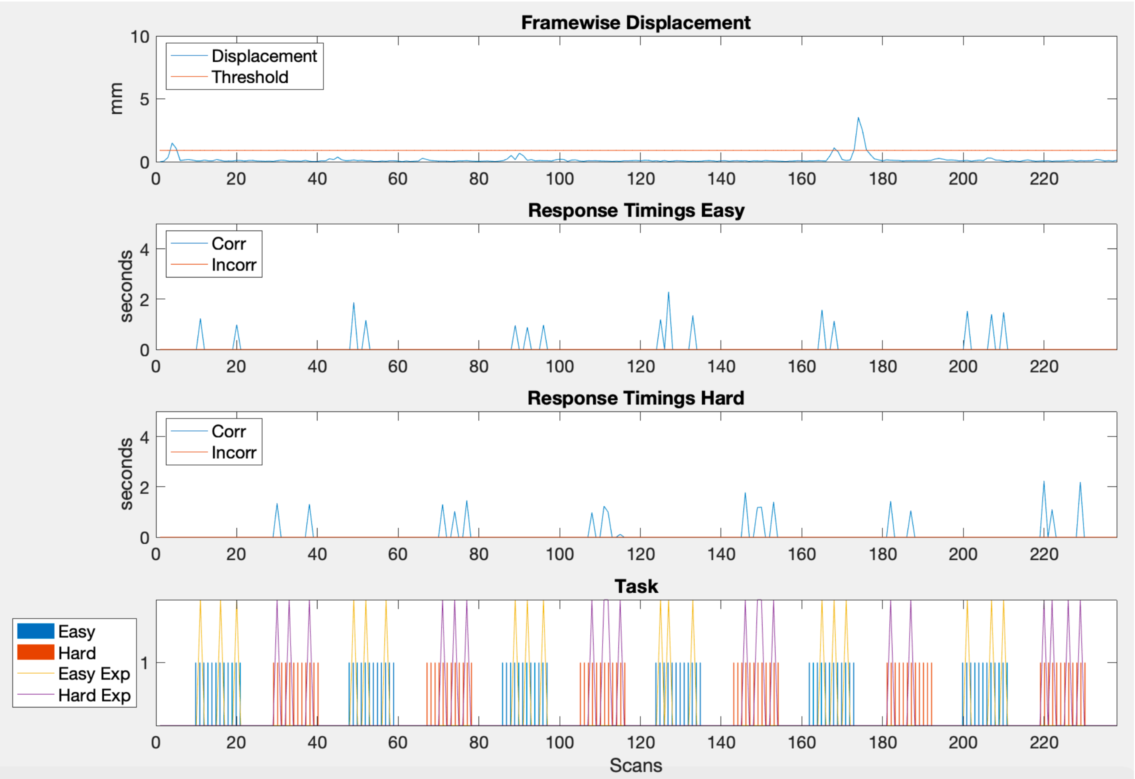
Framewise Displacement
Displacement
Threshold
mm
Response Timings Easy
Corr
Incorr
seconds
Response Timings Hard
Corr
Incorr
seconds
Task
Easy
Hard
Easy Exp
Hard Exp
Scans

2

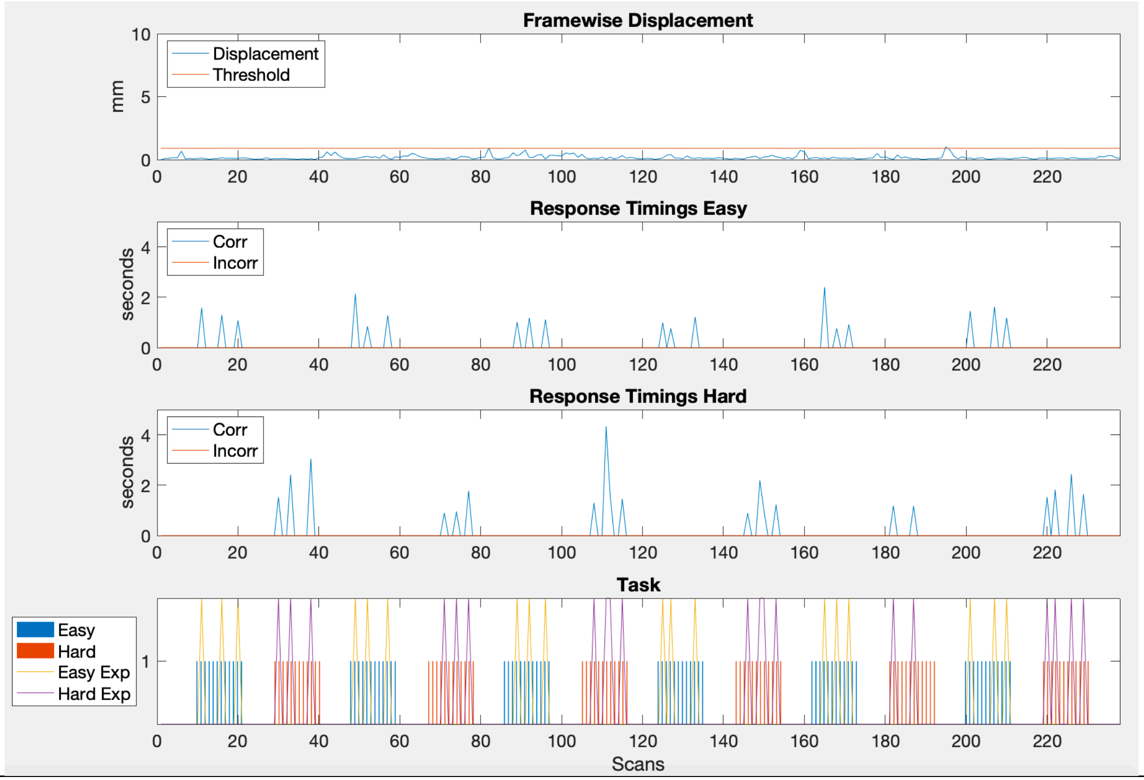
Framewise Displacement
Displacement
Threshold
mm
Response Timings Easy
Corr
Incorr
seconds
Response Timings Hard
Corr
Incorr
seconds
Task
Easy
Hard
Easy Exp
Hard Exp
Scans

| 3 | 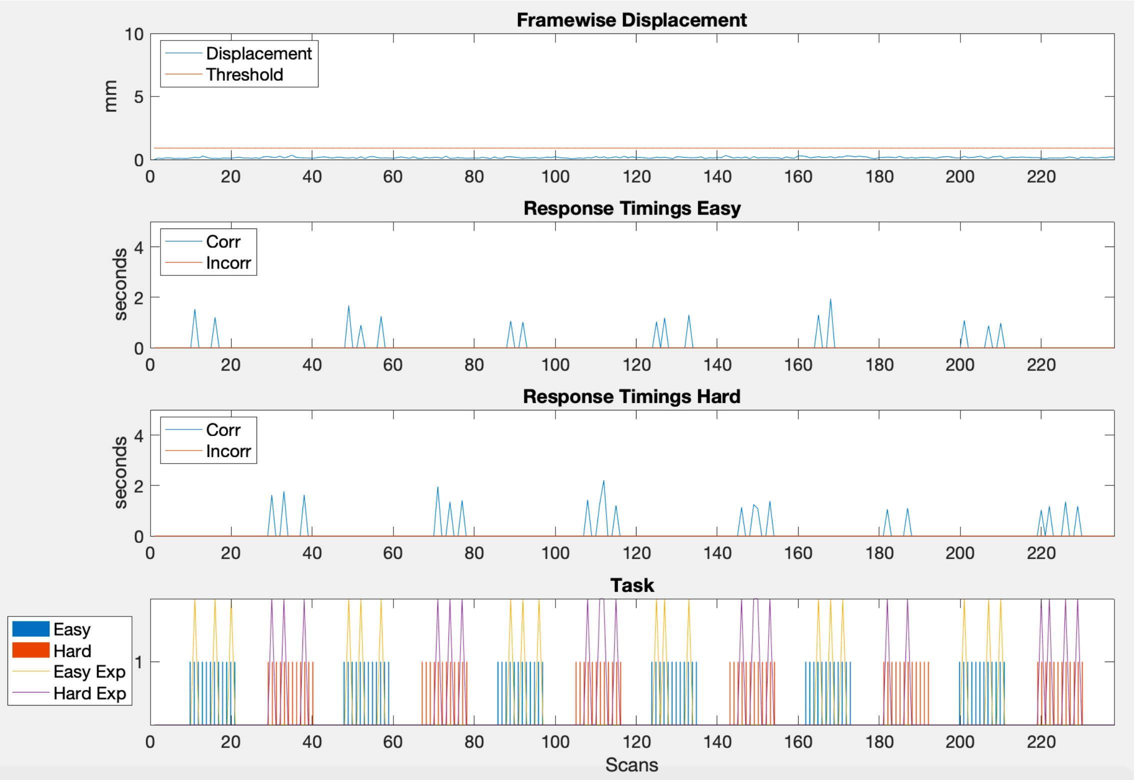Framewise Displacement
Displacement
Threshold
mm
Response Timings Easy
Corr
Incorr
seconds
Response Timings Hard
Corr
Incorr
seconds
Task
Easy
Hard
Easy Exp
Hard Exp
Scans |

4

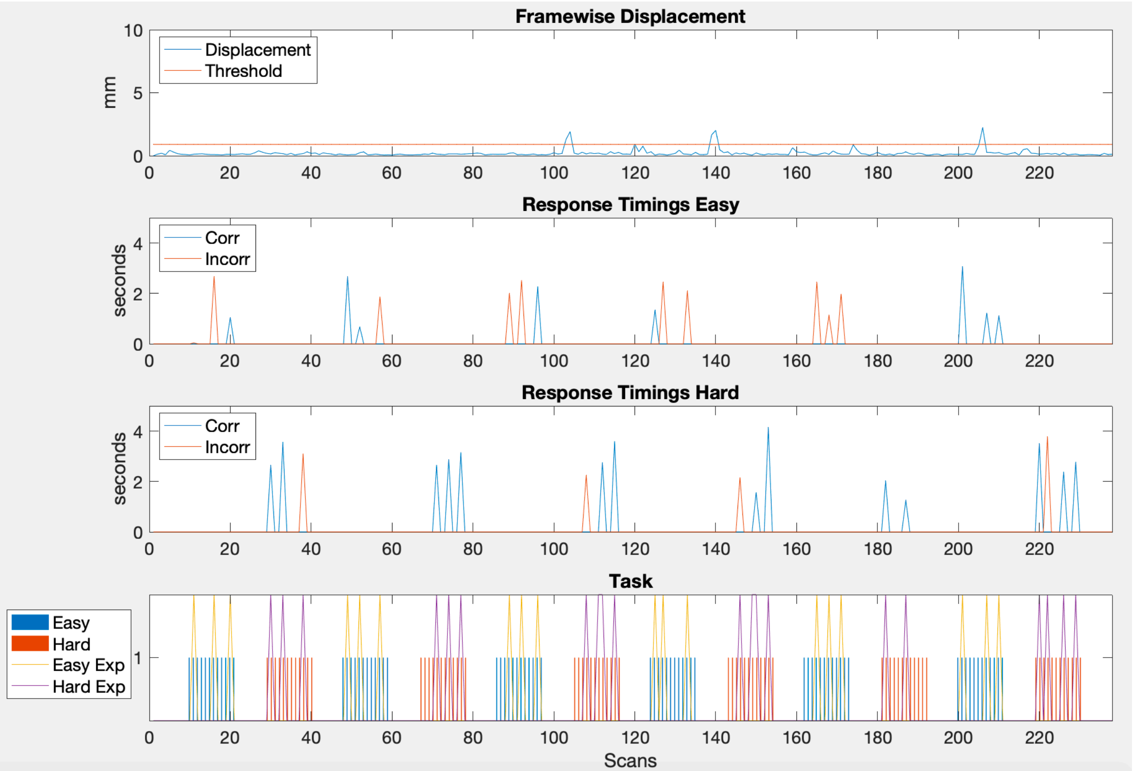
Framewise Displacement
Displacement
Threshold
mm
Response Timings Easy
Corr
Incorr
seconds
Response Timings Hard
Corr
Incorr
seconds
Task
Easy
Hard
Easy Exp
Hard Exp
Scans

| | |
|---|---|
| 5 | 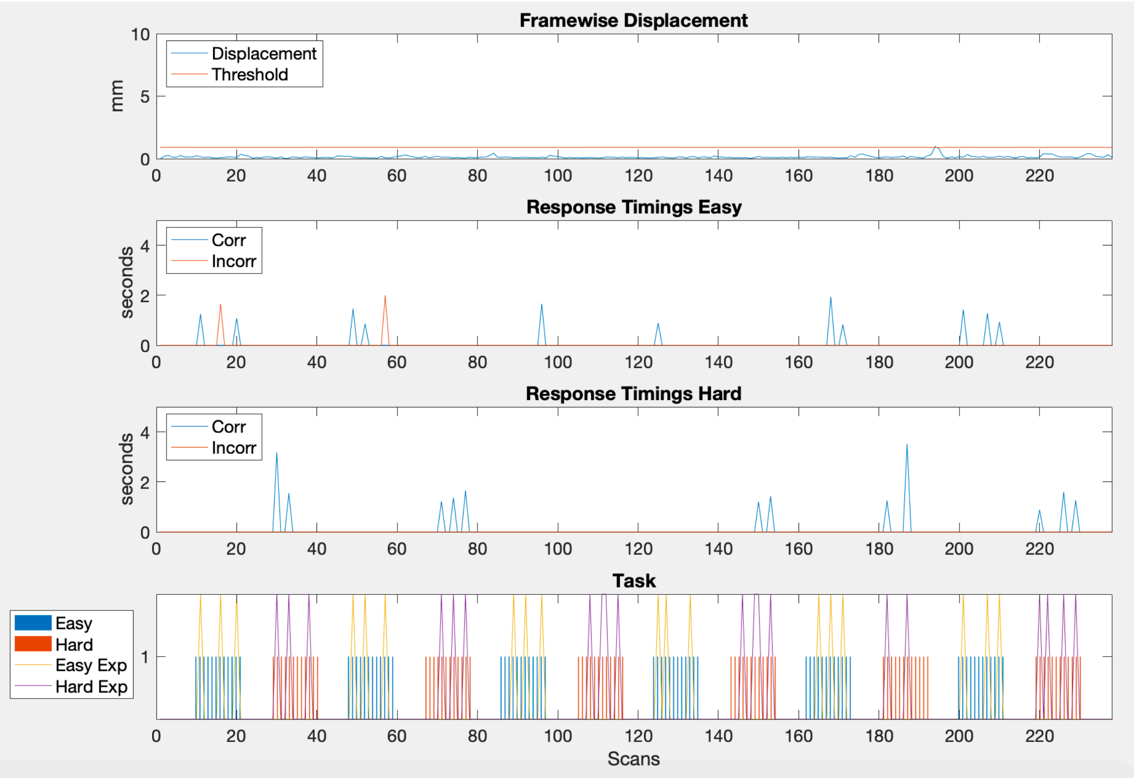 |

Framewise Displacement
Displacement
Threshold
mm
Response Timings Easy
Corr
Incorr
seconds
Response Timings Hard
Corr
Incorr
seconds
Task
Easy
Hard
Easy Exp
Hard Exp
Scans

6

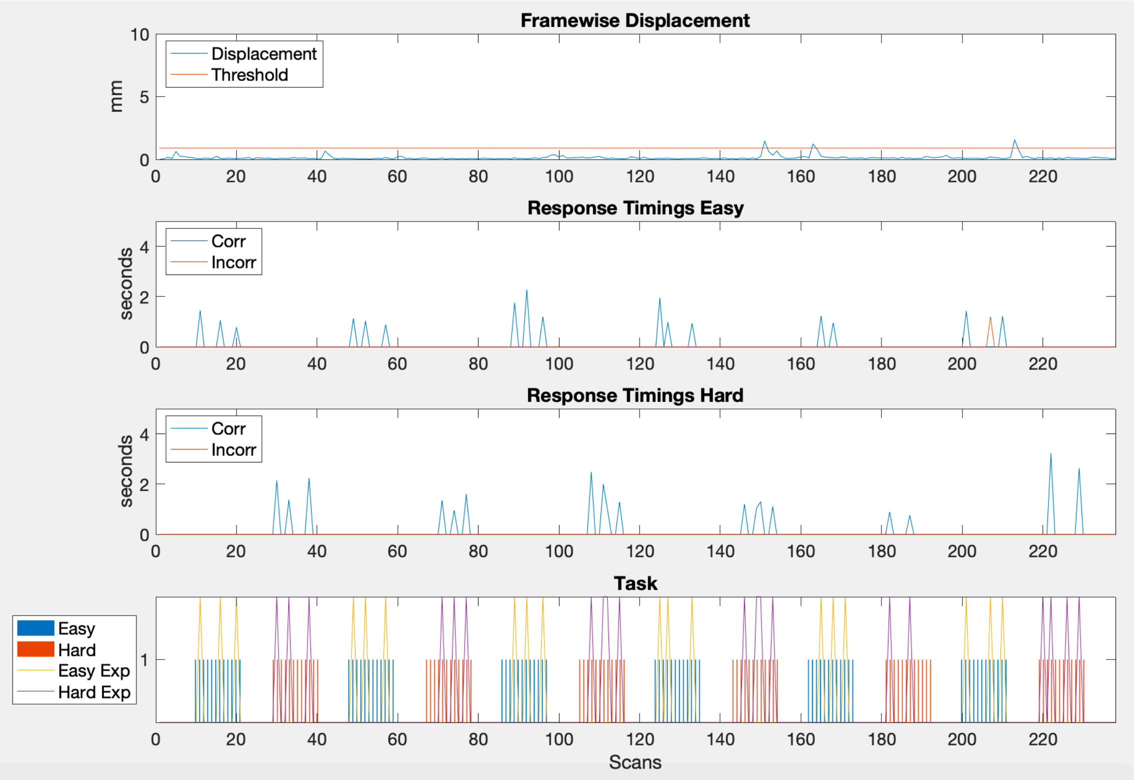

Framewise Displacement
Displacement
Threshold
mm
Response Timings Easy
Corr
Incorr
seconds
Response Timings Hard
Corr
Incorr
seconds
Task
Easy
Hard
Easy Exp
Hard Exp
Scans

7

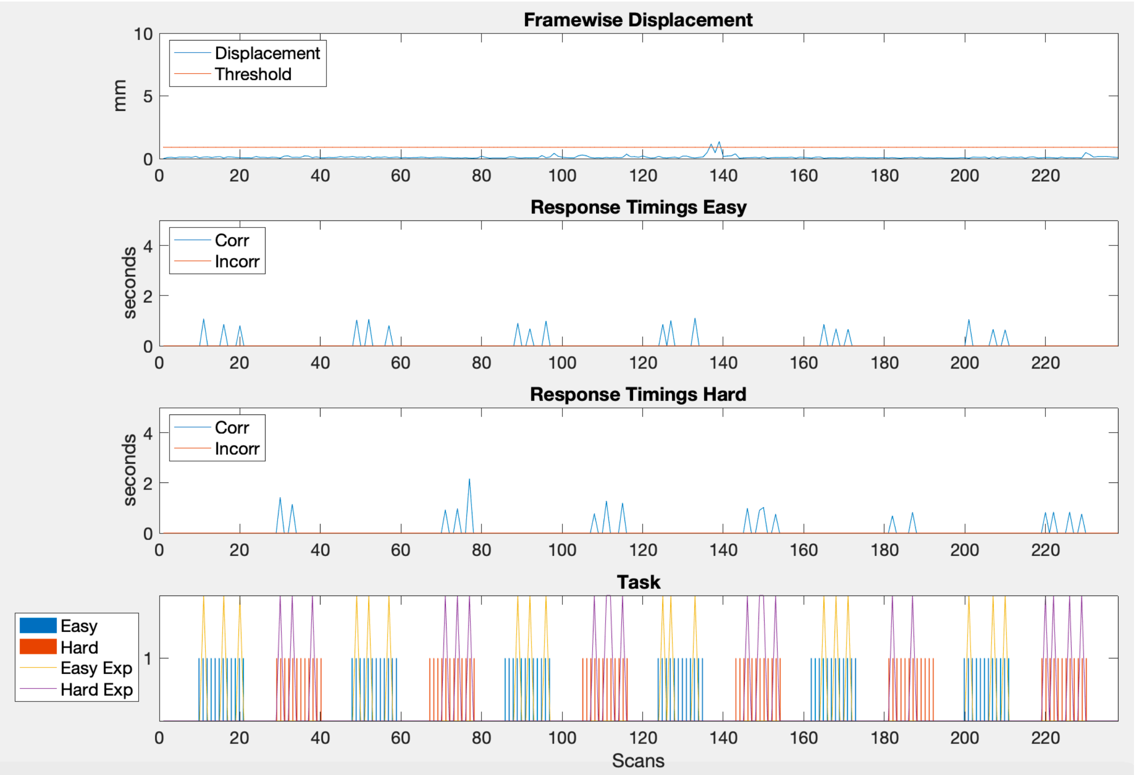
Framewise Displacement
mm
Displacement
Threshold
Response Timings Easy
seconds
Corr
Incorr
Response Timings Hard
seconds
Corr
Incorr
Task
Easy
Hard
Easy Exp
Hard Exp
Scans

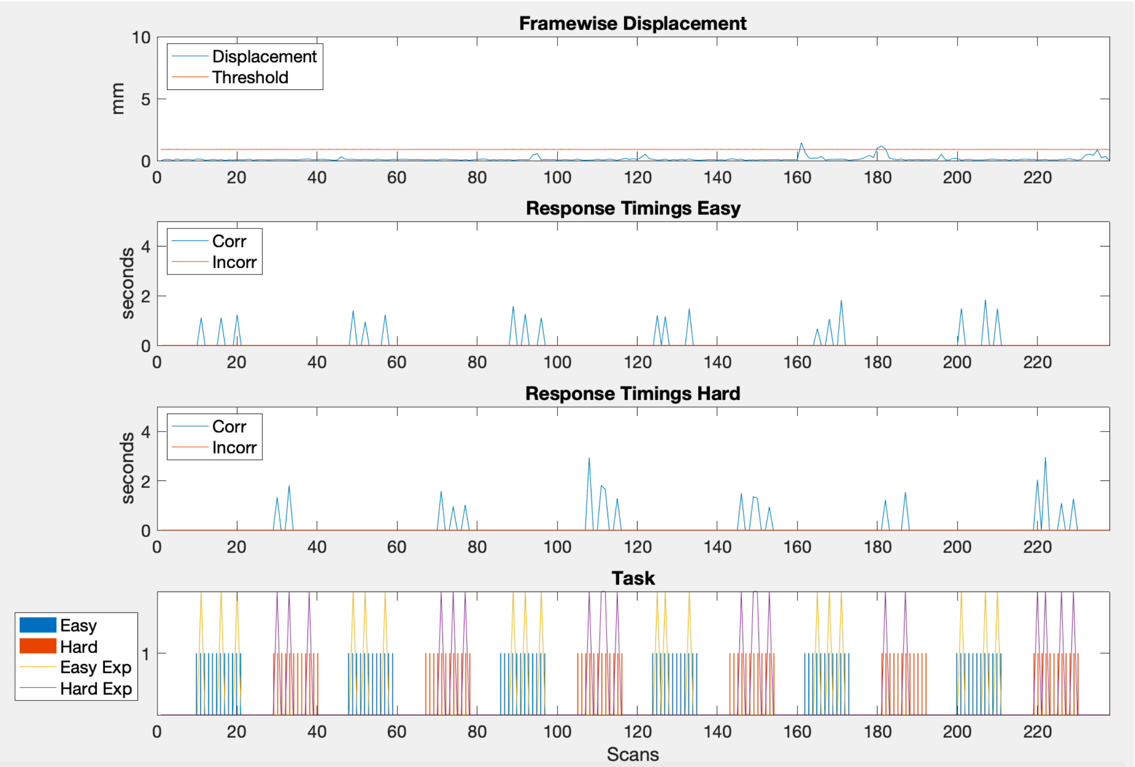
Framewise Displacement
Displacement
Threshold
mm
Response Timings Easy
Corr
Incorr
seconds
Response Timings Hard
Corr
Incorr
seconds
Task
Easy
Hard
Easy Exp
Hard Exp
Scans

9

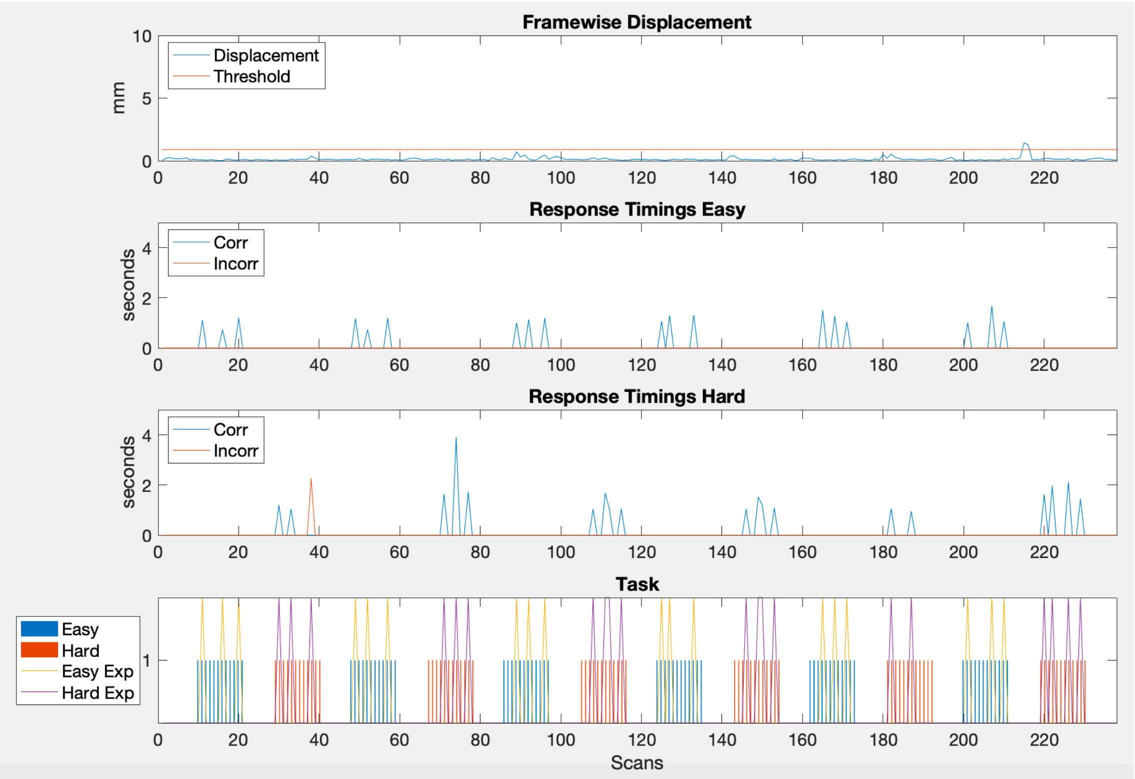

Framewise Displacement
mm
Displacement
Threshold
Response Timings Easy
seconds
Corr
Incorr
Response Timings Hard
seconds
Corr
Incorr
Task
Easy
Hard
Easy Exp
Hard Exp
Scans

**10**

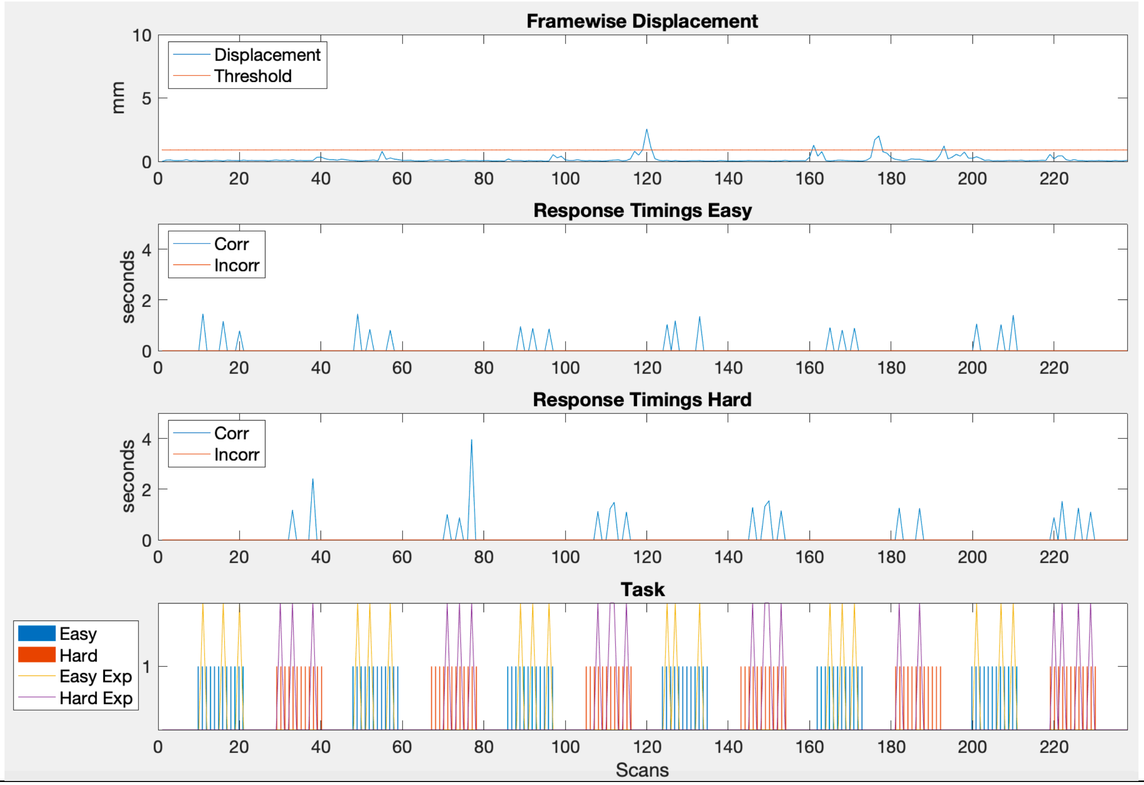

Framewise Displacement
Displacement
Threshold
mm
Response Timings Easy
Corr
Incorr
seconds
Response Timings Hard
Corr
Incorr
seconds
Task
Easy
Hard
Easy Exp
Hard Exp
Scans

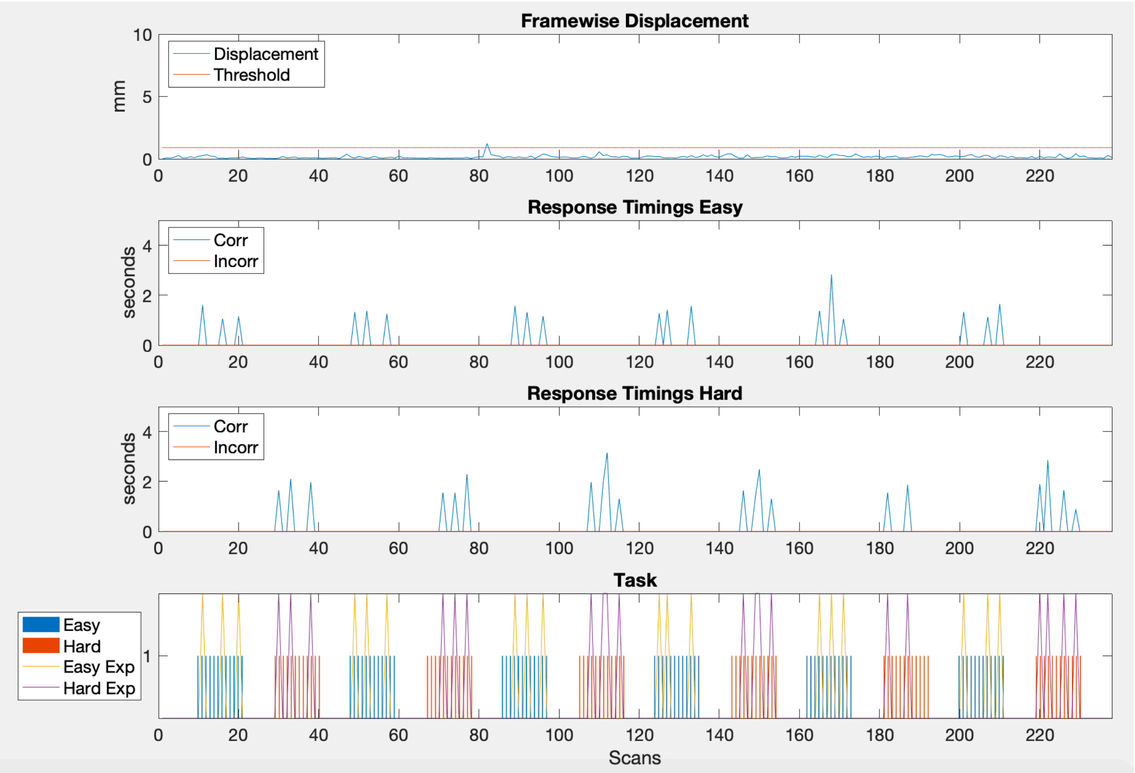
Framewise Displacement
Displacement
Threshold
mm
Response Timings Easy
Corr
Incorr
seconds
Response Timings Hard
Corr
Incorr
seconds
Task
Easy
Hard
Easy Exp
Hard Exp
Scans

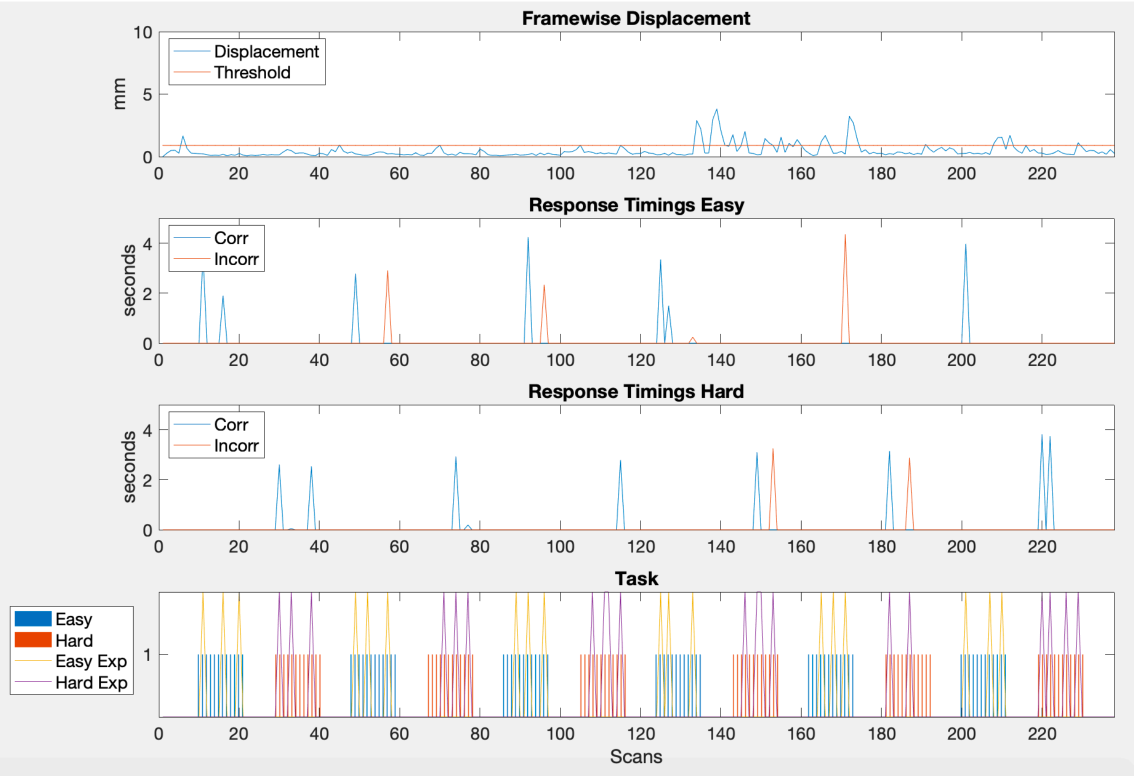
Framewise Displacement
mm
Displacement
Threshold
Response Timings Easy
seconds
Corr
Incorr
Response Timings Hard
seconds
Corr
Incorr
Task
Easy
Hard
Easy Exp
Hard Exp
Scans

**13**

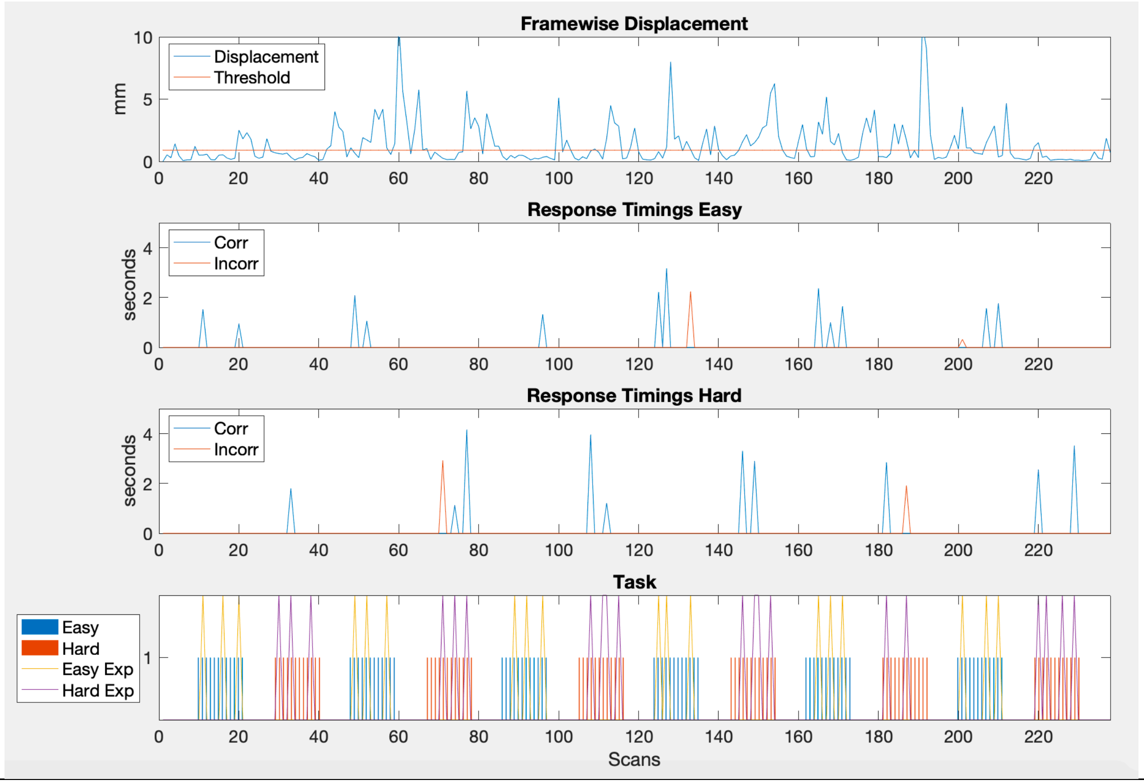

Framewise Displacement
Displacement
Threshold
mm
Response Timings Easy
Corr
Incorr
seconds
Response Timings Hard
Corr
Incorr
seconds
Task
Easy
Hard
Easy Exp
Hard Exp
Scans

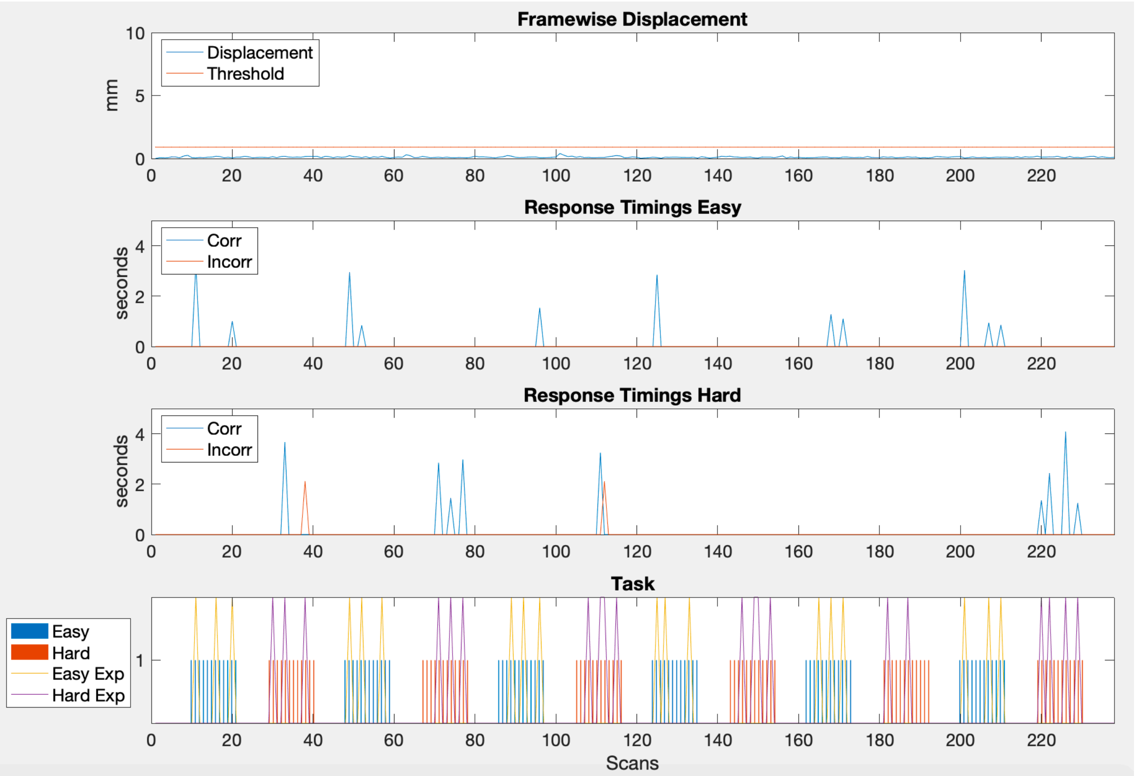
Framewise Displacement
mm
Displacement
Threshold
Response Timings Easy
seconds
Corr
Incorr
Response Timings Hard
seconds
Corr
Incorr
Task
Easy
Hard
Easy Exp
Hard Exp
Scans

15

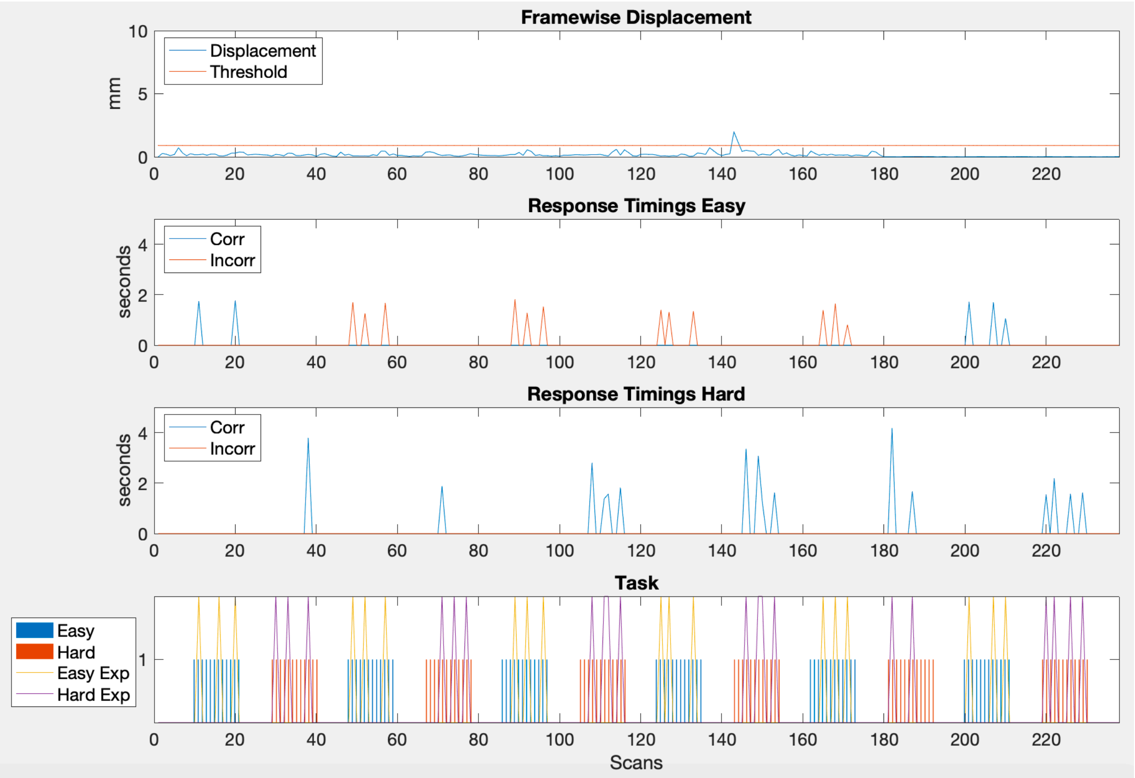

Framewise Displacement
Displacement
Threshold
mm
Response Timings Easy
Corr
Incorr
seconds
Response Timings Hard
Corr
Incorr
seconds
Task
Easy
Hard
Easy Exp
Hard Exp
Scans

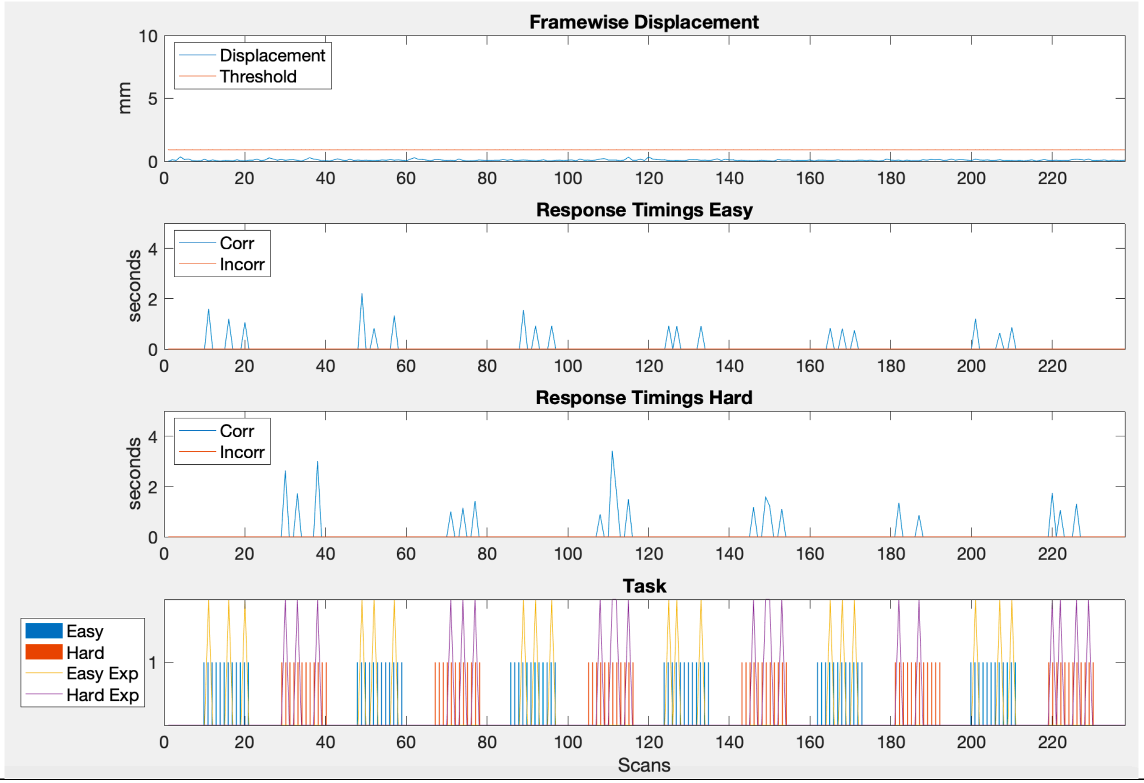
Framewise Displacement
mm
Displacement
Threshold
Response Timings Easy
seconds
Corr
Incorr
Response Timings Hard
seconds
Corr
Incorr
Task
Easy
Hard
Easy Exp
Hard Exp
Scans

17

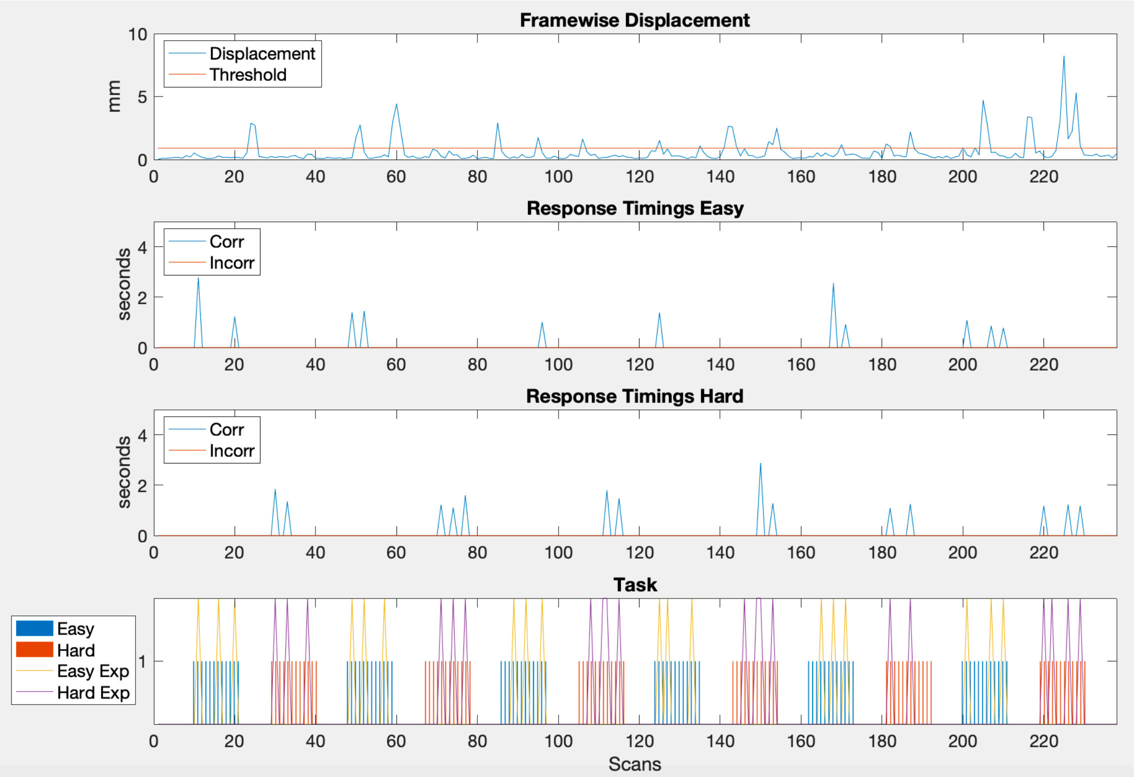
Framewise Displacement
mm
Displacement
Threshold
10
5
0
0 20 40 60 80 100 120 140 160 180 200 220
Response Timings Easy
seconds
Corr
Incorr
4
2
0
0 20 40 60 80 100 120 140 160 180 200 220
Response Timings Hard
seconds
Corr
Incorr
4
2
0
0 20 40 60 80 100 120 140 160 180 200 220
Task
Easy
Hard
Easy Exp
Hard Exp
1
0 20 40 60 80 100 120 140 160 180 200 220
Scans

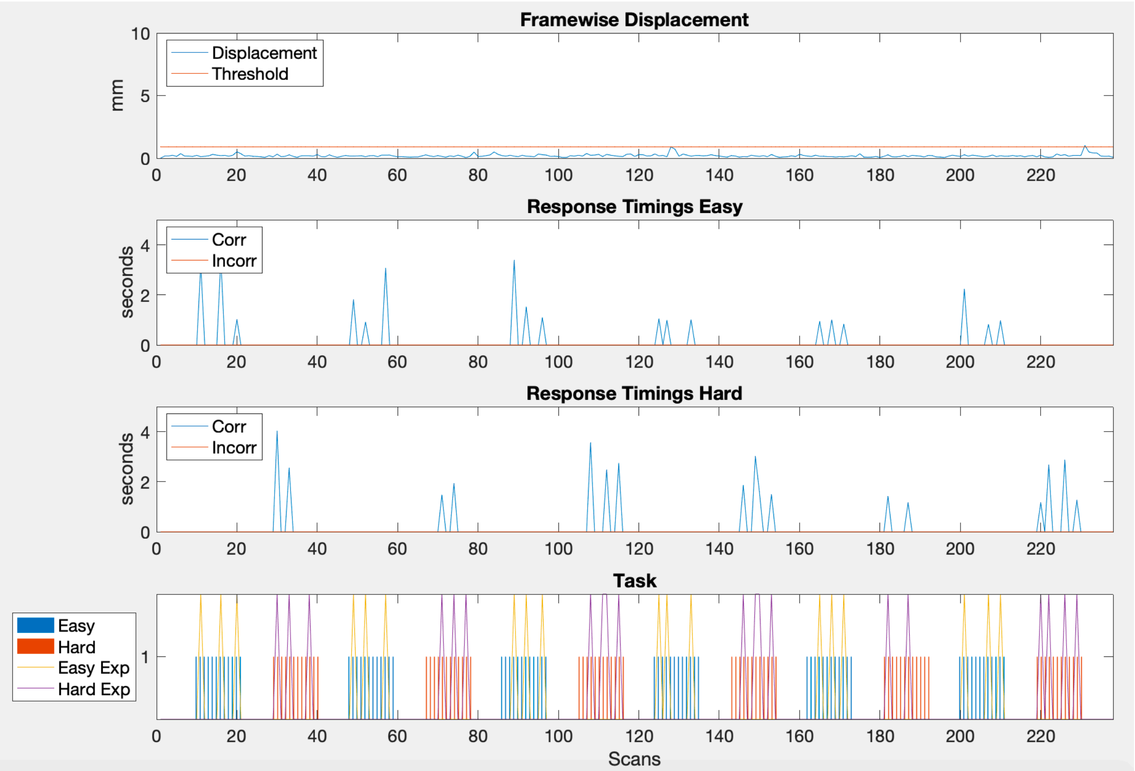
Framewise Displacement
mm
Displacement
Threshold
Response Timings Easy
seconds
Corr
Incorr
Response Timings Hard
seconds
Corr
Incorr
Task
Easy
Hard
Easy Exp
Hard Exp
Scans

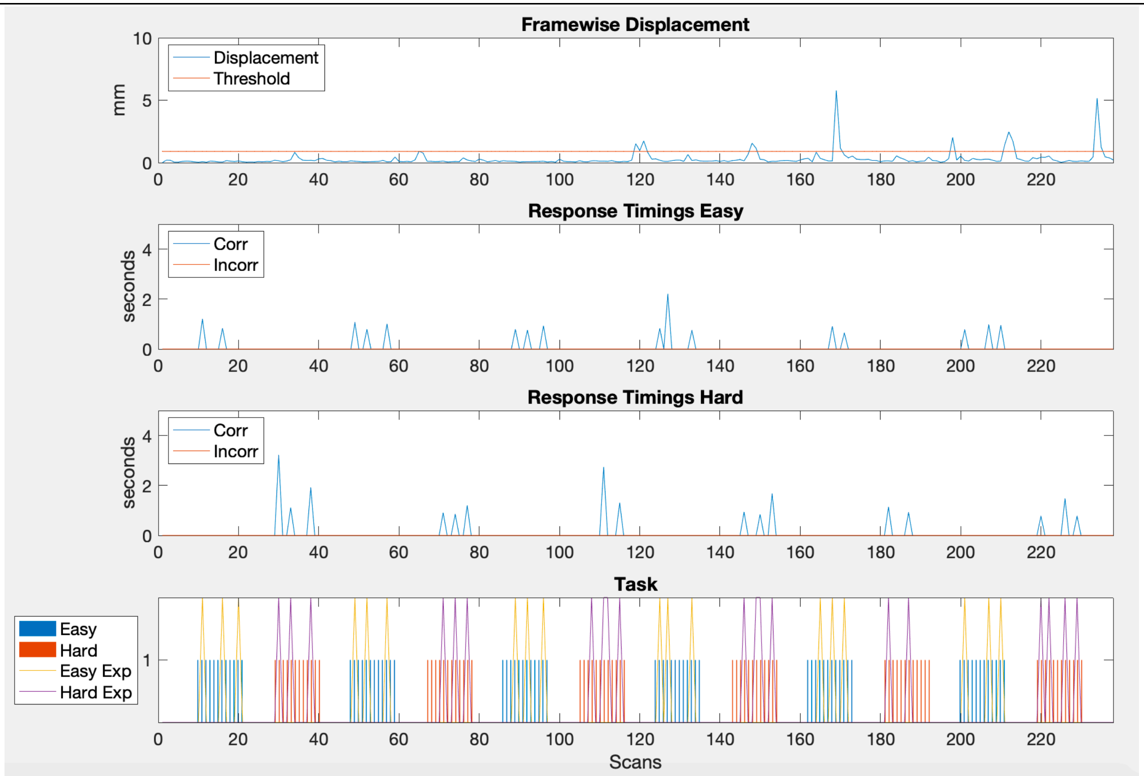
Framewise Displacement
mm
Displacement
Threshold
Response Timings Easy
seconds
Corr
Incorr
Response Timings Hard
seconds
Corr
Incorr
Task
Easy
Hard
Easy Exp
Hard Exp
Scans

**20**

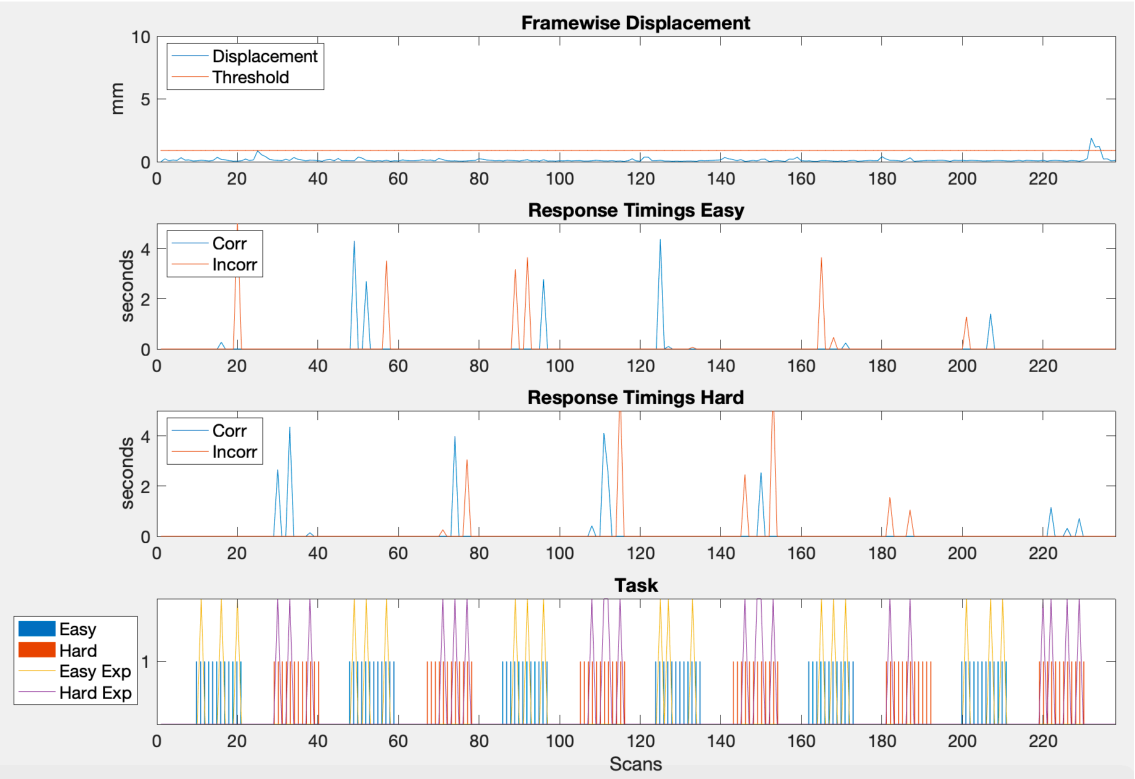

Framewise Displacement
mm
Displacement
Threshold
10
5
0
0 20 40 60 80 100 120 140 160 180 200 220
Response Timings Easy
seconds
Corr
Incorr
4
2
0
0 20 40 60 80 100 120 140 160 180 200 220
Response Timings Hard
seconds
Corr
Incorr
4
2
0
0 20 40 60 80 100 120 140 160 180 200 220
Task
Easy
Hard
Easy Exp
Hard Exp
1
0 20 40 60 80 100 120 140 160 180 200 220
Scans

**21**

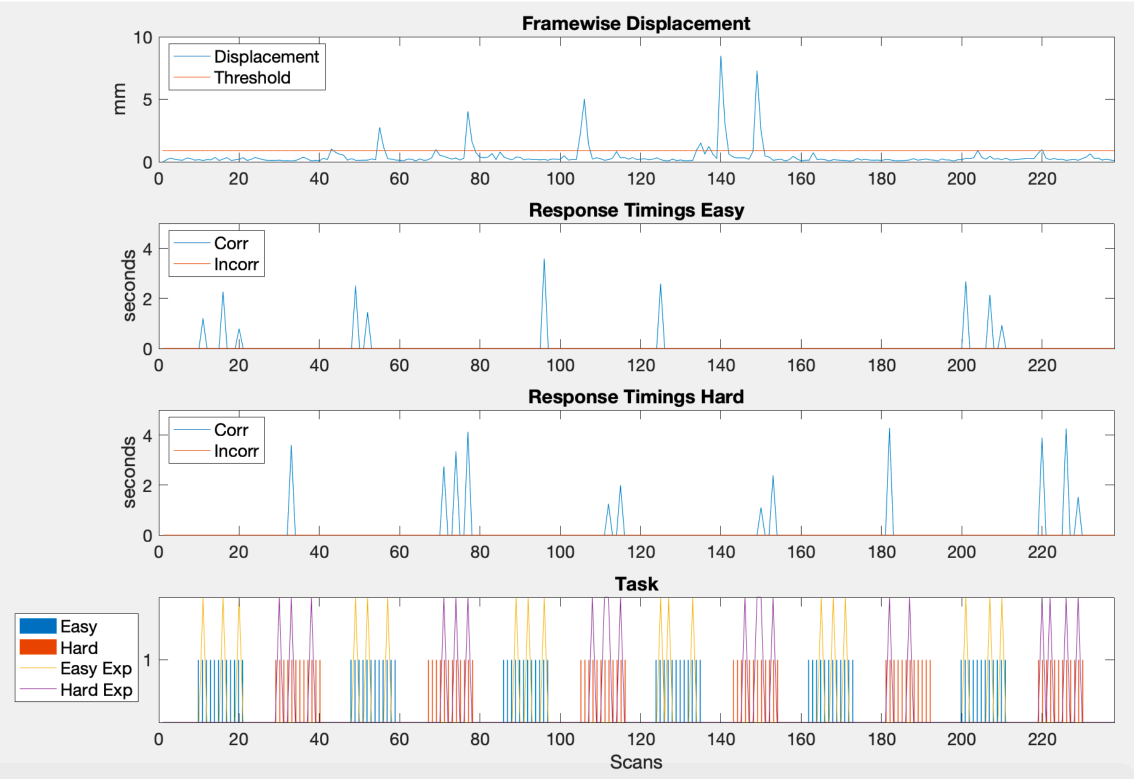
Framewise Displacement
mm
Displacement
Threshold
Response Timings Easy
seconds
Corr
Incorr
Response Timings Hard
seconds
Corr
Incorr
Task
Easy
Hard
Easy Exp
Hard Exp
Scans

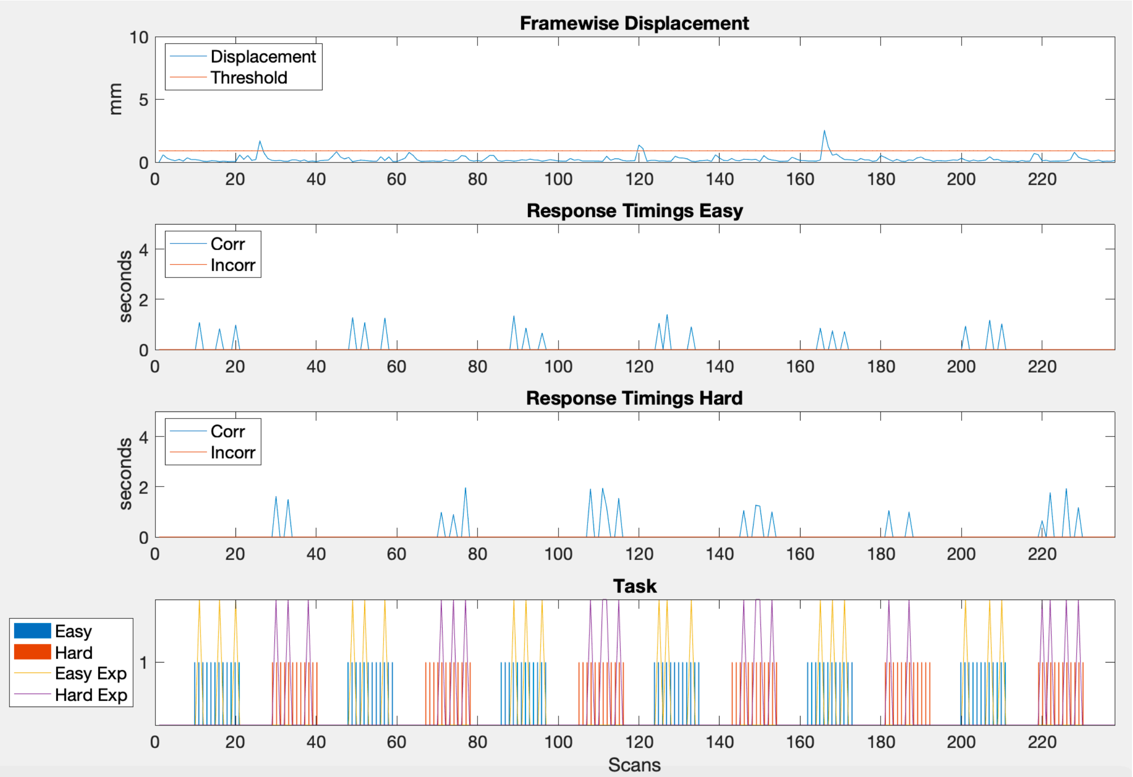
Framewise Displacement
mm
Displacement
Threshold
Response Timings Easy
seconds
Corr
Incorr
Response Timings Hard
seconds
Corr
Incorr
Task
Easy
Hard
Easy Exp
Hard Exp
Scans

23

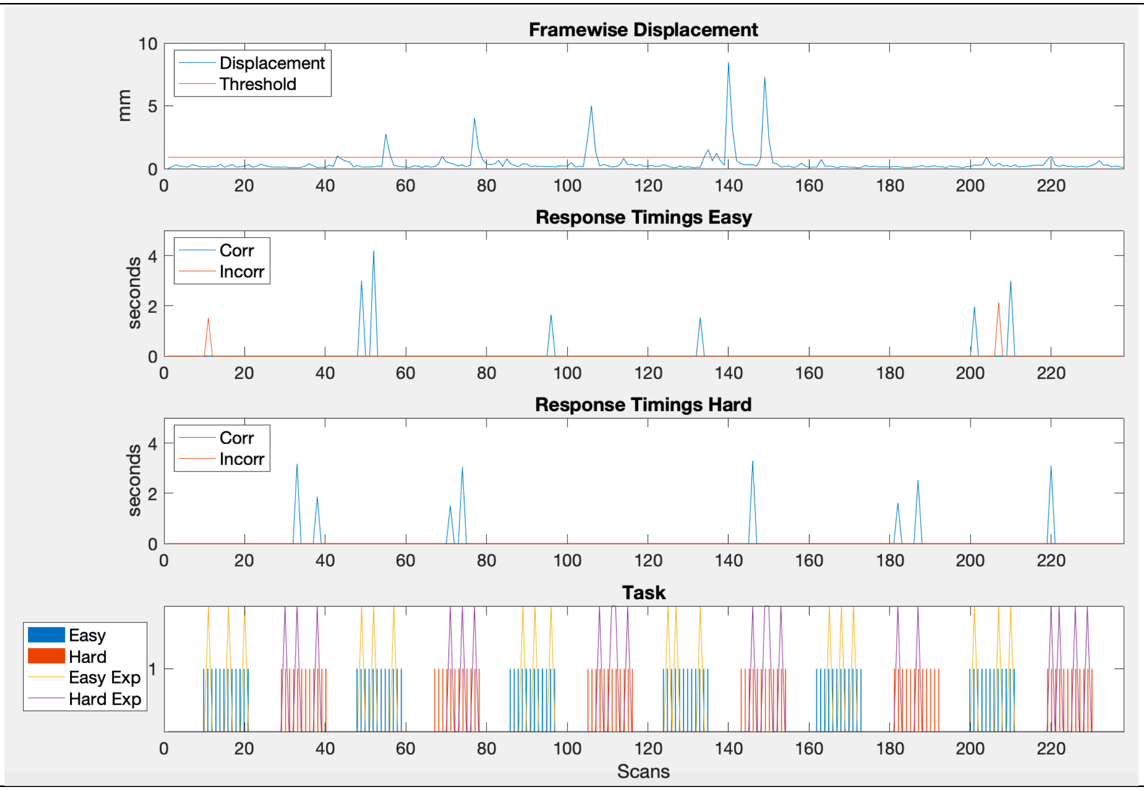

**Table S4:** List of activations from a group analysis using FSL. Easy and hard levels for controls and EPT subjects are reported for early stopping and full scan durations. Thresholds of $p < 0.001$ uncorrected and minimum cluster extent = 10 voxels have been applied.

| Group | Cluster No. | Coordinates (MNI) | No. of Voxels | Peak z-score | Side | Location |
|---|---|---|---|---|---|---|
| **2-Block Early stopping: Easy** | | | | | | |
| **EPT** | 1 | 32 -60 40 | 283 | 7.62 | Right | Precuneus |
| | 2 | -6 8 48 | 274 | 6.32 | Left | Cingulate Gyrus |
| | 3 | -40 16 0 | 30 | 3.67 | Left | Insula |
| | 4 | -54 10 0 | 28 | 6.87 | Left | Superior Temporal Gyrus |
| | 5 | -50 4 38 | 15 | 4.37 | Left | Precentral Gyrus |
| | 6 | -42 8 30 | 14 | 4.89 | Left | Inferior Frontal Gyrus |

| | | | | | | |
|---|---|---|---|---|---|---|
| | 7 | -42 -6 40 | 13 | 4.55 | Left | Precentral Gyrus |
| | 8 | -28 -60 42 | 13 | 6.15 | Left | Precuneus |
| | 9 | 36 16 4 | 11 | 3.87 | Right | Claustrum |
| | 10 | 6 -28 10 | 10 | 3.94 | Right | Thalamus |
| **Control** | 1 | -46 2 28 | 652 | 7.07 | Left | Precentral Gyrus |
| | 2 | 42 -44 46 | 498 | 8.88 | Right | Inferior Parietal Lobule |
| | 3 | -12 16 32 | 292 | 6.44 | Left | Cingulate Gyrus |
| | 4 | -48 -40 40 | 231 | 6.13 | Left | Supramarginal Gyrus |
| | 5 | -8 0 50 | 124 | 5.38 | Left | Cingulate Gyrus |
| | 6 | 20 -32 18 | 96 | 3.84 | Right | Caudate |
| | 7 | -38 14 -2 | 28 | 3.94 | Left | Claustrum |
| | 8 | 40 18 -2 | 12 | 3.39 | Right | Insula |
| | 9 | 50 -28 36 | 11 | 3.44 | Right | Inferior Parietal Lobule |
| | 10 | 36 -56 64 | 10 | 5.2 | Right | Superior Parietal Lobule |
| **Hard** | | | | | | |
| **EPT** | 1 | 8 22 34 | 94 | 8.43 | Right | Cingulate Gyrus |
| | 2 | -8 8 50 | 56 | 6.79 | Left | Medial Frontal Gyrus |
| | 3 | -20 10 30 | 38 | 4.08 | Left | Cingulate Gyrus |
| | 4 | 40 -40 46 | 33 | 7.87 | Right | Inferior Parietal Lobule |
| | 5 | -12 16 34 | 31 | 6.24 | Left | Cingulate Gyrus |
| | 6 | -34 16 28 | 31 | 5.05 | Left | Inferior Frontal Gyrus |
| | 7 | 18 12 62 | 31 | 4.9 | Right | Medial Frontal Gyrus |
| | 8 | -40 0 30 | 29 | 6.9 | Left | Precentral Gyrus |
| | 9 | -36 -4 58 | 19 | 5.52 | Left | Precentral Gyrus |
| | 10 | -38 -44 62 | 15 | 4.51 | Left | Inferior Parietal Lobule |
| | 11 | 32 -46 64 | 10 | 5.06 | Right | Superior Parietal Lobule |
| | 12 | 2 16 56 | 10 | 4.82 | Left | Superior Frontal Gyrus |
| **Control** | 1 | -8 20 30 | 1058 | 7.92 | Left | Cingulate Gyrus |
| | 2 | -46 4 30 | 420 | 10.3 | Left | Precentral Gyrus |
| | 3 | -48 -38 42 | 384 | 5.92 | Left | Supramarginal Gyrus |
| | 4 | 40 -44 44 | 287 | 8.94 | Right | Inferior Parietal Lobule |
| | 5 | -56 22 28 | 177 | 6.9 | Left | Middle Frontal Gyrus |

| | | | | | | |
|---|---|---|---|---|---|---|
| | 6 | 12 -12 28 | 91 | 4.57 | Right | Cingulate Gyrus |
| | 7 | -18 6 66 | 53 | 5.21 | Left | Middle Frontal Gyrus |
| | 8 | -46 -68 -12 | 47 | 4.02 | Left | Fusiform Gyrus |
| | 9 | -8 -12 26 | 22 | 3.53 | Left | Cingulate Gyrus |
| | 10 | -40 -52 62 | 21 | 5.43 | Left | Superior Parietal Lobule |
| | 11 | 46 -64 -10 | 18 | 3.76 | Right | Fusiform Gyrus |
| | 12 | 44 -76 -10 | 15 | 3.71 | Right | Fusiform Gyrus |
| | 13 | 16 6 64 | 14 | 4.58 | Right | Superior Frontal Gyrus |
| **4-Block** | | | | | | |
| **Early stopping:** | | | | | | |
| **Easy** | | | | | | |
| **EPT** | 1 | -6 8 46 | 379 | 7.25 | Left | Cingulate Gyrus |
| | 2 | 32 -60 40 | 251 | 8.73 | Right | Precuneus |
| | 3 | -42 10 30 | 51 | 5.95 | Left | Inferior Frontal Gyrus |
| | 4 | -28 -60 42 | 22 | 7.67 | Left | Precuneus |
| | 5 | 36 18 46 | 21 | 3.83 | Right | Middle Frontal Gyrus |
| | 6 | -52 10 2 | 14 | 5.13 | Left | Superior Temporal Gyrus |
| | 7 | -42 -4 40 | 11 | 4.93 | Left | Precentral Gyrus |
| | 8 | -34 16 4 | 11 | 4.2 | Left | Insula |
| | 9 | 38 18 2 | 11 | 3.49 | Right | Insula |
| **Control** | 1 | -48 2 26 | 700 | 8.09 | Left | Inferior Frontal Gyrus |
| | 2 | 42 -44 46 | 680 | 9.41 | Right | Inferior Parietal Lobule |
| | 3 | -8 2 52 | 675 | 5.66 | Left | Cingulate Gyrus |
| | 4 | -48 -40 40 | 285 | 7.01 | Left | Supramarginal Gyrus |
| | 5 | 20 -32 18 | 181 | 4.53 | Right | Caudate |
| | 6 | -18 -6 26 | 181 | 4.23 | Left | Caudate |
| | 7 | 52 -32 42 | 15 | 3.92 | Right | Inferior Parietal Lobule |
| | 8 | -38 14 -2 | 14 | 3.9 | Left | Claustrum |
| | 9 | 18 -10 24 | 12 | 3.73 | Right | Caudate |
| **Hard** | | | | | | |
| **EPT** | 1 | 36 -42 48 | 342 | 4.99 | Right | Precuneus |
| | 2 | -10 -2 52 | 301 | 4.63 | Left | Medial Frontal Gyrus |

| | | | | | | |
|---|---|---|---|---|---|---|
| | 3 | -14 14 34 | 167 | 6.15 | Left | Cingulate Gyrus |
| | 4 | -44 0 38 | 60 | 5.28 | Left | Precentral Gyrus |
| | 5 | -36 -4 58 | 32 | 3.89 | Left | Precentral Gyrus |
| | 6 | 12 20 28 | 17 | 5.21 | Right | Cingulate Gyrus |
| | 7 | -30 -58 42 | 15 | 4.02 | Left | Angular Gyrus |
| | 8 | -32 16 34 | 14 | 3.85 | Left | Middle Frontal Gyrus |
| **Control** | 1 | -6 20 32 | 731 | 8.54 | Left | Cingulate Gyrus |
| | 2 | 40 -44 44 | 364 | 9.74 | Right | Inferior Parietal Lobule |
| | 3 | -44 12 24 | 335 | 7.29 | Left | Inferior Frontal Gyrus |
| | 4 | -48 -38 42 | 227 | 6.54 | Left | Supramarginal Gyrus |
| | 5 | -14 -6 22 | 75 | 4.14 | Left | Caudate |
| | 6 | -44 34 30 | 46 | 4.05 | Left | Middle Frontal Gyrus |
| | 7 | -46 20 20 | 45 | 5.13 | Left | Middle Frontal Gyrus |
| | 8 | -16 -24 22 | 29 | 4.21 | Left | Caudate |
| | 9 | 40 10 26 | 24 | 4.22 | Right | Precentral Gyrus |
| | 10 | 8 8 24 | 17 | 3.92 | Right | Cingulate Gyrus |
| | 11 | -42 -46 62 | 17 | 5.5 | Left | Inferior Parietal Lobule |
| | 12 | 10 -12 28 | 15 | 4.43 | Right | Cingulate Gyrus |
| | 13 | 16 -22 22 | 13 | 3.79 | Right | Caudate |
| | 14 | 20 -10 26 | 13 | 3.44 | Right | Caudate |
| | 15 | -36 28 22 | 10 | 4.25 | Left | Middle Frontal Gyrus |
| **Full duration:** | | | | | | |
| **Easy** | | | | | | |
| **EPT** | 1 | 28 -56 50 | 150 | 4.93 | Right | Superior Parietal Lobule |
| | 2 | -10 8 44 | 144 | 4.49 | Left | Cingulate Gyrus |
| | 3 | -40 12 32 | 44 | 4.81 | Left | Precentral Gyrus |
| | 4 | 52 20 38 | 28 | 5.48 | Right | Middle Frontal Gyrus |
| | 5 | 32 18 44 | 28 | 4.54 | Right | Middle Frontal Gyrus |
| | 6 | -30 -60 44 | 17 | 4.30 | Left | Angular Gyrus |
| | 7 | 8 30 38 | 14 | 4.16 | Right | Cingulate Gyrus |
| | 8 | 46 14 28 | 13 | 5.75 | Right | Precentral Gyrus |

| | | | | | | |
|---|---|---|---|---|---|---|
| **Control** | 1 | -46 0 26 | 754 | 6.52 | Left | Inferior Frontal Gyrus |
| | 2 | 40 -44 46 | 612 | 9.21 | Right | Inferior Parietal Lobule |
| | 3 | -12 14 36 | 573 | 6.05 | Left | Cingulate Gyrus |
| | 4 | -36 -46 44 | 225 | 9.53 | Left | Inferior Parietal Lobule |
| | 5 | 20 -30 18 | 65 | 5.06 | Right | Caudate |
| | 6 | -40 18 -2 | 17 | 3.62 | Left | Insula |
| **Hard** | | | | | | |
| **EPT** | 1 | -14 14 34 | 446 | 6.78 | Left | Cingulate Gyrus |
| | 2 | 36 -42 48 | 418 | 5.93 | Right | Precuneus |
| | 3 | -44 2 38 | 66 | 5.09 | Left | Precentral Gyrus |
| | 4 | -30 -58 42 | 40 | 4.31 | Left | Angular Gyrus |
| | 5 | -50 22 24 | 15 | 3.65 | Left | Inferior Frontal Gyrus |
| | 6 | -32 16 34 | 15 | 3.98 | Left | Middle Frontal Gyrus |
| | 7 | 8 0 68 | 14 | 4.2 | Right | Medial Frontal Gyrus |
| | 8 | 12 4 28 | 11 | 4.26 | Right | Cingulate Gyrus |
| | 9 | -10 -4 66 | 11 | 3.68 | Left | Medial Frontal Gyrus |
| | 10 | 40 10 26 | 10 | 4.25 | Right | Precentral Gyrus |
| **Control** | 1 | -10 14 36 | 680 | 7.70 | Left | Cingulate Gyrus |
| | 2 | 38 -44 44 | 590 | 9.12 | Right | Inferior Parietal Lobule |
| | 3 | -34 -46 44 | 277 | 9.47 | Left | Inferior Parietal Lobule |
| | 4 | -44 0 28 | 259 | 5.16 | Left | Precentral Gyrus |
| | 5 | -46 22 22 | 128 | 5.00 | Left | Middle Frontal Gyrus |
| | 6 | -16 -24 22 | 68 | 4.02 | Left | Caudate |
| | 7 | 20 -32 18 | 63 | 3.94 | Right | Caudate |
| | 8 | -40 16 -6 | 45 | 5.62 | Left | Insula |
| | 9 | -22 6 66 | 12 | 3.43 | Left | Superior Frontal Gyrus |
| | 10 | 0 -86 6 | 11 | 3.69 | Left | Lingual Gyrus |
